# Comparative efficacy of exercise therapy for patellofemoral pain: A network meta-analysis of randomized controlled trials

**DOI:** 10.1101/2023.01.01.22284033

**Authors:** Yu-Xiu Ji, Yu-Jie Xie, Xiao-Zhu Zhou, Shuang Wang, Ling Li, Akira Miyamoto, Chi Zhang

**Affiliations:** Rehabilitation Medicine Department, The Affiliated Hospital of Southwest Medical University, Luzhou, 646000, Sichuan, People’s Republic of China; Chinese Evidence-based Medical and Cochrane China Center, West China Hospital, Chengdu, 610041, Sichuan, People’s Republic of China; Department of Physical Therapy Faculty of Rehabilitation of Kobe International University, Japan

**Author notes:** Corresponding author: Chi Zhang. Co-first author: Yu-Jie Xie.

**Keywords:** biomechanical phenomena, knee, patellofemoral pain, exercise, osteoarthritis

## Abstract

**Objective:** To investigate the effective exercise prescription in randomized controlled trials (RCTs) for patellofemoral pain (PFP).

**Design:** A network meta-analysis.

**Data sources:** PubMed (including Medline), Embase, Web of Science, PEDro, Clinicaltrials.gov and other resourses for RCTs.

**Eligibility criteria for selecting studies:** RCTs of exercise interventions for PFP with outcomes of pain intensity or functional improvement.

**Primary outcome measure:** Pain intensity is measured by ‘worst pain in the past week’ on a Visual Analogue Scale (VAS) or Numerical Rating Pain Scale (NRS).

**Data extraction:** Two researchers independently extracted data and assessed the bias of risks. We used Grading of Recommendations, Assessment, Development, and Evaluation to appraise the strength of the evidence.

**Results:** A total of 45 trials with 42,319 patients were included in this network meta-analysis (NMA). For the primary outcomes, all included treatments were superior to a wait-and-see approach: PNF + exercise (SMD -2.88, 95%CIs -4.75 to -1.02), whole body exercise (-1.57, -3.15 to -0.00), hip-and knee-focused exercise therapy (-1.32, -2.57 to -0.06), foot orthoses + exercise (-1.06, -2.92 to -0.06), hip exercise (-1.10, -2.44 to 0.24), knee brace + exercise(-0.91, -2.54 to 0.72), gait retraining exercise (-2.55, -4.72 to -0.37), knee exercise (-0.92, -2.16 to 0.33), knee arthroscopy + exercise (-0.61, -2.44 to 1.22), target exercise (-0.52, -2.38 to 1.33), kinesiotaping + exercise (-0.54, -2.07 to 0.99), education + exercise (-0.47, -2.31 to 1.38), feedback exercise (-0.22, -1.86 to 1.43). Exercise therapy with education (SMD -0.25, 95%CIs -1.76 to 1.26) was better than exercise alone in alleviating pain intensity.

**Conclusion:** The knee and hip combination strength training is highly effective in muscle strength improvement. All treatments in our NMA were superior to nontreatment, we recommend avoiding a wait-and-see approach. Comprehensive therapy based on individual evaluation can effectively improve the symptoms of patients.

**Key points (what are the new findings?):**

- The combination of knee and hip strength training is highly effective in pain relief and function improvement for PFP.
- The exercise therapy with biomechanical devices, such as foot orthoses, braces, and kinesiotaping can improve knee function.
- Education with exercise therapy shows a great effect on pain relief and function improvement than exercise alone.
- Target exercise based on individual evaluation is superior to general exercise therapy for keen function.

## Introduction

Patellofemoral pain (PFP) is a common musculoskeletal disorder clinical condition.^1^ It is characterized by retropatellar and/or peripatellar pain associated with lower limb loading.^2^ PFP is a common pathology with an annual prevalence reported as 23%.^3, 4^ More than 50% of PFP individuals experience persistent pain after 12 months, even after being offered evidence-based treatment.^5-8^ PFP has a significant impact on health-related quality of life and burden of life.^9^

One of the most common causes of PFP is biomechanical dysfunction, which involves abnormal joint alignment, trochlear morphology, and muscle weakness.^3, 10-12^ Biomechanical dysfunction has a variety of treatments, such as muscle strengthening exercises, biomechanical devices, biofeedback exercises, manual therapy, and gait retraining exercise.^3, 5^ A recent PFP consensus statement recommends exercise therapy as the cornerstone of treatment.^5^ However, the comparative effectiveness of all available exercise therapy had never been examined. There was no detailed recommendation on exercise therapy for the PFP, such as exercise type, frequency, intensity, mode, time, and rest intervals.^13^ How to choose the most effective exercise therapy for PFP is still a challenge.^14^

This study used network meta-analysis (NMA) to compare the effectiveness of all available exercise therapy for PFP. We aim to provide new evidence for exercise therapy on PFP.

## Methods

### Protocol registration

The systematic review with NMA was prospectively registered on PROSPERO. The findings are reported following the Preferred Reporting Items for Systematic Reviews and Meta-Analyses checklist extension for NMA.^15^

### Administration, dissemination, and updating of the NMA

This review will be administered at Southwest Medical University, Sichuan Province, China, and we plan to update the NMA annually for a minimum of 5 years. As described in our protocol, we will screen the literature annually to identify new data that may alter our conclusions and recommendations. When new data have become available, we will update the analysis and present the updated findings on the website of Southwest Medical University. This study will also provide a plain-language summary for patients and clinicians dealing with PFP.

### Patient and public involvement

#### Primary outcome measure

Pain intensity is measured by “worst pain in the past week” on a Visual Analogue Scale (VAS; 0–10/0–100) or Numerical Rating Pain Scale (NRS; 0–10/0–100). The reliability is excellent, ICC=0.76.^16^

#### Secondary outcome measures

knee function in activities is measured by questionnaires, such as the anterior knee pain scale (APKS). The AKPS has demonstrated high test-retest reliability and appears to be responsive to clinical changes in patients with PFP.^16^

#### Research question

Which exercise therapy is most effective for PFP?

#### Eligibility criteria

##### Type of studies

Randomized controlled trials (RCTs).

##### Type of population

All patients with a clinical diagnosis of PFP were included. Studies were included if they used synonyms for PFP, but as a minimum criterion, described patients with retropatellar or peripatellar pain, of at least 6-week duration, and a non-traumatic onset. The diagnostic criteria used in the original studies were followed, given that pain was described as being retropatellar or peripatellar pain. Studies examining other conditions were excluded (eg, patellar dislocations, patellofemoral osteoarthrosis, patellar tendinopathy, Osgood-Schlatter, iliotibial band syndrome, Sinding-Larsen-Johansson syndrome). Trials that included participants diagnosed with PFP, but with concomitant pain around the patella caused by other conditions (eg, patellar tendinopathy), were eligible for inclusion.

##### Type of treatments and control treatments

Exercise therapies were eligible for inclusion. Specifically, only RCTs evaluating an exercise intervention for treating PFP were included. Any exercise intervention applied on its own, or in combination with other non-surgical interventions was included, providing the other intervention was also applied to the control group.

##### Type of outcomes

Because the course of exercise therapy is not less than one cycle, so studies assessing the treatment effect after a minimum of 4 weeks were included.

Studies assessing the primary or secondary outcomes above were included.

#### Search strategy

We developed a sensitive search strategy that included a mix of indexed and free-text terms (see Winters ^13^ et al). No restrictions (eg, language or full-text availability) were applied. We searched Embase, PubMed (including Medline), Web of Science, PEDro, Clinicaltrials.gov and other resourses. The sources were searched from their date of inception up till 30 April 2021. For unpublished or ongoing studies, we searched the WHO International Clinical Trials Registry Platform Clinical Trials.gov. Finally, we screened reference lists of all Cochrane reviews on PFP and the reports included in this review for possible relevant studies that were not identified by our search.

#### Study selection

Two review authors (X Zeng And XZ Zhou) selected potentially eligible articles by reviewing the title and abstract of each citation. The consensus was sought in cases of initial disagreement.

The report was included for full-text evaluation if consensus could not be reached. After obtaining full articles, both authors independently performed study selection. In case of disagreement, a consensus was sought; however, a third author (L Li) arbitrated the decision if disagreement persisted.

#### Data extraction

Data were extracted by two researchers using standardized extraction forms adapted from the Cochrane Collaboration.^17^ Any disagreements were resolved by consensus. We extracted the following data:

1. Publication and study details: Authors, year of publication, design, and unit of allocation.
2. Population: Number of patients included, population characteristics for age, sex, height, weight, body mass index, baseline scores for outcome measures (mean, SDs, standard errors extracted for continuous outcomes and number and percentage for categorical outcomes).
3. Eligibility criteria and diagnostic criteria used for PFP.
4. Treatments: Number randomized to the group, detailed description, for example, application, dose, intensity, frequency, number of sessions, delivery, tailoring (individual/group), duration of treatment, follow-up. We used items from the Template for Intervention Description and Replication checklist^13, 18^ to assure comprehensive data extraction in this section of the extraction form.^19^
5. Outcomes: Timepoints measured, and the timepoints reported on, outcome definition, the person measuring, unit of measurement, scales (upper and lower limits), imputation of missing data, primary and secondary outcomes used in the original trials, unintentional outcomes.
6. Data and analysis: Comparisons, outcomes, subgroups, timepoints; mean for both groups, mean difference (MD), SDs/95 CIs/standard errors), statistical methods used and appropriateness of these.
7. Other information: Key conclusions of study authors.

#### Risk of bias assessment

The Cochrane Risk of Bias was used to assess the risk of bias. We assessed bias following the intention-to-treat principle.^20^ We assessed the following domains: random sequence generation; allocation concealment; blinding of participants and personnel; blinding of outcome assessment; incomplete outcome data; selective reporting; and other biases. Other sources of bias included bias from a major imbalance in baseline characteristics and performance bias. Another bias is from lack of comparability in clinical experience, implementation details of the intervention and compliance with the intervention.

#### Data synthesis and statistical methods

We constructed network plots using Stata software (StataCorp. V.2017. Stata Statistical Software: 118 Release 15. College Station, Texas: StataCorp LLC) to visualize all head-to-head comparisons for all outcomes.^21^ For primary and secondary outcomes, we presented all treatment comparisons using a network graph separately. YJ Xie appraised the clinical homogeneity before the start of the analysis, by tabulating study and population characteristics and inspecting them for differences in potential effect modifiers. This informed assessment of the assumption of exchangeability required for NMA. Treatments were also assigned to categories (ie, classes). For our primary outcome measures, the worst pain in the past week was expressed as MDs, with 95% CrIs. For our secondary outcome measures, the functional ability of knee function in activities was also expressed as MDs, with 95% CrIs.

#### Measures of treatment effect

We calculated mean differences (MDs) with 95% confidence intervals (CIs) for continuous outcomes as appropriate. When two or more studies presented their data derived from the same instrument of evaluation (with the same units of measurement), we pooled data as a mean difference. Conversely, we used the standardized mean difference (SMD) when primary studies express the same variables through clearly different instruments (and different units of measurement). In the case of pooling of different units of measurements, we scaled values to 0 to 10 (lower is better) for pain and 0 to 100 (higher is better) for functional ability. To re-express SMDs in VAS (0 to 10) and AKPS (0 to 100), we multiplied SMDs and 95% CIs by an estimate (the median of all control and intervention standard deviations (SDs)) of the SD of VAS or AKPS respectively.

## Results

### Selection process

A total of 45 trials with 42,319 patients were included in the network meta-analysis. A flow diagram summarising the study selection process is shown in Figure 1. Online supplemental appendix 1 shows the excluded studies and the reasons for exclusion. Online supplemental appendix 2 and 3 also show studies awaiting classification and studies identified in trial registers.

**Fig1.**
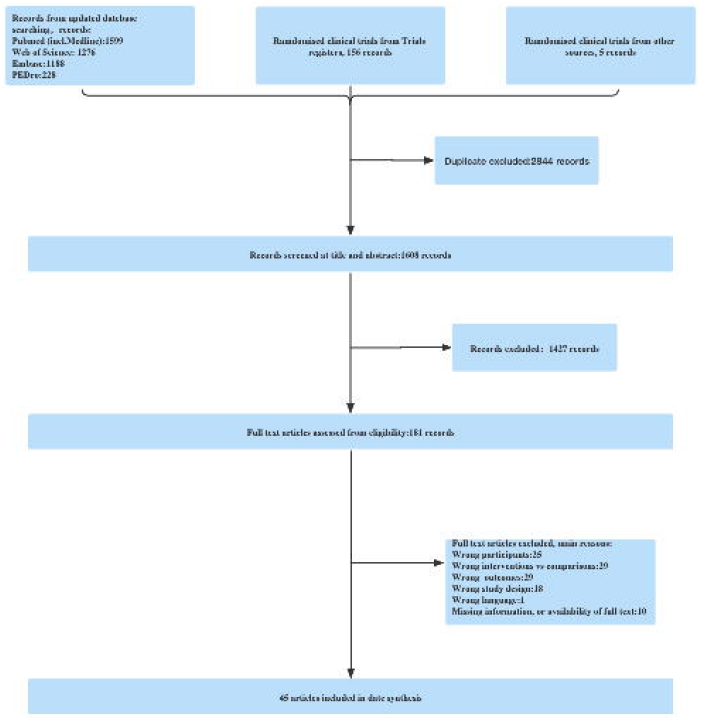
Study selection process. RCTs, randomized controlled trials.

### Characteristics of the studies included

Twenty-one treatments were investigated in forty-five trials. For full details see online supplemental appendix 4 and 5. Cumulatively, 42319 patients with PFP were included. Online supplemental appendix 5 details all studies and patient characteristics of the studies included.

### Assessment of risk of bias

The quality of the studies varied as shown in online supplementary appendix 6.

### Network meta-analyses

Figure 2 shows the network of eligible comparisons for the outcomes involving all trials. Twenty treatments were included in network analysis for the primary outcome, and fourteen treatments were included in network analysis for the secondary outcome. Descriptive details of the included treatments and resulting networks are provided in figure 2 and online supplemental appendix 4.

**Fig2.**
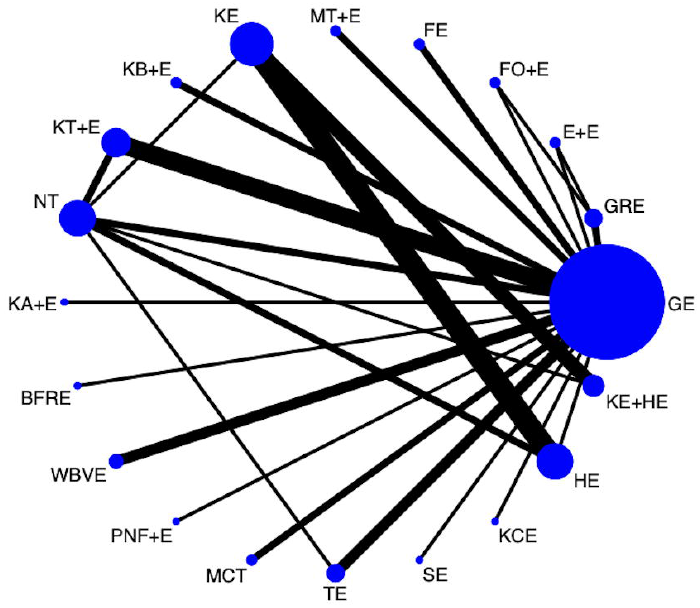
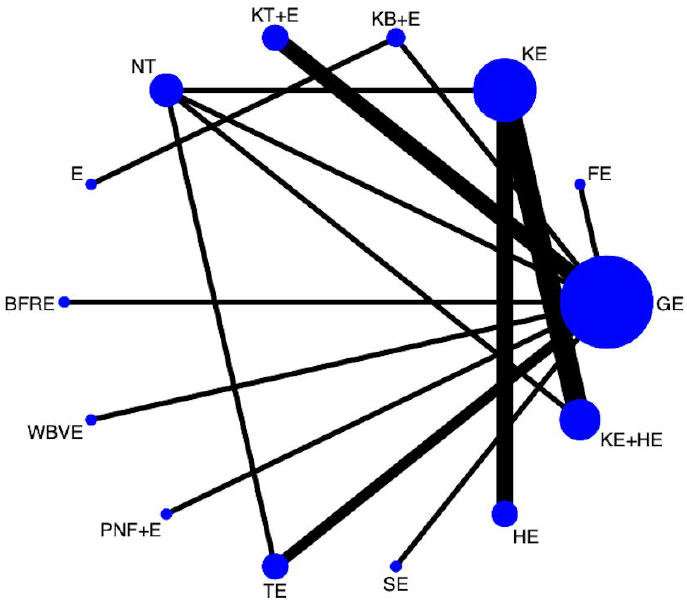
Network graphs for direct treatment comparisons for VAS(Figure2A) and AKPS(Figure2B). The thickness of the lines and the size of the dots are proportional to the number of trial comparisons and the number of participants in the treatment arms, respectively. GE=General Exercise; GRE=Gait Retraining Exercise; E+E=Education + Exercise; FO+E=Foot Orthoses + Exercise; FE=Feedback Exercise; MT+E=Manipulation Treatment +Exercise; KE=Keen Exercise; KB+E=Knee Brace + Exercise; KT+E=KinesioTaping + Exercise; NT=Nontreatment; KA+E=Knee Arthroscopy + Exercise; E=Education; BFRE=Blood Flow Restriction Exercise; WBVE=Whole Body Vibration Exercise; PNF+E=Proprioceptive Neuromuscular Facilitation + Exercise; MCT=Motor Control Training; TE=Target Exercise; SE=Supervised Exercise; KCE= Kinetic Chain Exercise; HE=Hip Exercise; KE+HE=Knee Exercise + Hip Exercise

### Comparative treatments effectiveness on the outcomes

A total of 45 trials with 42,319 patients were included in this network meta-analysis (NMA).

#### Pain relief

In terms of pain relief, all included treatments were superior to a wait-and-see approach: PNF + exercise (SMD -2.88, 95%CIs -4.75 to -1.02), whole body exercise (-1.57, -3.15 to -0.00), hip-and knee-focused exercise therapy (-1.32, -2.57 to -0.06), foot orthoses + exercise (-1.06, -2.92 to -0.06), hip exercise (-1.10, -2.44 to 0.24), knee brace + exercise(-0.91, -2.54 to 0.72), supervised exercise (-0.93, -2.72 to 0.86), gait retraining exercise (-2.55, -4.72 to -0.37), knee exercise (-0.92, -2.16 to 0.33), blood flow restriction exercise (-0.76, -2.55 to 1.02), knee arthroscopy + exercise (-0.61, -2.44 to 1.22), target exercise (-0.52, -2.38 to 1.33), kinesiotaping + exercise (-0.54, -2.07 to 0.99), manipulation treatment (-0.53, -2.21 to 1.15), education + exercise (-0.47, -2.31 to 1.38), motor control training (-0.40, -2.06 to 1.26), kinetic chain exercise (-0.27, -2.10 to 1.55), feedback exercise (-0.22, -1.86 to 1.43) (online supplemental appendix 8).

Exercise therapy with education (SMD -0.25, 95%CIs -1.76 to 1.26) was better than exercise alone in alleviating pain intensity.

#### Function improvement

Based on this NMA, all included treatments also showed a significant increase in function improvement compared with nontreatment: hip-and knee-focused exercise (SMD 1.40, 95%CIs 0.39 to 2.41), knee brace + exercise (1.31, -0.23 to 2.85), target exercise (1.27, 0.22 to 2.33), feedback exercise (1.30, 0.22 to 2.33), supervised exercise (1.27, -0.18 to 2.72), blood flow restriction exercise (1.17, -0.31 to 2.64), PNF + exercise (1.16, -0.41 to 2.72), kinesiotaping + exercise (1.14, -0.31 to 2.64), education (1.15, -0.73 to 3.03), whole body vibration (1.11, -0.45 to 2.68), general exercise (0.96, -0.10 to 2.01), knee exercise (0.66, -0.35 to 1.67), hip exercise (0.57, -0.61 to 1.76) (online supplemental appendix 8).

Education (SMD 0.58, 95%CIs -1.65 to 2.80) was superior to exercise in knee function improvement (online supplemental appendix 8).

#### Treatment ranking(figure 3)

The surface under the cumulative ranking curve (SUCRA) also supported the NMA results by showing the best treatment as the hip and knee-focused exercise for pain relief (69.6%) and function improvement (69.6%) (figure 3A and 3B). The exercise with PNF had the SUCRA 94.8% on the outcome of pain intensity (figure 3A). Exercise with the biomechanical device was better than exercise alone for pain relief (exercise with foot orthoses 62.6%; exercise with knee brace 59.3%; exercise with kinesiotaping 40.5%) and function improvement (exercise with knee brace 63.2%; exercise with kinesiotaping 54.3%) (figure 3A and 3B).

**Fig3.**
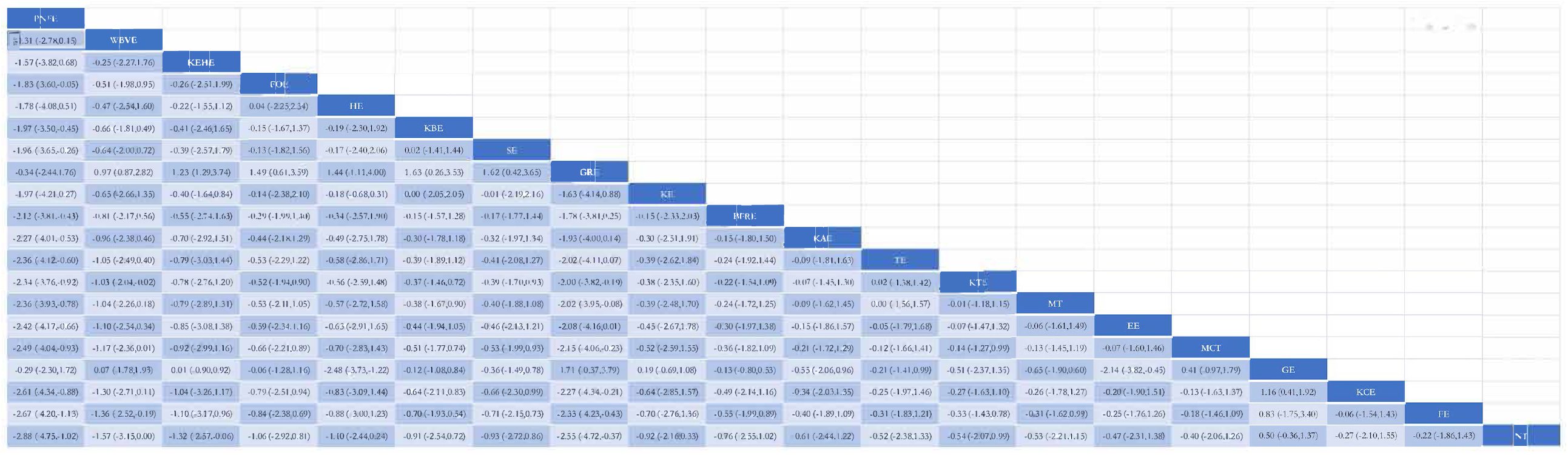
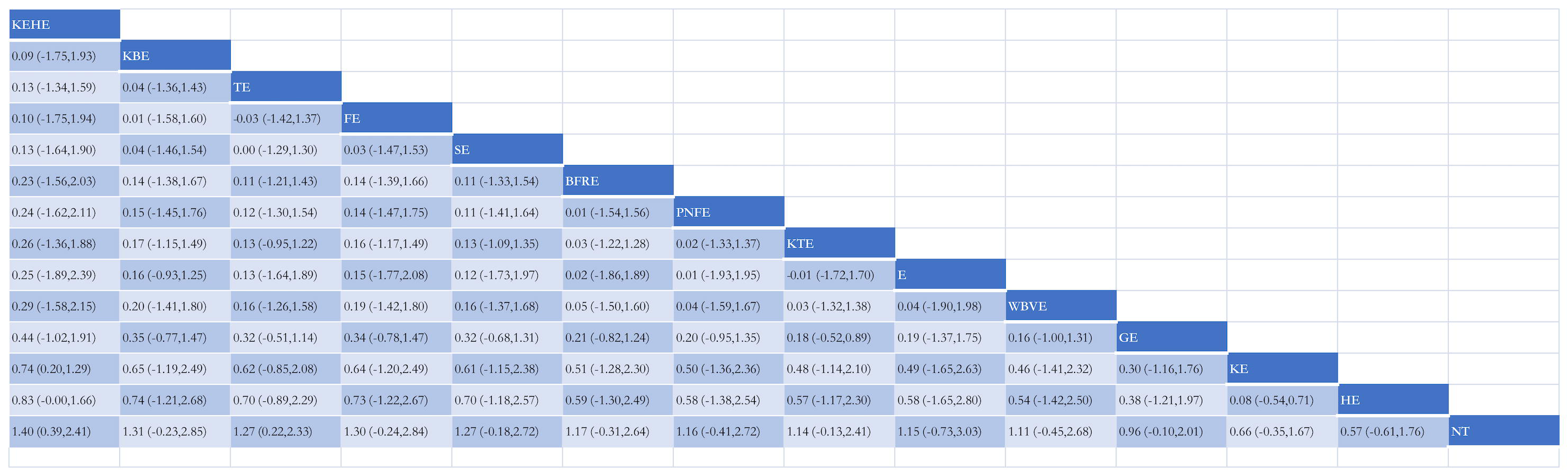
Ranking of treatment strategies based on probability of their protective effects on outcomes of pain relief (Figure3A)) and function improvement (Figure3B) according to the cumulative ranking area (SUCRA). Larger probability, stronger protective effects. GE=General Exercise; GRE=Gait Retraining Exercise; E+E=Education + Exercise; FO+E=Foot Orthoses + Exercise; FE=Feedback Exercise; MT+E=Manipulation Treatment +Exercise; KE=Keen Exercise; KB+E=Knee Brace + Exercise; KT+E=KinesioTaping + Exercise; NT=Nontreatment; KA+E=Knee Arthroscopy + Exercise; E=Education; BFRE=Blood Flow Restriction Exercise; WBVE=Whole Body Vibration Exercise; PNF+E=Proprioceptive Neuromuscular Facilitation + Exercise; MCT=Motor Control Training; TE=Target Exercise; SE=Supervised Exercise; KCE=Kinetic Chain Exercise; HE=Hip Exercise; KE+HE=Knee Exercise + Hip Exercise

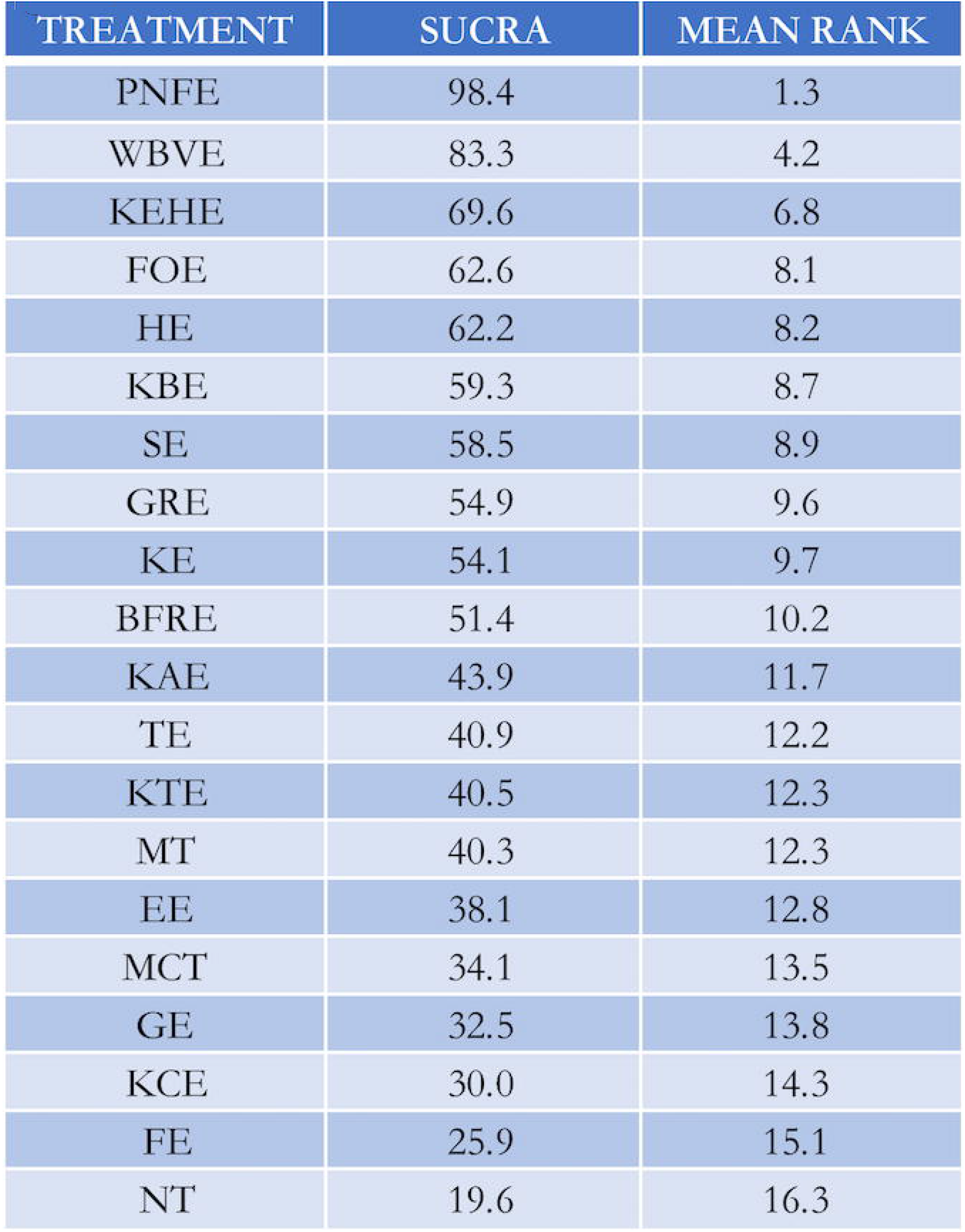
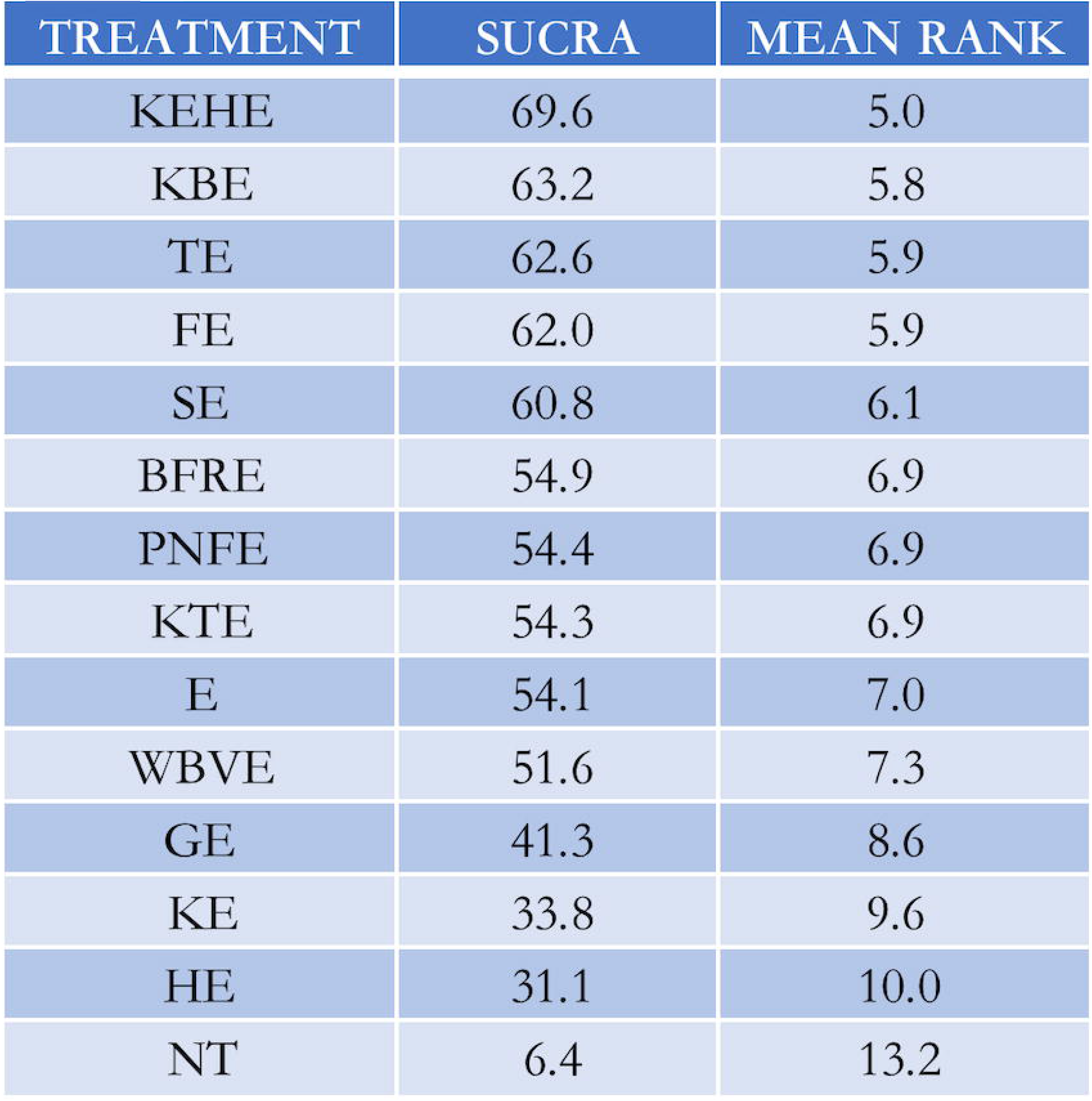

Target exercise (SUCRA 62.6%) displayed a better effect on knee function (figure 3B).

## Discussion

This NMA including 45 RCTs with a total of 42319 patients evaluated the effects of exercise therapy on pain and function in individuals with PFP. Based on this NMA, we have several new findings.

Firstly, the hip-and-knee combination muscle strength training is better than the general exercise therapy in knee function improvement and pain relief. General exercise is referred to as resistance exercise to improve muscle strength, endurance, or mass. The way of the general exercise could be delivered face to face or as a home exercise program (online supplemental appendix 4).^3, 5^ Although the multifactorial etiology of the PFP is not fully understood, muscle weakness is thought to be a major cause, which includes both local and non-local muscles.^2, 3, 5^ The local factor is related to the impaired quadriceps function, such as muscle size, muscle force, and vastus mediales obliques muscle reflex response time.^3, 22^ The non-local factor is related to proximal muscle dysfunction specifically in the abductors and external rotators, including lower gluteus minimus and medius peak muscle force, and lower hip abductor strength.^3, 23, 24^ The hip-and-knee combination exercise program could maintain the biodynamics of the patellofemoral joint (PFJ) by improving proximal and distal muscle strength.^5, 25-34^ So, compared with the general exercise, the hip-and-knee combination strengthening exercises can comprise exercises for both local and non-local muscle weakness.

Secondly, exercise therapy combined with the biomechanical device is better than general exercise for pain reduction and functional improvement.Individuals with PFP demonstrate abnormal biomechanics, including patellofemoral joint alignment and trochlear morphology.^3, 10, 35^ Carlson found that the distance between the tibial tubercle and the tibial chute was > 15 mm in 30% of PFP participants by magnetic resonance imaging.^36^ Excessive lateral displacement of the tibial tubercle causes the patella to be pulled laterally during knee flexion.^37-40^

Mal-alignment may lead to a deviant tracking pattern of the patella to the femoral groove and abnormal distribution of joint reaction stress on the patella subchondral bone.^41^ Moreover, the insufficient stabilization of the pelvis and the femoropatellar joint could increase the dynamic Q angle and further influence PFJ kinematics. ^24, 37-42^ Altered mechanics and increased PFJ stress are thought to be involved in the progression of PFJ osteoarthritis (OA).^5^ The biomechanical device could help to reduce PFJ stress and limit structural damage by altering the patella position in the trochlea.^14, 40^ Foot orthoses can support the media arch and prevent excessive pronation of the foot. The orthoses are meant to alter patella position in the trochlea by supporting patella tracking and redistributing PFJ stress.^43, 44^ Knee braces can promote a reduction in anterior pelvic tilt (APT), decrease femoral internal rotation, and ease the lateral pull on the patella.^45-47^ The braces can increase hamstring activity and inhibit the hip flexor muscle by increasing pelvic stability and optimizing the position of the patella.^46^ Kinesiotaping allows a partial to the full range of motion for the applied muscles and joints with different pulling forces to the skin.^48^ The tapings can lift the skin and increase the spaces between the skin and muscle, hence reducing the localized pressure and helping to promote circulation and lymphatic drainage.^48-51^ This NMA found that exercise therapy with biomechanical devices has a better effect in altering biomechanical dysfunction than routine training alone.Therefore, we recommend using biomechanical devices after individual evaluation.

Thirdly, education with exercise therapy shows a great effect on pain relief and function improvement than exercise alone.

There is a high recurrence rate in PFP patients.^4^ Chronic pain may have a significant impact on quality of life and burden of life, such as loss of physical function, loss of self-identity, pain-related confusion, and fear.^9^ Patient education can enhance patient compliance and adherence to self-management and help the sufferers overcome kinesophobia.^45, 52^ Patient education is less likely to have adverse effects. Clinicians can advocate the importance of active participation in exercise therapy and explain basic knowledge. Moreover, clinicians can provide targeted health education to patients, such as load management and appropriate body quality management. Future trials can explore the most effective education method balancing the cost and scalable intervention.

Finally, all treatments show a better effect on pain relief and function improvement than a wait-and-see approach. Also, this NMA found exercise therapy combined with additional training showed advantages over routine exercise therapy alone.

Although guidelines for musculoskeletal pain often recommend a wait-and-see approach in general practice.^13^ This NMA found the wait-and-see approach is the least effective treatment available. The etiology of PFP appears to be multi-factorial.^5^ Consequently, the interventions utilized to address the factorial changes in kinesiology or biomechanics are inevitable. We recommend avoiding a wait-and-see approach. This NMA found a multi-modal approach revealed a better clinical effect, such as gait retraining, feedback training, and WBV training. According to SUCRA, gait retraining (SUCRA 54.9%) is better than general exercise in pain relief. The gait retraining can tender evaluation of step rate and strike pattern manipulation, strategies to alter proximal kinematics and cues to reduce impact loading variables.^53^ Compared with general exercise, gait retraining can alter a running technique to treat lower limb injuries by reducing the load in certain muscle groups and joints.^43, 54, 55^ There was a consensus that the use of real-time feedback is effective in reducing variables related to ground reaction forces and modifying identified risky lower extremity kinematic movement patterns.^56^ The real-time feedback can supplement a rehabilitation program on perceived pain, patellar tracking, and isokinetic knee extension strength.^57^ Despite biofeedback exercise (SUCRA 25.9%) did not alleviate pain significantly, it can improve knee function (SUCRA 62%) based on this NMA. This finding may give clinicians a new way to improve knee function by applying biofeedback. One of the studies included in this NMA^58^ demonstrated that WBV training can improve pain intensity effectively. Compared with general exercise, WBV training shows a better effect on pain intensity (SUCRA 83.3%) and knee function (SUCRA 51.6%). The tonic vibration reflex can increase the recruitment of the motor units and the activity of the proprioceptive system by the mechanical platform vibrations.^59, 60^ Additionally, WBV exercise can increase blood flow and reduce muscle stiffness after vibration resulting in a clinical improvement.^59-61^ Combination therapy can provide additional clinical benefits compared to general exercise therapy. Clinicians may consider a multi-modal approach in the management of PFP when optimal management of PFP remains unclear.

### Strengths and weaknesses

Previous studies on the management of PFP were restricted to traditional comparisons of one treatment versus another or comparisons of several categories. Consistent with the findings of a previously published review,^13^ we found any treatment, even if education only, is superior to a wait-and-see approach. The hip-and-knee combination muscle training is most effective for pain relief and functional improvement. Besides, comprehensive exercise therapy, (such as exercise with taping, and education) has a better effect than exercise only. This NMA will be updated when new evidence becomes available, ensuring a contemporary overview of the evidence for the best treatment.

Limitations in the conclusions drawn by the NMA are primarily caused by the original data. First, to analyze the efficacy of exercise therapy as best as possible, we did not limit the duration of follow-up, which resulted in no further analysis of the long-term and short-term effects of the included interventions. Second, only two trials included more than 100 participants per group, which may introduce bias due to small study effects. Third, data were pooled from the study duration. Because the duration of the studies was not entirely consistent, assessments of pain and function have not yet completely stabilized. Fourth, the SUCRA curve was used to estimate a ranking probability of comparative effectiveness between the different therapies, but it has limitations and the results should be interpreted with caution.

The need for an individual exercise therapy approach to PFP treatment. This NMA allows clinicians to devise an individual prescription. Personalized exercise prescriptions should be further studied by clinicians or therapists in the future (online online supplemental appendix 9). Larger RCTs are needed to resolve the uncertainty around the efficacy of exercise therapy for PFP.

## Conclusion

The knee and hip combination strength training is highly effective in muscle strength improvement. All treatments in our NMA were superior to nontreatment, we recommend avoiding a wait-and-see approach. Comprehensive therapy based on individual evaluation can effectively improve the symptoms of patients.

### Web extra

Extra materials supplied by authors.

### Note

## Data Availability

All data produced in the present study are available upon reasonable request to the authors.

## Contributors

YJX, YXJ, and CZ came up with the study idea.

YJX, YXJ, CZ designed the study.

YJX, YXJ, and CZ designed the statistical analyses plan.

YJX constructed the search with help of a research librarian.

YJX and YXJ performed the search and selection process.

XZZ, SW and AM performed data extraction.

XZZ, XZ, and LL performed the risk of bias assessments.

YJX performed all data analyses.

YJX, YXJ, and LL performed Grading of Recommendations, Assessment, Development, and Evaluation assessments.

YJX, YXJ, and CZ drafted the first version of the manuscript. YJX, YXJ, and CZ are the study guarantors.

All authors provided feedback and gave important intellectual input. All authors read and consented to the content of the article.

## Provenance and peer review

Not commissioned; externally peer reviewed.

