## Supplemental materials for "Comparative efficacy of exercise therapy for patellofemoral pain: A network meta-analysis of randomized controlled trials"

*Co-first author: Yu-Jie Xie

**Abstract:**

**Objective**: To investigate the effective exercise prescription in randomized controlled trials (RCTs) for patellofemoral pain (PFP).

**Design**: A network meta-analysis.

**Data sources**: PubMed (including Medline), Embase, Web of Science, PEDro, Clinicaltrials.gov and other resourses for RCTs.

**Eligibility criteria for selecting studies**: RCTs of exercise interventions for PFP with outcomes of pain intensity or functional improvement.

**Primary outcome measure**: Pain intensity is measured by 'worst pain in the past week' on a Visual Analogue Scale (VAS) or Numerical Rating Pain Scale (NRS).

**Data extraction:**

Two researchers independently extracted data and assessed the bias of risks. We used Grading of Recommendations, Assessment, Development, and Evaluation to appraise the strength of the evidence.

**Results:**

A total of 45 trials with 42,319 patients were included in this network meta-analysis (NMA). For the primary outcomes, all included treatments were superior to a wait-and-see approach: PNF + exercise (SMD -2.88, 95%CIs -4.75 to -1.02), whole body exercise (-1.57, -3.15 to -0.00), hip-and knee-focused exercise therapy (-1.32, -2.57 to -0.06), foot orthoses + exercise (-1.06, -2.92 to -0.06), hip exercise (-1.10, -2.44 to 0.24), knee brace + exercise(-0.91, -2.54 to 0.72), gait retraining exercise (-2.55, -4.72 to -0.37), knee exercise (-0.92, -2.16 to 0.33), knee arthroscopy +

exercise (-0.61, -2.44 to 1.22), target exercise (-0.52, -2.38 to 1.33), kinesiotaping + exercise (-0.54, -2.07 to 0.99), education + exercise (-0.47, -2.31 to 1.38), feedback exercise (-0.22, -1.86 to 1.43). Exercise therapy with education (SMD -0.25, 95%CIs -1.76 to 1.26) was better than exercise alone in alleviating pain intensity.

**Conclusion**:

The knee and hip combination strength training is highly effective in muscle strength improvement. All treatments in our NMA were superior to nontreatment, we recommend avoiding a wait-and-see approach. Comprehensive therapy based on individual evaluation can effectively improve the symptoms of patients.

**Keywords:** biomechanical phenomena; knee; patellofemoral pain; exercise; osteoarthritis;

**Web appendices**

Contents

Web appendix1 Excluded studies with reasons for exclusions, including references to excluded studies

Web appendix2 Studies awaiting classification

Web appendix3 Studies identified in trial registers

Web appendix4 Treatments and treatment categories (ie, classes)

Web appendix5 Characteristics included studies, interventions and outcomes

Web appendix6 Risk of bias judgements per study

Web appendix7 Certainty of the Evidence (GRADE approach)

Web appendix8 Data analyses, treatment level results

Web appendix9 A recommended exercise therapy

### WEB APPENDIX1. EXCLUDED STUDIES AND REASONS FOR EXCLUSION

| Study identifier | Reason(s) for exclusion |
|---|---|
| Lee, J.H., et al 2021 | Wrong intervention: compares static and dynamic quadriceps stretching exercises |
| Lee, J.H., et al 2021 | Wrong intervention: compare static and dynamic hamstring stretching |
| Almeida, G.P.L., et al 2021 | Wrong intervention: compare anteromedial versus posterolateral hip musculature strengthening |
| Abd-Elmonem, A.M., et al 2021 | Wrong study populations: 72 children, seven to twelve years old |
| Zarei, H., et al 2020 | Wrong intervention: exercise therapy or exercise-therapy+dry needling group |
| Yosmaoğlu, H.B., et al 2020 | Wrong intervention: received standard multimodal treatment |
| Wang, B., et al 2020 | Wrong outcomes: changes from baseline in the patellofemoral joint stress and contact force. |
| Tan, J.M., et al 2020 | Wrong study populations: Twenty-one participants with PFOA |
| Tan, J.M., et al 2020 | Wrong study design: cross-sectional study<br>Wrong study populations: 179 individuals aged 50 years and diagnosed PFOA |
| Tan, J.M., et al 2020 | Wrong study design: cross-sectional study<br>Wrong study populations: 188 participants with symptomatic PFOA |
| Talbot, L.A., et al 2020 | Wrong intervention: neuromuscular electrical stimulation |
| O'Sullivan, I.C., et al 2020 | Wrong study design: Protocol |
| Matthews, M., et al 2020 | Wrong study design: A two-arm parallel, multicentre randomised superiority clinical trial |
| Marshall, A.N., et al 2020 | Wrong outcomes: triplanar kinematics at the trunk, hip, knee, and ankle were collected via 3-dimensional motion capture. |
| Ma, Y.T., et al 2020 | Wrong intervention: trigger point dry needling |
| Letafatkar, A., et al 2020 | Wrong outcomes: Kinematic and kinetic measurements |
| Hott, A., et al 2020 | Wrong study design: Cohort study |
| Glaviano, N.R., et al 2020 | Wrong intervention: patterned electrical neuromuscular stimulation |
| Celik, D., et al 2020 | Wrong intervention: Neuromuscular electrical stimulation |
| Albornoz-Cabello, M., et al 2020 | Wrong intervention: Monopolar dielectric diathermy by emission of radiofrequency |
| Straszek, C.L., et al 2019 | Wrong study design: A randomized crossover study |
| Smith, B.E., et al 2019 | Wrong study purpose: to explore potential barriers and facilitators with patients and physiotherapists with patellofemoral pain |

| | |
|---|---|
| Smith, B.E., et al 2019 | Wrong intervention: a loaded self-managed exercise programme (n =30) or usual physiotherapy (n=30). |
| Rees, D., et al 2019 | Wrong study purpose: make inferences about knee kinematics during running. |
| Prohorova, E. S., et al 2019 | Wrong language: Russian |
| Lewinson, R. T., et al 2019 | Wrong study purpose: To determine whether footwear insoles alter biomechanical variables associated with running injuries. |
| Glaviano, N.R., et al 2019 | Wrong intervention: Patterned electrical neuromuscular stimulation |
| Dos Santos, A.F., et al 2019 | Wrong intervention: gait retraining<br>Wrong study design: Case series report |
| Aliberti S PhD, P.T., et al 2019 | Wrong study purpose: To analyze the immediate effects of a distal gait modification session |
| Adel, J., et al 2019 | Wrong study populations: participants with PFOA |
| Wyndow, N., et al 2019 | Wrong study populations: Fifty-one participants with PFOA |
| Sit, R.W.S., et al 2018 | Wrong study populations: 208 patients with knee osteoarthritis |
| Sit, R.W.S., et al 2018 | Wrong study populations: 208 participants with coexistence of PFOA and TFOA |
| Ramskov, D., et al 2018 | Wrong study populations: healthy recreational runners |
| Mulvad, B., et al 2018 | Wrong study purpose: to describe the incidence proportion of different types of running-related injuries (RRI) |
| Esculier, J.F., et al 2018 | Wrong study design: Secondary analyses |
| Earl-Boehm, J.E., et al 2018 | Wrong study purpose: To develop clinical prediction rules |
| Bonanno, D.R., et al 2018 | Wrong study purpose: To evaluate the effectiveness of prefabricated foot orthoses for the prevention of lower limb overuse injuries. |
| Son, S.J., et al 2017 | Wrong intervention: transcutaneous electrical nerve stimulation |
| Roper, J.L., et al 2017 | Wrong study purpose: investigated the effects of gait retraining from rearfoot strike to forefoot strike |
| Matthews, M., et al 2017 | Wrong study design: protocol |
| Lee, J., et al 2017 | Wrong outcomes: Surface electromyography activities in the VMO, VL, and AbdH |
| Greuel, H., et al 2017 | Wrong study populations: 22 healthy participants |
| Espí-López, G.V., et al 2017 | Wrong intervention: trigger point dry needling |
| Cotic, M., et al 2017 | Wrong study purpose: isolated resurfacing of the trochlea using an inlay prosthesis without changing the complex kinematics of the patellofemoral joint. |
| Collins, N.J., et al 2017 | Wrong study design: A cross-over, proof-of-concept study |
| Bazett-Jones, D.M., et al 2017 | Wrong study design: Crossover study |
| Servodio Iammarrone, C., et al 2016 | Wrong intervention: pulsed electromagnetic fields |

| | |
|---|---|
| Riel, H., et al 2016 | Wrong outcomes: the mean deviation of the prescribed time under tension per repetition in seconds |
| Rathleff, M.S., et al 2016 | Wrong outcomes: sEMG of the VL and VM during stair descent |
| Rathleff, M.S., et al 2016 | Wrong outcomes: the change in localized and distal from baseline to follow-up |
| Petersen, W., et al 2016 | Wrong intervention: a medially directed patellar realignment brace and supervised exercise |
| Keays, S. L., et al 2016 | Wrong study design: prospective cohort study |
| Glaviano, N. R., et al 2016 | Wrong intervention: patterned electrical neuromuscular stimulation<br>Wrong study design: Cohort study |
| Glaviano, N.R., et al 2016 | Wrong intervention: patterned electrical neuromuscular stimulation |
| Esculier, J.F., et al 2016 | Wrong intervention: Group 1: education; Group 2 : education +an exercise program ; Group 3: education +running gait retraining advice |
| Araújo, C.G., et al 2016 | Wrong outcomes: the level of muscle activity of each muscle |
| Tsai, L.C., et al 2015 | Wrong intervention: lower extremity off-axis training |
| Strecker, W., et al 2015 | Wrong study populations: Patellofemoral maltracking |
| Palmer, K., et al 2015 | Wrong outcomes: Knee kinematics |
| Lewinson, R.T., et al 2015 | Wrong intervention: an experimental 3 mm lateral wedge or control 6 mm medial wedge group |
| Lankhorst, N.E., et al 2015 | Wrong study design: Secondary exploratory analysis |
| Keays, S. L., et al 2015 | Wrong study design: crossed study |
| Hott, A., et al 2015 | Wrong study design: protocol |
| Crossley, K.M., et al 2015 | Wrong study populations: 92 people aged ≥40 years with symptomatic and radiographic PFJ OA participated. |
| Baldon Rde, M., et al 2015 | Wrong outcomes: the lower limb and trunk kinematics in the frontal plane assessed during a single-legged squat task |
| Sugimoto, D., et al 2014 | Wrong study populations: 21 high school women volleyball players |
| Rabelo, N.D., et al 2014 | Wrong study design: protocol |
| Petersen, W., et al 2014 | Wrong study design: Pro study |
| Knoop, J., et al 2014 | Wrong study populations: Ninety-five participants with knee OA |
| Khayambashi, K., et al 2014 | Wrong study design: Comparative control trial |

| | |
|---|---|
| Bonacci, J., et al 2014 | Wrong study populations: 22 trained runners |
| Baldon Rde, M., et al 2014 | Wrong outcomes: trunk endurance and eccentric hip and knee muscle strength |
| Toumi, H., et al 2013 | Wrong study purpose: To investigate the extent to which quadriceps muscle activation and strength are responsible for patellofemoral pain. |
| Rodrigues, P., et al 2013 | Wrong study populations: Kinematics of 16 asymptomatic and 17 runners with AKP |
| Pattyn, E., et al 2013 | Wrong study purpose: to examine by muscle functional magnetic resonance imaging |
| Østerås, B., et al 2013 | Wrong intervention: high-dose, high-repetition medical exercise therapy, and the control group received low-dose, low-repetition exercise therapy. |
| Østerås, B., et al 2013 | Wrong intervention: high-dose, high-repetition MET for the experimental group, and low-dose, low-repetition exercise therapy for the control group. |
| Osorio, J.A., et al 2013 | Wrong study design: Crossover experimental design. |
| Lewinson, R. T., et al 2013 | Wrong study populations: Nine healthy runners participated |
| Boldt, A.R., et al 2013 | Wrong study populations: Twenty female runners with and without PFPS participated |
| Rathleff, M.S., et al 2012 | Wrong outcomes: fill out self-reported questionnaires 、 a 7-point Likert scale |
| Pattyn, E., et al 2012 | Wrong study purpose: aims to identify factors that could predict the short-term functional outcome |
| Kettunen, J.A., et al 2012 | Wrong study design: 5-year follow-up，secondary study |
| Willy, R. W., et al 2011 | Wrong outcomes: Using a handheld dynamometer and standard motion capture procedures |
| Shih, Y. F., et al 2011 | Wrong study populations: pronated-foot runners with overuse knee or foot pain |
| Mostamand, J., et al 2011 | Wrong outcomes: using an EMG unit |
| Tan, S.S., et al 2010 | Wrong study purpose: determine the cost effectiveness of exercise therapy |
| MacLean, C. L., et al 2010 | Wrong study populations: injured runners |
| Irish, S.E., et al 2010 | Wrong outcomes: The EMG of VMO and VL was measured and used to calculate the VMO:VL ratio |
| Balci, P., et al 2009 | Wrong outcomes: the MRFS System、a visual analog scale、the Kujala questionnaire |
| Selfe, J., et al 2008 | Wrong study purpose: investigated the effect of patellar bracing and taping on the three-dimensional mechanics of the knee during a controlled eccentric step down task. |

| | |
|---|---|
| Ng, G. Y., et al 2008 | Wrong outcomes: VMO/VL EMG ratio |
| Crossley, K.M., et al 2008 | Wrong study design: protocol<br>Wrong study populations: 90 people with PFJ OA |
| Bily, W., et al 2008 | Wrong study design: a pilot study |
| Bakhtiary, A. H., et al 2008 | Wrong study populations: 32 female university students with a diagnosis of patellar chondromalacia |
| McCrory, J.L., et al 2007 | Wrong outcomes: Three-dimensional kinematic data were collected for each subject at 60 Hz |
| Herrington, L., et al 2007 | Wrong intervention: Group 1 performed knee extension exercises, group 2 performed seated leg press exercises, and group 3 received no treatment. |
| Avraham, F., et al 2007 | Wrong study design: a pilot study |
| van Linschoten, R., et al 2006 | Wrong study design: The PEX study |
| Singer, B.J., et al 2006 | Wrong study purpose: An open label pilot investigation of the efficacy of Botulinum toxin type A |
| Cowan, S.M., et al 2006 | Wrong outcomes: the EMG amplitude of the vastus medialis obliquus and vastus lateralis during the concentric phase of stair stepping. |
| Boling, M.C., et al 2006 | Wrong study populations: Fourteen subjects diagnosed with PFPS and 14 healthy control subjects |
| Bennell, K., et al 2006 | Wrong outcomes: EMG onsets of VMO and VL、stance phase knee flexion and vertical ground reaction force |
| O'Sullivan, S. P., et al 2005 | Wrong outcomes: maximum VMO activity |
| Macintyre, J. G. 2005 | Wrong study design: secondary study |
| Hazneci, B., et al 2005 | Wrong outcomes: knee passive joint position sense, quadriceps and hamstring muscle strength |
| Denton, J., et al 2005 | Wrong outcomes: measurements of hip internal and external rotation, hip extension, and iliotibial band muscle length |
| Crossley, K.M., et al 2005 | Wrong outcomes: Stance-phase knee flexion |
| Coqueiro, K.R., et al 2005 | Wrong outcomes: electromyographic muscle data |
| Ward, S. R., et al 2004 | Wrong study purpose: To determine if persons with patella alta exhibit elevated patellofemoral joint stress compared to pain-free controls during normal and fast walking speeds. |
| Van Tiggelen, D., et al 2004 | Wrong study populations: 167 military recruits without history of knee pain |
| Quilty, B., et al 2003 | Wrong study populations: who had knee pain and predominant PFJ OA |
| Sathe, V.M., et al 2002 | Wrong study design: an MRI study |

| | |
|---|---|
| Ng, G. Y., et al 2002 | Wrong study design: Pre- and post-treatment design |
| Cowan, S.M., et al 2002 | Wrong outcomes: electromyographic (EMG) activity of the vasti |
| Schneider, F., et al 2001 | Wrong study purpose: alternatives for cases of therapy resistance |
| Lam, P. L., et al 2001 | Wrong outcomes: The ratio of surface-integrated electromyographic signals of vastus medialis obliquus over vastus lateralis |
| Witvrouw, E., et al 2000 | Wrong intervention: only closed kinetic chain exercises or only open kinetic chain exercises |
| Callaghan, M.J., et al 2000 | Wrong study populations: twenty healthy volunteers |
| Timm, K.E. 1998 | Wrong study purpose: examined the effects of a high volume of submaximal knee muscle exercise on objective measures of PFP and PFC |
| Laprade, J., et al 1998 | Wrong study populations: eight PFPS female subjects and 19 controls |
| Thomeé, R. 1997 | Wrong outcomes: torque measurements |
| Bynum, E. B., et al 1995 | Wrong intervention: closed kinetic chain exercises or open kinetic chain exercises |
| Werner, S., et al 1993 | Wrong outcomes: Borg's pain scale |
| Kannus, P., et al 1992 | Wrong intervention: intraarticular injections |

References to excluded studies in web appendix 2[1-126]

108. Denton, J., et al., The addition of the Protonics brace system to a rehabilitation protocol to address patellofemoral joint syndrome. J Orthop Sports Phys Ther, 2005. 35(4): p. 210-9.
109. Crossley, K.M., et al., Physical therapy improves knee flexion during stair ambulation in patellofemoral pain. Med Sci Sports Exerc, 2005. 37(2): p. 176-83.
110. Coqueiro, K.R., et al., Analysis on the activation of the VMO and VLL muscles during semisquat exercises with and without hip adduction in individuals with patellofemoral pain syndrome. J Electromyogr Kinesiol, 2005. 15(6): p. 596-603.
111. Ward, S.R. and C.M. Powers, The influence of patella alta on patellofemoral joint stress during normal and fast walking. Clin Biomech (Bristol, Avon), 2004. 19(10): p. 1040-7.
112. Van Tiggelen, D., et al., Effect of bracing on the prevention of anterior knee pain--a prospective randomized study. Knee Surg Sports Traumatol Arthrosc, 2004. 12(5): p. 434-9.
113. Quilty, B., et al., Physiotherapy, including quadriceps exercises and patellar taping, for knee osteoarthritis with predominant patello-femoral joint involvement: randomized controlled trial. J Rheumatol, 2003. 30(6): p. 1311-7.
114. Sathe, V.M., et al., Acute effects of the Protonics system on patellofemoral alignment: an MRI study. Knee Surg Sports Traumatol Arthrosc, 2002. 10(1): p. 44-8.
115. Ng, G.Y. and J.M. Cheng, The effects of patellar taping on pain and neuromuscular performance in subjects with patellofemoral pain syndrome. Clin Rehabil, 2002. 16(8): p. 821-7.
116. Cowan, S.M., et al., Physical therapy alters recruitment of the vasti in patellofemoral pain syndrome. Med Sci Sports Exerc, 2002. 34(12): p. 1879-85.
117. Schneider, F., K. Labs, and S. Wagner, Chronic patellofemoral pain syndrome: alternatives for cases of therapy resistance. Knee Surg Sports Traumatol Arthrosc, 2001. 9(5): p. 290-5.
118. Lam, P.L. and G.Y. Ng, Activation of the quadriceps muscle during semisquatting with different hip and knee positions in patients with anterior knee pain. Am J Phys Med Rehabil, 2001. 80(11): p. 804-8.
119. Witvrouw, E., et al., Open versus closed kinetic chain exercises for patellofemoral pain. A prospective, randomized study. Am J Sports Med, 2000. 28(5): p. 687-94.
120. Callaghan, M.J., et al., The reproducibility of multi-joint isokinetic and isometric assessments in a healthy and patient population. Clin Biomech (Bristol, Avon), 2000. 15(9): p. 678-83.
121. Timm, K.E., Randomized controlled trial of Protonics on patellar pain, position, and function. Med Sci Sports Exerc, 1998. 30(5): p. 665-70.
122. Laprade, J., E. Culham, and B. Brouwer, Comparison of five isometric exercises in the recruitment of the vastus medialis oblique in persons with and without patellofemoral pain syndrome. J Orthop Sports Phys Ther, 1998. 27(3): p. 197-204.
123. Thomeé, R., A comprehensive treatment approach for patellofemoral pain syndrome in young women. Phys Ther, 1997. 77(12): p. 1690-703.
124. Bynum, E.B., R.L. Barrack, and A.H. Alexander, Open versus closed chain kinetic exercises after anterior cruciate ligament reconstruction. A prospective randomized study. Am J Sports Med, 1995. 23(4): p. 401-6.
125. Werner, S. and E. Eriksson, Isokinetic quadriceps training in patients with patellofemoral pain syndrome. Knee Surg Sports Traumatol Arthrosc, 1993. 1(3-4): p. 162-8.
126. Kannus, P., et al., Effect of intraarticular glycosaminoglycan polysulfate treatment on. patellofemoral pain syndrome. A prospective, randomized double-blind trial comparing glycosaminoglycan polysulfate with placebo and quadriceps muscle exercises. Arthritis Rheum,

1992. 35(9): p. 1053-61.

#### Web appendix2. Studies found through database searches, awaiting classification

| *Study identifier* | *Comment* |
|---|---|
| Matthews, M., et al 2021 | Infographic. |
| Aghakeshizadeh, F., et al 2021 | No full text made available. |
| Zago, J., et al 2020 | No full text made available. |
| Begum, R., et al 2020 | No full text made available. |
| Foroughi, F., et al 2019 | Box plot, line chart. Lack of specific data. |
| Hamstra-Wright, K.L., et al 2017 | Box plot. Lack of specific data. |
| Yılmaz Yelvar, G.D., et al 2015 | No full text made available. |
| Ismail, M. M., et al 2013 | No full text made available. |
| Crossley, K., et al 2002 | Box plot, line chart. Lack of specific data. |
| Eng, J. J., et al 1993 | Line chart. Lack of specific data. |

References to excluded studies in web appendix 3

**WEB APPENDIX3. Potentially eligible trials from trial registers**

| | Trial identifier | Treatment comparison | Status | Comments |
|---|---|---|---|---|
| 1. | NCT03054701 | Hip specific resistance exercise VS Knee specific resistance exercise | Completed | No full text |
| 2. | NCT05261100 | Core stability exercises VS Conventional Physical Therapy | Recruiting | |
| 3. | NCT04340453 | Exercise with Blood Flow restriction training VS Hip and Knee Exercise Program VS stretching | Ongoing | |
| 4. | NCT04631614 | Warm up VS Passive calf muscle stretching VS Ankle joint mobilization - weight-bearing mobilization with movement (WB-MWM) technique | Not yet recruiting | |
| 5. | NCT05120583 | Pilates exercises VS Traditional physical therapy program | Ongoing | |
| 6. | NCT03985254 | Strength Training Group (STG) VS Strength and Power Training Group (SPTG) | Unknown | Study Protocol |
| 7. | NCT02827084 | Kinesio Taping VS Placebo | Unknown | Exclude, wrong outcomes, web appendix 2 |
| 8. | NCT03163290 | Posterolateral Hip Complex Exercises VS Anteromedial Hip Complex Exercises | Ongoing | |
| 9. | NCT04538508 | Diathermy VS Supervised knee exercises | Completed | |
| 10. | NCT02591680 | Neuromuscular training VS Muscle strengthening | Unknown | |
| 11. | NCT00496964 | Botulinum toxin A + exercise VS Placebo | Terminated | Exclude, wrong intervention, web appendix 2 |
| 12. | NCT03364855 | Star excursion balance test VS Kinesthetic Ability Trainer 00 VS SEBT and KAT 2000 | Unknown | |

| | | | | |
|---|---|---|---|---|
| 13. | NCT04011436 | Strengthening program | Completed | No full text |
| 14. | NCT02352909 | Education VS Exercise program VS Gait retraining | Completed | No full text |
| 15. | NCT02841384 | taping patellar McConnell VS Placebo taping | Completed | No full text |
| 16. | NCT04225000 | Exercise Group VS Mobilization Group | Completed | No full text |
| 17. | NCT05083897 | hip adduction isometric contraction | Not yet recruiting | |
| 18. | NCT04462718 | Radiofrecuency VS Therapeutic Exercise | Completed | |
| 19. | NCT03042559 | Protonics Knee brace VS Sports Cords | Completed<br>Has result | |
| 20. | NCT02597673 | Home Exercise Program (HEP) VS NMES VS TENS | Completed<br>Has results | |
| 21. | NCT02114294 | Isolated hip strengthening VS Quadriceps based training VS Active control | Active, not recruiting | |
| 22. | NCT02707679 | Mulligan's Straight leg-raise with traction VS Mulligan's Mobilization with Movement VS Kinesiotaping VS Exercise | Completed | |
| 23. | NCT02241148 | Acetaminophen 500mg VS Kinesio taping | Terminated | |
| 24. | NCT00445224 | Hip Progressive Resistive Exercise VS Quad Progressive Resistive Exercises | Completed<br>Has results | |
| 25. | NCT0197531 | Lumbopelvic Manipulation VS Passive lumbar spine flexion and extension | Completed | |
| 26. | NCT01691170 | Stretching Hamstring VS Quadriceps Strengthening | Completed | |
| 27. | NCT01504100 | | Completed | |
| 28. | NCT03324204 | Neuromuscular Training VS Extracorporeal Shock Wave Therapy (ESWT) | Unknown | |
| 29. | NCT00401050 | chiropractic manipulative therapy VS knee exercises VS | Completed | |

| | | | | |
|---|---|---|---|---|
| | | Graston nstrument Soft Tissue Mobilization (GISTM) | | |
| 30. | NCT01290705 | High exercise therapy VS Low exercise therapy | Completed | |
| 31. | NCT03069547 | Quadriceps Exercise program VS Hip Exercise program | Active, not recruiting | |
| 32. | NCT05125263 | kinesiotaping group VS mulligan taping group | Active, not recruiting | |
| 33. | NCT00451347 | Quadriceps strength training VS Taping VS Home exercise | Unknown | |
| 34. | NCT04748692 | Local exercise therapy VS Spinal manual therapy | ongoing | |
| 35. | NCT04975113 | progressive neuromuscular exercise program VS Exercise and Taping | ongoing | |
| 36. | NCT00736736 | Additional Hip Muscle Strengthening to Leg Press Exercise VS Leg Press Exercise | Completed | |
| 37. | NCT02123602 | core stabilization VS lower extremity training only | Unknown | |
| 38. | NCT02674841 | Feedback on TUT VS No feedback on TUT VS Exercise | Completed | Exclude, wrong study design, web appendix 2 |
| 39. | NCT04478422 | Muscle strengthening with vascular occlusion VS Conventional muscle strengthening | Not yet recruiting | |
| 40. | NCT00451438 | muscle activity of gluteus medius and tensor fascia lata during submaximal isometric muscle contraction | Completed | Exclude, wrong study design, “case-control”, web appendix 2 |
| 41. | NCT04086615 | NMES and exercise supplemented with high BFR VS NMES and exercise supplemented with low BFR | Recruiting | |
| 42. | NCT04480528 | Blood Flow Restriction Training (BFRT) VS Sham Blood Flow Restriction Training (Sham BFRT) | Completed | No full text |
| 43. | NCT02854774 | Muscle activation exercises | Terminated | No additional funding is available to continue this pilot study at this |

| | | | | |
|---|---|---|---|---|
| | | | | time |
| 44. | NCT03285464 | Hip strengthening program VS use the trunk as a lever to strengthen the hip | Completed | Exclude, wrong outcomes, web appendix 2 |
| 45. | NCT02624245 | Physical Therapy VS Movement control training | Completed | No full text |
| 46. | NCT00451087 | leg press training VS isokinetic training | Unknown | |
| 47. | NCT02837289 | Therapy taping VS Placebo taping | Completed | Exclude, wrong outcomes, web appendix 2 |
| 48. | NCT04264429 | Infrapatellar strap VS Elastic band | Recruitment | |
| 49. | NCT03101956 | Lumbar Spine Manipulation VS Lumbar Spine Manipulation Placebo | Completed | No full text |
| 50. | NCT03099512 | SFE VS Exercise | Completed | No full text |
| 51. | NCT01727596 | Taping | Completed | No full text |
| 52. | NCT04760158 | Taping group vs Sham Taping group vs exercise group | Completed | No full text |
| 53. | NCT04173468 | MWM Group vs Mulligan Taping Group | Ongoing | |
| 54. | NCT00662493 | Motor control retraining program VS Quadriceps strengthening program | Completed | No full text |
| 55. | NCT00348647 | physical training program to prevent injuries | Unknown | |
| 56. | NCT02322515 | Intervention taping VS Placebo Taping | Completed | No full text |
| 57. | NCT03117205 | Application of Kinesio Taping® VS Kinesio Taping® placebo application | Completed | No full text |
| 58. | NCT03771495 | Joint Mobilization VS Laying on of Hands | Completed | Exclude, wrong outcomes, web appendix 2 |
| 59. | NCT02333617 | Dry Needling VS Soft Tissue Mobilization VS Placebo Control VS Hip and core strengthening exercises | Terminated | |

| | | | | |
|---|---|---|---|---|
| 60. | NCT05168332 | EMG-BF guided patellar taping VS sham patellar taping without EMG-BF guided stimulation; maximum voluntary isometric contraction exercise | Recruiting | |
| 61. | NCT04031248 | Whole body vibration VS EXERCISE group | Completed | No full text |
| 62. | NCT03293121 | Strength and Coordination Training VS Walking | Completed | No full text |
| 63. | NCT03067545 | Step rate increase | Completed | Exclude, wrong participants, web appendix 2 |
| 64. | NCT04119310 | Lumbar-thrust mobilization VS Sham thrust-mobilization | Unknown | |
| 65. | NCT03620799 | Manual therapy | Unknown | |
| 66. | NCT00166777 | exercise training | Unknown | |
| 67. | NCT04747223 | Run Gait Retraining | Not yet recruiting | |
| 68. | NCT02514005 | Manual therapy VS Dry needling | Completed | Exclude, wrong comparations |
| 69. | NCT04989023 | Blood flow restriction (BFR) with low load resistance training | Not yet recruiting | |
| 70. | NCT02825238 | Supervised exercise program | Completed | Exclude, wrong study design, a feasibility, web appendix2 |
| 71. | NCT04332900 | Elastic hip strap VS Insole | Not yet recruiting | |
| 72. | NCT04589871 | Taping technique VS A supervised exercise protocol | Completed | Exclude, wrong participants, patellofemoral arthritis, web appendix2 |

**Web appendix4: Treatments and treatment categories (ie, classes)**

| Categories (ie, classes) | Category definition | Treatments | Studies |
|---|---|---|---|
| Gait retraining exercise | Exercise in combination with personalized advice on running gait modifications | Gait retraining exercise | Jean-Francois Esculier 2017[1]<br>Jason Bonacci 2017[2]<br>Jenevieve L Roper 2016[3] |
| Foot orthoses +exercise | Exercise in combination with prefabricated orthotics to be placed under the foot in the shoe to support the arch | Foot orthoses +exercise | Jason Bonacci 2017[2]<br>Carsten M Mølgaard 2017[4] |
| Feedback exercise | Exercise in combination with visual and auditory feedback on contraction time and pulling force | Feedback exercise | Henrik Riel 2018[5]<br>Selina L M Yip 2006[6]<br>N Dursun 2001[7] |
| Manipulation treatment + exercise | Exercise in combination with soft tissue treatment, manipulative procedures to the lumbosacral, sacroiliac, hip, ankle, and foot adjustments | Manipulation treatment + exercise | James W Brantingham 2009[8]<br>Gustavo Telles 2016[9] |
| Knee brace +exercise | Exercise in combination with a patellar brace | Knee brace +exercise | Liliam B Priore 2019[10]<br>Victor M Y Lun 2005[11]<br>Mastour S Alshaharani 2019[12] |

| | | | |
|---|---|---|---|
| Kinesiotaping + exercise | Exercise in combination with application of taping/movement to the patella | Kinesiotaping + exercise | Serdar Demirci 2017[13]<br>Eda Akbaş 2011[14]<br>Lucas Simões Arrebola 2019[15]<br>Martin Whittingham 2004[16]<br>Jehoon Lee 2014[17]<br>Marjon Mason 2011[18] |
| Knee arthroscopy and exercise | Exercise in combination with resection of inflamed/scarred medial plicae, abrasion of chondral lesions and shaving of excessive and inflamed synovium | Knee arthroscopy + exercise | Jyrki A Kettunen 2007[19] |
| Education | Education consisted of information or advice given by a health care practitioner on patellofemoral pain, (aggravating) exercise and management of pain/symptoms.<br>Delivered either face to face or in the form of written materials | Education<br><br>Education + exercise | Liliam B Priore 2019[10]<br><br>M S Rathleff 2015[20] |
| Blood flow restriction + exercise | Exercise in combination with the cuff on the proximal thigh and inflated to the prescribed pressure in the resting position of the exercise to be performed | Blood flow restriction + exercise | Lachlan Giles 2017[21] |

| | | | |
|---|---|---|---|
| Whole Body Vibration Exercise | Exercise in combination with vibration stimuli | Whole Body Vibration Exercise | Angel Yañez-Álvarez 2020[22]<br>Ebrahim Rasti 2020[23]<br>Mustafa Corum 2018[24] |
| Exercise therapy | Resistance exercise, that is muscles contracting against resistance provided in the form of weights, bands or body/limb weight with the goal of improving muscle strength, endurance or mass.<br>Programs targeted the lower limb and/or trunk muscles (with/without general aerobic conditioning as warm-up/cool-down).<br>This could be delivered face to face or as a home exercise program | General exercise | Khalil Khayambashi 2012[25]<br>Chen-Yi Song 2009[26]<br>Mahsa Emamvirdi 2019[27] |
| | | Knee exercise | Alexandra Hott 2020[28]<br>Marcelo Camargo Saad 2018[29]<br>Alexandra Hott 2019[30]<br>Reed Ferber 2014[31]<br>Kimberly L Dolak 2011[32]<br>Mehtap Şahin 2016[33]<br>Thiago Yukio Fukuda 2012[34]<br>Thiago Yukio Fukuda 2011[35]<br>Theresa Helissa Nakagawa 2008[36]<br>Lori A Bolgla 2016[37] |
| | | Hip exercise | Alexandra Hott 2020[28]<br>Marcelo Camargo Saad 2018[29]<br>Alexandra Hott 2019[30]<br>Reed Ferber 2014[31]<br>Kimberly L Dolak 2011[32]<br>Chen-Yi Song 2009[26] |

| | | | |
|---|---|---|---|
| | | Knee and hip exercise | Mehtap Şahin 2016[33]<br>Thiago Yukio Fukuda 2012[34]<br>Thiago Yukio Fukuda 2011[35]<br>Theresa Helissa Nakagawa 2008[36] |
| | | Stretching therapy | Lori A Bolgla 2016[37] |
| | | Motor Control Training | F Revelles Moyano 2012[38] |
| | | Target Exercise | Nayra Deise Dos Anjos Rabelo 2017[39]<br>Alireza Motealleh 2019[40]<br>Benjamin T Drew 2017[41]<br>G Syme 2008[42] |
| | | Supervised Exercise | Farzin Halabchi 2015[43] |
| | | Kinetic Chain Exercise | R van Linschoten 2009[44] |
| No treatment | No treatment or continuing with current/planned management | No treatment | Erik Witvrouw 2004[45]<br>R van Linschoten 2009[44]<br>Thiago Yukio Fukuda 2011[35]<br>Khalil Khayambashi 2012[25]<br>Mahsa Emamvirdi 2019[27] |

#### WEB APPENDIX N5. CHARACTERISTICS OF INCLUDED STUDIES

##### Table1. Study characteristic

| RCT | Type of population | Sample size | Main baseline characteristics | Treatments | Outcome measures | Follow-up |
|---|---|---|---|---|---|---|
| Jean-Francois Esculier 2017 | Participants with PFP for at least 3 months | Total n=69<br>Group1:23<br>Group2:23<br>Group3:23 | ***Variable*** group1 group2 group3<br>*Sex, F/M* 15/8 14/9 14/9<br>*Worst pain, VAS 0-10, mean (SD)* 5.8±1.8 7.0±1.4 6.0±2.0<br>*Duration of symptoms(month), mean(rang)* 16.4±16.3 42.2±47.4 28.0±42.4 | Group1: education<br>Group2: exercises + exercise<br>Group3: gait retraining +exercise | •Visual analogue scale (VAS)<br>•Knee Outcome Survey of the Activities of Daily Living Scale (KOS-ADLS) | •20 weeks |
| Jason Bonacci 2017 | Active runners with clinically diagnosed PFP | Total n =16<br>Group1:8<br>Group2:8 | ***Variable*** group1 group2<br>*Sex, F/M* 8/0 4/4<br>*Worst pain, VAS 0-100, mean (SD)* 78.13 ± 6.56 81.88 ± 7.10<br>*Duration of symptoms(month), mean(rang)* 48.25 ± 56.87 46.50 ± 40.72 | Group1: foot orthoses group<br>Group2: gait retraining group | •worst and average pain on a 100 mm visual analogue scale<br>•global improvement, anterior knee pain scale | •12 weeks |
| Jenevieve L Roper 2016 | Recreational runners | Total n =16<br>Group1:8<br>Group2:8 | ***Variable*** group1 group2<br>*Sex, F/M* ? ?<br>*Worst pain, VAS 0-10, mean (Media)* 5.3 (4.2 to 6.4) 4.4 (3.3 to 5.5)<br>*Duration of symptoms(month), mean(rang)* ? ? | Group: exercise group<br>Group2: control group | •knee pain during and/or after running; | •4 weeks |
| Henrik Riel 2018 | Adolescents with PFP | Total n =40<br>Group1:20<br>Group2:20 | ***Variable*** group1 group2<br>*Sex, F/M* 16/4 19/1<br>*Worst pain, VAS 0-100, mean (SD)* 67.1 (11.2) 69.2 (11.6)<br>*Duration of symptoms(week)* > 6 weeks > 6 weeks | Group1: Control group<br>Group2: Feedback group | •Kujala Patellofemoral Scale (KPS); | •4 weeks |
| Selina L M Yip 2006 | Subjects diagnosed with PFP | Total n = 26<br>Group1:13<br>Group2:13 | ***Variable*** group1 group2<br>*Sex, F/M* ? ?<br>*Worst pain, VAS 0-10, mean (SD)* 39.99±19.6 41.99±18.6 | Group1: exercise-only group<br>Group2: biofeedback + exercise group | •patellar alignments and perceived pain severity | •No |

| | | | | | | |
|---|---|---|---|---|---|---|
| | | | *Duration of symptoms(month), mean(rang)* ? ? | | | |
| N Dursun 2001 | Sixty patients (48 women, 12 men; age range, 17–50yr) were selected for this study | Total n = 60<br>Group1:30<br>Group2:30 | ***Variable*** group1 group2<br>*Sex, F/M* 24/6 24/6<br>*Worst pain, VAS 0-10, mean (SD)* 7.3 ±1.5 7.5 ± 1.6<br>*Duration of symptoms(month), mean(rang)* 9.7 ±8.1 10.8 ±7.7 | Group1: control group<br>Group2: biofeedback group | •visual analog scale (VAS)<br>•Functional Index Questionnaire (FIQ) | •12 weeks |
| James W Brantingham 2009 | Males and females 18 to 45 | Total n = 31<br>Group1:13<br>Group2:18 | ***Variable*** group1 group2<br>*Sex, F/M* 2/11 4/14<br>*Worst pain, VAS 0-10, mean (SD)* 2.56 ±2.10 3.06 ±2.38<br>*Duration of symptoms(month), mean(rang)* >3months >3months | Group1: a local manipulative group<br>Group2: a full kinetic chain manipulative therapy group | •AKPS<br>•VAS | •8 weeks |
| Gustavo Telles 2016 | Patients with a clinical diagnosis of anterior knee pain was conducted. | Total n = 18<br>Group1:9<br>Group2:9 | ***Variable*** group1 group2<br>*Sex, F/M* ? ?<br>*Worst pain, VAS 0-10, mean (SD)* 8.0±2.2 6.5±2.6<br>*Duration of symptoms(month), mean(rang)* ? ? | Group1: strengthen the hip muscles and home exercises<br>Group2: strengthening the hip muscles, myofascial techniques, stretching and home exercises | •the numeric pain rating scale (NPRS)<br>•the Lower Extremity Functional Scale (LEFS) | •No |
| Carsten M Mølgaard 2017 | Forty adult individuals ±28 women, 12 men) diagnosed with PFPS. | Total n = 40<br>Group1:20<br>Group2:20 | ***Variable*** group1 group2<br>*Sex, F/M* ? ?<br>*Worst pain, VAS 0-100, mean (SD)* 68-10 64-14<br>*Duration of symptoms(weeks)* >12weeks >12weeks | Group1: the control group (CG), receiving the standard knee targeted exercises<br>Group2: the intervention group (IG), receiving the standard knee targeted exercises combined with foot targeted exercises and foot orthoses | •the Knee Injury and Osteoarthritis Outcomes Score self - reported questionnaire where the sub scale "Pain" was chosen as the primary outcome | •4 months<br>•12 months |
| Liliam B Priore 2019 | Individuals with PFP aged 18-40 years. | Total n = 50<br>Group1:25<br>Group2:25 | ***Variable*** group1 group2<br>*Sex, F/M* ? ?<br>*Worst pain, VAS 0-100, mean (SD)* 73.52 (8.79) 76.60 (11.80)<br>*Duration of symptoms(month), mean(rang)* 50.52(43.02) 51.24 (64.02) | Group1: Brace Group, participants included in this group received a knee brace plus an educational leaflet | •Primary outcome: The Tampa scale for kinesiophobia<br>•Secondary outcomes: The AKPS; The IPAQ-short form; | •4 weeks |

| | | | | | | |
|---|---|---|---|---|---|---|
| | | | | Group2: Leaflet Group , participants received an educational leaflet containing general information about PFP | The FSDT | |
| Victor M Y Lun 2005 | 136 subjects diagnosed with PFPS | Total n =136<br>Group1:34<br>Group2:32<br>Group3:32<br>Group4:31 | ***Variable*** group1 group3<br>*Sex, F/M* ? ?<br>*Worst pain, VAS 0-10, mean (SD)* 4.4 (2.9) 4.2 (3.0)<br>*Duration of symptoms(month), mean(rang)* 11 (8) 10 (7) | Group1: the structured home rehabilitation program only group (E group)<br>Group2: the patellar brace group only (B group)<br>Group3: the structured home rehabilitation program and a patellar brace group (EB group)<br>Group4: the structured home rehabilitation program and a knee sleeve group (ES group) | •knee function (KF)<br>•10-cm visual analogue scale (VAS) pain ratings | •12 weeks |
| Mastour S Alshaharani 2019 | 41 subjects with patellofemoral pain | Total n = 41<br>Group1:21<br>Group2:20 | ***Variable*** group1 group2<br>*Sex, F/M* 12/9 8/12<br>*Worst pain, VAS 0-10, mean (SD)* 4.5±1.5 3.8±0.7<br>*Duration of symptoms(day), mean(rang)* 730 (30,4705) 530 (37,3650) | Group1: the Protonics™ knee brace group<br>Group2: the sport cord group | •Global Rating of Change (GROC) scale<br>•the Kujala score<br>•the Numeric Pain Rating Scale<br>•lateral step-down test | •No |
| Serdar Demirci 2017 | 35 female patients diagnosed with unilateral PFP | Total n =35<br>Group1:18<br>Group2:17 | ***Variable*** group1 group2<br>*Sex, F/M* 18/0 17/10<br>*Worst pain, VAS 0-10, mean (SD)* 3.5 ± 2.01 4.2 ± 1.40<br>*Duration of symptoms(month)* >2 months >2 months | Group1: MWM group<br>Group2: KT group | •Kujala Patellofemoral Pain Scoring | •6 weeks |
| Eda Akbaş 2017 | 31 women with PFPS | Total n = 31<br>Group1:16 | ***Variable*** group1 group2<br>*Sex, F/M* 16/0 15/0 | Group1: kinesio taping group<br>Group2: control group | •Visual analog scale<br>•Anterior Knee Pain Scale / | •No |

| | | | | | | |
|---|---|---|---|---|---|---|
| | | Group2:15 | *Worst pain, VAS 0-10, mean (SD)* 7.08 ±2.49 6.11 ±2.43<br>*Duration of symptoms(month), mean(rang)* 11.80±10.84 14.75±16.32 | | Kujala Scale | |
| Lucas Simões Arrebola 2019 | 43 women with at least a 3-month history of PFPS | Total n = 43<br>Group1:13<br>Group2:14<br>Group3:16 | ***Variable*** group1 group3<br>*Sex, F/M* 13/0 16/0<br>*Worst pain, VAS 0-10, mean (SD)* 4.08 (3.01) 4.13 (3.18)<br>*Duration of symptoms(month)* >3months >3months | Group1: kinesio taping group<br>Group2: KT-LRFT group, using KT® for lateral rotation of the femur and tibia<br>Group3: control group | •Numerical pain rating scale (NPRS)<br>•Kujala Anterior Knee Pain Scale (AKPS) | •12 weeks |
| Martin Whittingham 2004 | Twenty-four men and 6 women aged 17 to 25 years participated in the study | Total n = 30<br>Group1:10<br>Group2:10<br>Group3:10 | ***Variable*** group1 group3<br>*Sex, F/M* ? ?<br>*Worst pain, VAS 0-10, mean (SD)* 7.5 ± 1.0 7.5 ± 0.8<br>*Duration of symptoms(month), mean(rang)* ? ? | Group1: Taping and Exercise group, patella taping combined with a standardized exercise program<br>Group2: Placebo Taping and Exercise group, placebo patella taping and exercise program<br>Group3: exercise Alone group, exercise program alone | •Visual analog scale<br>•Functional Index Questionnaire Scores | •No |
| Jehoon Lee 2014 | 34 (21 men, 13 women) elite athletes | Total n = 34<br>Group1:13<br>Group2:11<br>Group3:10 | ***Variable*** group1 group2 group3<br>*Sex, F/M* 6/7 3/8 4/6<br>*Worst pain, VAS 0-10, mean (SD)* 3.9 (1.5) 4.4 (1.4) 3.8 (1.2)<br>*Duration of symptoms(month), mean(rang)* ? ? ? | Group1: elastic band exercise group<br>Group2: a sling exercise group<br>Group3: control group | •visual analogue scale (VAS)<br>•static and dynamic Q angles<br>•onset time of electromyographic activity of vastus medialis oblique (VMO) and vastus lateralis (VL) | •No |
| Marjon Mason 2011 | 41 subjects took part in this study | Total n = 41, (60 knees)<br>Group1:15 k<br>Group2:15 k | ***Variable*** group1 group2<br>*Sex, F/M* ? ?<br>*Worst pain, VAS 0-10, mean (SD)* 3.43 (2.4) 3.25 (2.1)<br>*Duration of symptoms(month)* >1month >1month | Group1: infrapatellar taping group<br>Group2: quadriceps strengthening group, open chain terminal | •isokinetic quadriceps strength<br>•quadriceps length<br>•pain<br>•pain-free knee flexion angle | •No |

| | | | | | | |
|---|---|---|---|---|---|---|
| | | Group3:15 k<br>Group4:15 k | | extension quadriceps strengthening<br>Group3: quadriceps stretching group, quadriceps stretching<br>Group4: control group | during a step-down | |
| Jyrki A Kettunen 2007 | A total of 56 patients with chronic PFPS. | Total n = 56<br>Group1:28<br>Group2:28 | ***Variable*** group1 group2<br>*Sex, F/M* 17/11 10/18<br>*Worst pain, VAS 0-100, mean (SD)* 39.0 (28.1) 41.4 (28.7)<br>*Duration of symptoms(month), mean(rang)* 54.9 (73.4) 45.0 (74.9) | Group1:an arthroscopy group (N = 28), knee arthroscopy + 8-week home exercise program;<br>Group2: a control group (N = 28), the same 8-week home exercise program only | •the Kujala score on patellofemoral pain and function<br>•visual analog scales (VASs) | •9 months<br>•24 months |
| M S Rathleff 2015 | 121 adolescents from 15-19 years of age | Total n = 121<br>Group1:59<br>Group2:62 | ***Variable*** group1 group2<br>*Sex, F/M* 51/8 46/16<br>*Worst pain, VAS 0-100, Median and IQR* 47 (33;69) 48 (34;64)<br>*Duration of symptoms(month), mean(rang)*<br>2–6nonths 1 5<br>6–12nonths 5 5<br>>12 months 53 52 | Group1: patient education<br>Group2: patient education combined with exercise therapy | •Knee pain<br>•Recovery<br>•Knee injury<br>•Osteoarthritis Outcome Score (KOOS)<br>•Physical Activity Scale (PAS)<br>•EuroQol5-dimensions (EQ-5D)<br>•Satisfaction measured on a five-point Likert scale | •3 months<br>•6 months<br>•12 months<br>•24 months |
| Lachlan Giles 2017 | Sixty-nine participants ±87%) completed the study | Total n = 69<br>Group1:35<br>Group2:34 | ***Variable*** group1 group2<br>*Sex, F/M* ? ?<br>*Worst pain, VAS 0-100, mean (SD)* 55.7 (13.9) 51.4 (15.3)<br>*Duration of symptoms(month), mean(rang)* 31.6 (40.9) 37.8 (55.5) | Group1: BFR group, n=35<br>Group2: standard group, n=34;; | •Kujala Patellofemoral Score<br>•Visual Analogue Scale<br>•isometric knee extensor torque<br>•quadriceps muscle thickness | •6 months |
| Angel Yañez-Álvarez | Adults who had reported anterior knee | Total n = 50<br>Group1:25 | ***Variable*** group1 group2<br>*Sex, F/M* 14/11 12/13 | Group1: exercise group + whole body vibration | •VAS of 10 cm<br>•Douleur Neuropathique-4 items | •No |

| | | | | | | |
|---|---|---|---|---|---|---|
| 2020 | pain | Group2:25 | *Worst pain, VAS 0-10, mean (SD)* 56 ± 20.2 59 ± 13.6<br>*Duration of symptoms(week)* >12weeks >12weeks | Group2: control group | (DN4)<br>•Knee flexion-extension range of movement (ROM)<br>•Lower Extremity Functional Scale (LEFS)<br>•Kujala Patellofemoral Score | |
| Ebrahim Rasti 2020 | Twenty-four male athletes with a diagnosis of PFP | Total n = 24<br>Group1:12<br>Group2:12 | ***Variable*** group1 group2<br>*Sex, F/M* 0/12 0/12<br>*Worst pain, VAS 0-10, mean (SD)* 5.83 ± 0.83 6.08 ± 0.99<br>*Duration of symptoms(month), mean(rang)* ? ? | Group1: WBV + exercise<br>Group2: exercise | •pain (a 0-to-10 linear numerical rating scale (NRS)) | •No |
| Mustafa Corum 2018 | 34 women with PFP , women aged between 18-40 years. | Total n = 34<br>Group1:18<br>Group2:16 | ***Variable*** group1 group2<br>*Sex, F/M* 18/0 16/0<br>*Worst pain, VAS 0-10, mean (SD)* 4.9±1.5 5.0±1.7<br>*Duration of symptoms(month), mean(rang)*<br>*3- 6 months* 2 (11.1) 4 (25.0)<br>*6-12months* 1 (5.6) 0 (0.0)<br>*>12 months* 15 (83.3) 12 (75.0) | Group1: WBV training + home exercise<br>Group2: home exercise | •visual analog scale (VAS)<br>•Kujala Patellofemoral Score (KPS)<br>•Short Form-36 (SF-36) | •6 months |
| F Revelles Moyano 2012 | 74 patients with a pain history more than six months | Total n = 74<br>Group1:35<br>Group2:33<br>Group3:26 | ***Variable*** group2 group3<br>*Sex, F/M* 14/19 5/21<br>*Worst pain, VAS 0-10, mean (SD)* 6.00 ± 1.43 6.05 ± 1.45<br>*Duration of symptoms(month)* >6months >6months | Group1: a classic stretching group<br>Group2: a proprioceptive neuromuscular facilitation and aerobic exercise group<br>Group3: a control treatment received educational materials (control group) | •Knee Society Score<br>•Visual analogue scale<br>•knee range of motion | •No |
| Nayra Deise | Thirty-four women | Total n = 34 | ***Variable*** group1 group2 | Group1: patients in the S group; | •AKPS | •3 months |

| | | | | | | |
|---|---|---|---|---|---|---|
| Dos Anjos Rabelo 2017 | were randomly assigned to two groups. | Group1:17<br>Group2:17 | *Sex, F/M* 17/0 17/0<br>*Worst pain, VAS 0-10, mean (SD)* 6.6 ± 1.0 6.1 ± 1.4<br>*Duration of symptoms(month), mean(rang)* 49.3 ± 40.5 46.2 ± 33.0 | Group2: patients in the MC&S group | •NPRS | •6 months |
| Alireza Motealleh 2019 | a sample of 28 women with unilateral PFPS | Total n = 28<br>Group1:14<br>Group2:14 | ***Variable*** group1 group2<br>*Sex, F/M* 14/0 14/0<br>*Worst pain, VAS 0-100, Median and IQR* 60.00 (40.00-72.5) 65.00 (47.5-70.00)<br>*Duration of symptoms(month)* >2months >2months | Group1: physical therapy exercise program<br>Group2: core neuromuscular training + physical therapy exercise program | •Visual Analog Scale<br>•Kujala patellofemoral questionnaire | •No |
| Benjamin T Drew 2017 | Twenty-six participants | Total n = 26<br>Group1:14<br>Group2:12 | ***Variable*** group1 group2<br>*Sex, F/M* 7/7 8/4<br>*Worst pain, VAS 0-10, mean (SD)* 4.7 (1.68) 5.4 (2.3)<br>*Duration of symptoms(month), Median and IQR* 30 (16.5–75.25) 33 (10.5–54) | Group1: MT group<br>Group2: UC group | •AKPS<br>•Worst NRS<br>•Average NRS<br>•GROC | •No |
| G Syme 2008 | 63 PFPS patients | Total n = 63<br>Group1:21<br>Group2:22<br>Group3:20 | ***Variable*** group1 group2 group3<br>*Sex, F/M* ? ? ?<br>*Average pain, VAS 0-100, mean (SD)* 47.7 (29.6) 51.3(29.4) 59.6(21.8)<br>*Duration of symptoms(month), mean(rang)* 49.0 (37.5) 45.5 (35.3) 50.5 (41.3) | Group1: Selective “vastus medialis oblique activation” group;<br>Group2: General “quadriceps femoris strengthening” group;<br>Group3: Control “no treatment” group | •NRS-101 average pain intensity<br>•McGill pain questionnaire<br>•SF-36 physical component summary score<br>•SF-36 mental component summary score | •No |
| Farzin Halabchi 2015 | Patients both sexes, 18-40 years) with clinically diagnosed PFPS of the duration over 2 months. | Total n =53<br>Group1:26<br>Group2:27 | ***Variable*** group1 group2<br>*Sex, F/M* ? ?<br>*Worst pain, VAS 0-100, mean (SD)* 62.8 (17.9) 53.4 (22)<br>*Duration of symptoms(month), mean(rang)* 31.9 (21.2) 30.1 (22.4) | Group1: n=26 in the intervention (according to the identified risk factors);<br>Group2: n=27 in the control group | •visual analog scale<br>•Kujala patellofemoral score | •12 weeks |
| R van Linschoten | A total of 131 participants were | Total n = 131<br>Group1:65 | ***Variable*** group1 group2<br>*Sex, F/M* 42/23 42/24 | Group1: n1=65 in the intervention group: received as | •7-point Likert scale<br>•numerical rating scale | •3 months<br>•12 months |

| | | | | | | |
|---|---|---|---|---|---|---|
| 2009 | included in the study. | Group2:66 | *Rest pain, VAS 0-10, mean (SD)* 4.14 (2.3) 4.03 (2.3)<br>*Duration of symptoms(month), mean(rang)*<br>*2-6 months (%)* 69.2 66.6;<br>*6-24 months (%)* 31.8 33.4 | standardized exercise program;<br>Group2: n2=66 in the control group: usual care, which comprised a "wait and see" approach of res; | •Kujala patellofemoral score | |
| Erik Witvrouw 2004 | 60 PFPS patients | Total n = 60<br>Group1:30<br>Group2:30 | ***Variable*** group1 group2<br>*Sex, F/M* ? ?<br>*Worst pain, VAS 0-10, mean (SD)* 5.3 (3.2) 5.0 (3.3)<br>*Duration of symptoms(month), Median and IQR* 15.1(1.5-28) 15.1(1.5-28) | Group1: CKC group: only closed kinetic chain exercises<br>Group2: OKC group: only open kinetic chain exercises | •visual analog scales (VAS)<br>•3 functional tests<br>•Muscle Strength Measurement | •5 years |
| Alexandra Hott 2020 | 112 patients with a clinical diagnosis of PFP (≥3 months ±mean 39 months) | Total n = 112<br>Group1:37<br>Group2:39<br>Group3:36 | ***Variable*** group1 group2 group3<br>*Sex, F/M* 24/13 25/14 24/12<br>*Worst pain, VAS 0-10, mean (95%CI)* 6.0 (5.2 to 6.8) 6.5 (5.8 to 7.1) 5.8 (5.1 to 6.5)<br>*Duration of symptoms(month), mean(rang)*<br>*3-6 months* 2 (5) 1 (3) 5 (14)<br>*6-12 months* 7 (19) 5 (13) 11 (31)<br>*12-24 months* 8 (22) 10 (25) 6 (17)<br>*>24 months* 20 (54) 23 (59) 14 (39) | Group1: The hip exercise group<br>Group2: The knee exercise group<br>Group3: The control group receive no prescribed exercise | •Visual Analog Scale (VAS)<br>•Tampa Scale of Kinesiophobia<br>•Knee Self-Efficacy Score<br>•Euro-Qol (EQ-5D-5L) | •12 months |
| Marcelo Camargo Saad 2018 | Forty recreational female athletes between the ages of 18 and 28 years with PFP. | Total n =40<br>Group1:10<br>Group2:10<br>Group3:10<br>Group4:10 | ***Variable*** group1 group2<br>*Sex, F/M* 10/0 10/0<br>*Worst pain, VAS 0-10, mean (SD)* 6.34 ± 1.85 5.05 ± 1.27<br>*Duration of symptoms(month), mean(rang)* ? ? | Group1: Quadriceps strengthening group (QG);<br>Group2: HIP strengthening group (HG);<br>Group3: Stretching group (SG)<br>Group4: Control group (CG) | •visual analog scale<br>•Anterior Knee Pain Scale AKPS)<br>•hip and quadriceps strength<br>•lower limb kinematics | •No |
| Alexandra Hott 2019 | 112 patients with a clinical diagnosis of | Total n = 112<br>Group1:39 | ***Variable*** group1 group2 group3<br>*Sex, F/M* 24/13 25/14 24/12 | Group1: education combined with isolated hip-focused | •Visual Analog Scale (VAS)<br>•Tampa Scale of Kinesiophobia | •3 months |

| | | | | | | |
|---|---|---|---|---|---|---|
| | PFP. | Group2:37<br>Group3:36 | *Worst pain, VAS 0-10, mean (95%CI)* 6.0 (5.2 to 6.8) 6.5 (5.8 to 7.1) 5.8 (5.1 to 6.5)<br>*Duration of symptoms(month), mean(rang)*<br>*3-6 months* 2 (5) 1 (3) 5 (14)<br>*6-12 months* 7 (19) 5 (13) 11 (31)<br>*12-24 months* 8 (22) 10 (25) 6 (17)<br>*>24 months* 20 (54) 23 (59) 14 (39) | exercise<br>Group2: traditional knee-focused exercise<br>Group3: free physical activity | •Knee Self-Efficacy Score<br>•Euro-Qol (EQ-5D-5L) | |
| Reed Ferber 2014 | 199 patients with PFP | Total n = 199<br>Group1:111<br>Group2:88 | ***Variable*** group1 group2<br>*Sex, F/M* 77/34 56/32<br>*Worst pain, VAS 0-10,* 5.12 ±1.66 4.96 ±1.66<br>Mean ±SD (95%CI) (4.76, 5.38) (4.61, 5.30)<br>*Duration of symptoms(month), mean(rang)* >4weeks >4weeks | Group1: HIP treatment group<br>Group2: KNEE treatment group | •visual analog scale<br>•Anterior Knee Pain Scale | •No |
| Kimberly L Dolak 2011 | Thirty-three females with PFPS | Total n = 33<br>Group1:17<br>Group2:16 | ***Variable*** group1 group2<br>*Sex, F/M* 17/0 16/0<br>*Worst pain, VAS 0-10, mean (SD)* 4.6 ± 2.5 4.2 ± 2.3<br>*Duration of symptoms(month), mean(rang)* 36 ± 34 27 ± 34 | Group1: hip strengthening<br>Group2: quadriceps strengthening | •VAS scores<br>•LEFS scores<br>•HABD and HER strength | •8 weeks<br>•3 months |
| Mehtap Şahin 2016 | fifty-five young female patients with patellofemoral pain syndrome were included. | Total n = 55<br>Group1:27<br>Group2:28 | ***Variable*** group1 group2<br>*Sex, F/M* 27/0 28/0<br>*Worst pain, VAS 0-10, median (IQR)* 6 (5.5–7) 7 (6–7)<br>*Duration of symptoms(month), median (IQR)* 6 (4–24) 8 (4–24) | Group1: knee-only exercise programs<br>Group2: hip-and-knee exercises | •Kujala Anterior Knee Pain Scale questionnaire<br>•VAS<br>•isokinetic muscle strength test;<br>•Trendelenburg and muscle tightness tests | •No |
| Thiago Yukio Fukuda 2012 | The study sample included women provoke PFPS. | Total n =54<br>Group1:26<br>Group2:28 | ***Variable*** group1 group2<br>*Sex, F/M* 26/0 28/0<br>*Worst pain, VAS 0-10, mean (SD)* 6.6 ± 1.2 6.2 ± 1.1<br>*Duration of symptoms(month), mean(rang)* 21.0 ± 17.7 23.2 ±19.0 | Group1:KE group<br>Group2: KHE group | •numeric pain rating scale (NPRS)<br>•LEFS<br>•Anterior Knee Pain Scale | •6 months<br>•12 months |

| | | | | | | |
|---|---|---|---|---|---|---|
| | | | | | (AKPS) | |
| Thiago Yukio Fukuda 2011 | The female patients were between 20 and 40 years of age | Total n =70<br>Group1:22<br>Group2:23<br>Group3:25 | ***Variable*** group1 group2 group3<br>*Sex, F/M* 22/0 23/0 25/0<br>*Worst pain, VAS 0-10, mean (SD)* 4.9 ± 2.5 4.9 ± 2.9 5.2 ± 1.6<br>*Duration of symptoms(month), mean(rang)* >3months >3months >3months | Group1: knee group;<br>Group2: knee + hip group<br>Group3:no-treatment group | •(NPRS)<br>•LEFS<br>•(AKPS) | •No |
| Theresa Helissa Nakagawa 2008 | Participants diagnosed with patellofemoral pain syndrome | Total n =14<br>Group1:7<br>Group2:7 | ***Variable*** group1 group2<br>*Sex, F/M* ? ?<br>*Worst pain, VAS 0-10, mean (SD)* 5.0 ± 2.1 5.5 ± 1.5<br>*Duration of symptoms(month), mean(rang)* ? ? | Group1: hip exercise<br>Group2: hip + knee exercise | •visual analogue scale<br>•isokinetic eccentric | •No |
| Khalil Khayambashi 2012 | Females with a diagnosis of bilateral PFP lasting at least 6 months (both knees) | Total n = 28<br>Group1:14<br>Group2:14 | ***Variable*** group1 group2<br>*Sex, F/M* 14/0 14/0<br>*Worst pain, VAS 0-10, mean (SD)* 7.9 ± 1.7 6.6 ± 2.0<br>*Duration of symptoms(month), mean(rang)* >6months >6months | Group1: exercise group<br>Group2: no-exercise control group | •visual analog scale<br>•WOMAC<br>•hip strength | •6 months |
| Chen-Yi Song 2009 | Eighty-nine patients with PFPS | Total n = 89<br>Group1:29<br>Group2:30<br>Group3:30 | ***Variable*** group1 group2 group3<br>*Sex, F/M* 21/8 22/8 26/4<br>*Worst pain, VAS 0-10, mean (SD)* 4.80±2.26 4.85±2.49 4.99±2.18<br>*Duration of symptoms(month), mean(rang)* 41.8±36.1 38.3±34.2 27.7±41.0 | Group1: hip adduction combined with leg-press exercise<br>Group2: leg-press exercise only<br>Group3: no exercise | •visual analog scale (VAS-W)<br>•Lysholm scale scores<br>•VMO morphology | •No |
| Mahsa Emamvirdi 2019 | Sixty-four amateur female volleyball players with PFPS and equal years of exercise experience | Total n = 64<br>Group1:32<br>Group2:32 | ***Variable*** group1 group2<br>*Sex, F/M* 32/0 32/0<br>*Worst pain, VAS 0-10, mean (SD)* 6.1 ± 1.18 6 ± 1.35<br>*Duration of symptoms(month)* >8weeks >8weeks | Group1：VCI exercise training<br>Group2：Written instructions + heat or ice treatment | •Visual analog scale (VAS)<br>•lower extremity performance tests<br>•Peak Torque and Time to Peak Torque | •No |
| Lori A Bolgla 2016 | One hundred eighty-five patients | Total n = 185<br>Group1:105<br>Group 2:80 | ***Variable*** group1 group2<br>*Sex, F/M* 73/32 51/29<br>*Worst pain, VAS 0-10, mean (SD)* 5.2(0.2) 5.0(0.2)<br>*Duration of symptoms(month), mean(rang)* >1months >1months | Group1: hip/core rehabilitation program<br>Group2: knee rehabilitation program | •Visual analog scale (VAS)<br>•Anterior Knee Pain Scale (AKPS)<br>•hip and knee isometric strength | •No |

RCT=randomized controlled trial, 95%CI=95% Confidence Interval, n= number, SD = standard deviation, IQ = interquartile, ? = unknown, F=Female, M=Male

### Table2. Study characteristic (extended)

| 1 | |
|---|---|
| Study | Jean-Francois Esculier 2017 |
| Methods | Group1: education on symptoms management and training modifications (education)<br>Group2: exercise programme in addition to education (exercises)<br>Group3: gait retraining in addition to education (gait retraining) |
| Recruitment of participants | Participants Participants were recruited using advertisements within the running community of Quebec City.<br>Included criteria:<br>(1) be aged 18 to 45 years; (2) report a minimal weekly running distance of 15 km; (3) present with PFP for at least 3 months; (4) experience minimum pain levels of 3/10 on a visual analogue scale (VAS) during running and during three tasks among stairs, kneeling, squatting and resisted knee extension7 and (5) score a maximum of 85/100 on the Knee Outcome Survey of the Activities of Daily Living Scale (KOS-ADLS; the primary outcome).<br>Excluded criteria:<br>(1) symptoms onset following an acute trauma; (2) symptoms believed to originate from patellar tendon27 or menisci28; (3) concurrent lower limb injuries; (4) past history of patellar dislocation or lower limb surgery and (5) presence of rheumatoid, neurological or degenerative diseases. |
| Treatment | Education group: Runners received education on load management and were instructed to self-modify running training according to symptoms. They were asked to increase the frequency of their weekly trainings, to decrease each session's duration and speed and to avoid downhill and stairs running. Run–walk intervals were allowed. Runners were instructed to maintain PFP level at no more than 2/10 during running. Furthermore, pain had to return to pretraining levels within 60 min post-training, without increases in symptoms the following morning. Individualised weekly programmes, which could be modified by runners depending on symptoms, were designed by the treating physiotherapists and progressed based on the evolution of symptoms. Gradually, running distance was increased according to symptoms, before adding speed and hills.29 This specific intervention was provided to all groups. The education group received no other instructions.<br>Exercises group: In addition to the education component, runners were asked to perform a standardised home exercise programme aimed at improving strength, capacity to sustain mechanical load and dynamic control of the lower limbs. The personalised programme included four phases of 2 weeks and gradually progressed through higher difficulty under physiotherapist guidance. Three to four exercises were performed three times per week (maximum 20 min/session), and one exercise (lower limb control) was performed daily (Supplementary file 1). |

| | |
|---|---|
| | Gait retraining group: Together with education， runners received personalised advice on running gait modifications. Runners were asked to increase step rate by 7.5%–10%. If deemed necessary by the physiotherapist (no significant reduction of impact or runner unable to increase step rate), runners were also asked to run softer and to adopt a nonrearfoot strike pattern. Participants had a 10-minute treadmill session with physiotherapist feedback at every visit to the clinic. |
| 2 | |
| Study | Jason Bonacci 2017 |
| Methods | Group1: foot orthoses group<br>Group2: gait retraining group |
| Recruitment of participants | Inclusion criteria:<br>(i) aged 18–40 years; (ii) antero-patella or retro-patella pain that was non-traumatic, longer than six weeks duration and provoked by jogging/running or squatting, hopping/jumping or kneeling or prolonged sitting; (iii) worst pain over the previous week of at least 30/100 mm on a visual analogue scale (0 = no pain, 100 = worst pain imaginable); (iv) running at least 10 km per week; (v) tender over the patellar facet; and (vi) pain on step-down from a 25-cm step or during a double leg squat. All participants used a rearfoot footfall at the time of enrollment.<br>Exclusion criteria:<br>(i) concomitant injury or pathology of other knee structures; (ii) a history of knee surgery; (iii) any foot condition that precludes the use of foot orthoses or running in a minimalist shoe; (iv) a history of use of foot orthoses or minimalist footwear; and (v) pain in and/or referred pain from the hip or lumbar spine. |
| Treatment | Participants were randomly allocated to either gait retraining or to wear foot orthoses.<br>Group1:<br>Participants allocated to wear foot orthoses attended up to four orthoses fitting sessions. Participants received a prefabricated, commercially available full-length orthoses (Vasyli International, Brisbane, Australia)<br>Group2:<br>Participants allocated to gait retraining underwent 10 supervised gait retraining sessions on a treadmill over the first six weeks. Participants attended a physiotherapy clinic twice in the first 4 weeks and once per week thereafter. Gait retraining included two key components: (i) running in a minimalist shoe (Vibram Seeya, Vibram, MA, USA); and (ii) a 10% increase in running cadence. Cadence was controlled by a metronome (Seiko DM51, Seiko Instruments Inc., Japan) and baseline self-selected running cadence was measured via digital video camera footage (Casio Exilim, Casio, Japan). |
| 3 | |
| Study | Jenevieve L Roper 2016 |
| Methods | Group1: exercise group(n=8)<br>Group2: control group(n=8)<br>Protocol Research personnel randomized subjects using a random numbers |

| | |
|---|---|
| | generator to either the control group or experimental group using blocked randomization so that there were an equal number (n=8) of subjects in each group. This trial was a parallel-group trial. Randomization was also done within sex to ensure equal representation and reduce the likelihood of influence of sex-specific variables. |
| Recruitment of participants | Include criteria:<br>Participants self-reported as runners who RFS and reported having mild to moderate chronic, running-related knee pain that occurred during and/or after they ran. All participants reported that running was included in their regular training regimen.<br>Exclude criteria:<br>Subjects were excluded if they had a history of knee surgery on the affected knee, traumatic patellar dislocation, pregnant subjects, and/or any neurological impediments that would influence gait. |
| Treatment | Eight subjects (experimental group) performed eight gait retraining sessions in the Exercise Physiology Lab at the University of New Mexico over a two -week period, while the control group (n=8) performed eight running sessions without the intervention.<br>Group1:<br>The eight gait retraining sessions took place on a treadmill. Run time started at 15 minutes and gradually increased to 30 minutes.<br>the experimental group using mirror feedback and scripted statements, such as "run on your toes" and/or "run on the balls of your feet" were used. If necessary, subjects received additional feedback, such as detailed verbal instructions on how to accurately perform forefoot strike running. The research team visually confirmed that subjects were indeed using a forefoot strike. During the first four sessions subjects were given continuous feedback. During the last four sessions, the feedback was gradually removed.<br>Group2:<br>The control group also performed eight training sessions that were comprised of the same exercise volume (15 minutes gradually increasing to 30 minutes) on the same treadmill. The subjects also ran in front of a mirror, but did not receive any verbal feedback that aided in modifying their running pattern. Subjects in the control group were told "nice job" or "keep it up" to ensure they made no adjustments to their running pattern. |
| 4 | |
| Study | Henrik Riel 2018 |
| Methods | Group1: Control group(n=20)<br>Group2: Feedback group(n=20) |
| Recruitment of participants | Inclusion criteria:<br>15 to 19 year of age; anterior knee pain of nontraumatic origin, which is provoked by at least two of the following activities-prolonged sitting with bent knees or kneeling, squatting, running, jumping, or ascending or descending stairs; tenderness on palpation of the peripatellar borders; pain of more than 6 wk |

| | |
|---|---|
| | duration; and self-reported worst pain during the previous week Q30 mm on a 100-mm visual analog scale (VAS).<br>Exclusion criteria:<br>concomitant pain from other structures in the knee (e.g., ligament, tendon, or cartilage), the hip, or the lumbar spine; previous knee surgery; and self-reported patellofemoral joint instability. |
| Treatment | Forty 15- to 19-yr-old adolescents with PFP were randomized to real-time BandCizeri-iPad feedback on contraction time or not by a physiotherapist.<br>Adolescents received different settings of real-time feedback from the BandCizeri app on the iPad. The feedback group was provided with visual and auditory feedback on contraction time and pulling force, whereas the control group was only provided with real-time feedback on pulling force.<br>Adolescents were instructed to perform three elastic band exercises: seated knee extension and freestanding hip abduction and extension. These exercises have previously been found effective in patients with PFP. |
| 5 | |
| Study | Selina L M Yip 2006 |
| Methods | Group1: exercise-only group(n=13)<br>Group2: biofeedback + exercise group(n=13) |
| Recruitment of participants | Inclusion criteria:<br>The inclusion criteria were men and women who had insidious onset of patellofemoral pain for more than six months. Subjects must have a positive apprehensive test result, and must experience anterior knee pain in at least two of the following activities: ascending stairs, descending stairs, squatting, kneeling, prolonged sitting, hopping and jumping.<br>Excluded criteria:<br>Subjects were excluded if they had degenerative changes on radiography, chondral damage, meniscal lesion, ligamentous instability, previous knee surgery or traumatic injury, and signs of acute inflammation. |
| Treatment | The subjects were then randomly assigned into either the EMG biofeedback + exercise group or the exercise-only group by drawing lots.<br>The exercise protocol was an eight-week home programme comprising the following components: (1) flexibility exercises (stretching of lower limb muscles including quadriceps, hamstring, gastrocnemius, hip adductors and iliotibial band; and mobilization to the patella); (2) strengthening exercises for the quadriceps with emphasis on vastus medialis obliquus recruitment (quadriceps set, terminal knee extension, semi-squatting, wall slide, lunge, step-up, step-down, eccentric hamstring exercise, hip adduction exercise); (3) balance and proprioception training; (4) pylometric and agility training. The subjects were asked to perform 15 minutes of home exercises daily. After 5 minutes of flexibility exercises, the subjects would perform each strengthening exercise for three sets with 10 repetitions. They were required to keep logs of their exercise time and frequency and the research physiotherapist contacted them weekly by phone to monitor their progress. |

| | |
|---|---|
| | The subjects in the EMG biofeedback + exercise group were given a surface EMG biofeedback machine,The subjects were asked to selectively increase the activity of vastus medialis obliquus while maintaining a relatively stable activity in vastus lateralis during the exercises. |
| 6 | |
| Study | N Dursun 2001 |
| Methods | Group1: control group (n =30)<br>Group2: biofeedback group (n =30) |
| Recruitment of participants | Included criteria:<br>1) a diagnosis of unilateral patellofemoral pain syndrome based on the subject having at least 5 of the 7 symptoms listed in table 1,8(2) no evidence of other intra- or extraarticular knee pathologies, determined by physical examination and radiographic evaluation; (3) normal range-of-motion values of the knee, as measured by a goniometer; (4) no history of knee trauma, intraarticular injection therapy, or surgery; and (5) no use of nonsteroid anti-inflammatory drugs within 15 days before treatment began. |
| Treatment | Group2 received electromyographic biofeedback training plus the conventional exercise program.<br>Group1(the control group) received the conventional exercise program only.<br>The conventional exercise program consisted of the following components: (1) strengthening exercises for quadriceps and vastus medialis obliquus (isometric exercises in the form of quadriceps setting, straight leg raising, hip adductor strengthening, terminal knee extension exercises, closed kinetic chain exercises); (2) flexibility training (stretching exercises for hamstrings, gastrocnemius soleus, iliotibial band, quadriceps); (3) proprioception training; and (4) endurance training by bicycling. The conventional exercise program for both groups was supervised by the same physical therapist 5 days a week for the first 4 weeks and 3 days a week thereafter.<br>Biofeedback training was performed with a Myomed 932, aa 2-channel electromyography machine. Clear and full-screen display of the electromyographic signal with a curve was obtained for both the vastus medialis and vastus lateralis. Also, auditory feedback for the vastus medialis was provided. The electrodes were applied to areas of greatest muscle bulk. The 2 active electrodes from each channel were placed as close together as possible along the direction of the fibers of each muscle. The ground electrode was placed equidistant from the corresponding active electrodes. The 30-minute training sessions were held 3 days a week for 4 weeks. |
| 7 | |
| Study | James W Brantingham 2009 |
| Methods | a local manipulative group (group A, n1=13)<br>a full kinetic chain manipulative therapy group (group B, n2=18) |
| Recruitment of participants | Diagnosis/Inclusion Criteria<br>1. Anterior, peripatellar, or retropatellar knee pain of more than 3 months from at least 2 of the following: prolonged sitting, stair climbing, squatting, running, |

| | |
|---|---|
| | kneeling, and hopping/jumping or overuse activities with the pain of any of these activities relieved by rest.<br>2. Insidious or gradual onset of symptoms unrelated to a traumatic incident.<br>3. Presence of pain upon palpation of the patellar facets, on step down from a 25-cm step, or during a double- legged squat.<br>4. X-ray or MRI findings were not required as there is no clear correlation between severity of complaints and arthroscopic or radiologic findings.<br>5. A VAS (worst pain) of ≥5.0 and an AKPS of ≥50.<br>6. The "PARTS" system was used to facilitate determination of concurrent segmental joint dysfunction or"subluxation complex" requiring chiropractic manipulative therapy (CMT).<br>Exclusion criteria:<br>included other disorders such as osteoarthritis, instability, or medial meniscus injuries. Specifically, (1) patellar subluxation/dislocation, locking due to meniscal, internal, and intraarticular derangement or pathology (ie, anterior cruciate ligament injury, ligament tear, laxity, or instability); (2) Osgood-Schlatters or Sinding-Larsen-Johanson syndromes, knee joint effusion, autoimmune or seronegative arthritidies, bursitis, patellar tendonitis; (3) previous knee surgery; (4) neurologic disorders that influence gait and similar disorders; (5) illiteracy or the inability to understand and answer questionnaires and/or consent forms; (6) inability to attend all treatment sessions; and (7) previous physical therapy, chiropractic, or massage therapy in the last 3 months. Prescribed and over-the-counter medication use was allowed if initiated before study entry, although patients were requested to not begin using medications or other common treatments, such as injections, during the study. Similarly, foot orthotics were allowed if currently worn but could not be added/modified during the trial. |
| Treatment | Group A: manipulative therapy (grades I through V) to the local knee joints in conjunction with soft tissue (GISTM) and exercise therapy.<br>Group B: manipulative therapy (grades I through V) to the FKC: lumbosacral, sacroiliac, and (all) lower-extremity joints including the knee, ankle and foot,and exercise and soft tissue (GISTM) treatment .<br>-a dosing level of 1 to 3 times per week<br>-generally 2 to 6 weeks<br>-combined with home exercises.<br>-Exercises were to be performed twice, daily except for Sunday (day off).<br>-Two months after the last or sixth treatment, a follow-up visit obtained outcome measures.<br><br>Manipulative therapy:<br>Protocol 1 (group A):<br>CMT procedures included (all grades of mobilization and/or high velocity low amplitude manipulation starting with lesser grades of mobilization and grades increased per PARTS indications, patient age, and tolerance; cavitation is not |

| | |
|---|---|
| | required nor necessary) the patellofemoral joint (patellar mobilization);tibiofemoral: axial elongation, AP (anterior to posterior) and PA tibial glide, internal/external rotation tibial glide, and varus or valgus tibiofemoral glide. Proximal fibular (proximal fibulotibial) technique included AP and PA, superior (S) to inferior (I) or IS glide.<br>Protocol 2 (group B):<br>For those randomized to group B (in addition to the above manipulative procedures as outlined in protocol 1), manipulative procedures to the lumbosacral, sacroiliac, hip, ankle, and foot adjustments were applied.<br>Soft Tissue Treatment (GISTM).<br>Groups A and B received GISTM procedures as the soft tissue therapy. All providers delivering this modality were required to be trained and certified in GISTM. GISTM was administered to patients with PFPS using methodology as outlined in the Graston Technique Instruction Manual.40Both treatment groups received the same GISTM treatment.<br>The following structures/soft tissue problems were evaluated using the scanning instrument GT 4 and treated using the instrument appropriate for the lesion: vastus medialis oblique fascial restriction, patellar femoral soft tissue joint restriction, iliotibial band insertion at the patellar attachment, rectus femorus adhesion, and/or restriction of the patellar tendon both suprapatellar and infrapatellar. GISTM was applied for a maximum of 3 minutes per site. For the purposes of this pilot study, ice was not applied after GISTM. Post-GISTM treatment allows for either icing or passive stretching posttreatment, with the latter being applied to the areas that had GISTM after therapy<br><br>EXERCISE—IDENTICAL FOR GROUPS 1 AND 2<br>Warm-up is performed at the clinic and at home and consisted of:<br><br>-Isometrics<br>O Three sets of 10 repetitions held for 5 to 10 seconds<br>O Quadriceps setting<br>O Supine straight leg raising, held at 30° to 45°<br>O Short arc quadriceps extension exercises<br>O Isometric hip abduction while standing (4 sets of a 30-second hold)<br><br>-Eccentric strengthening<br>O Squats (bilateral) up to 40°of knee flexion combined with isometric gluteal muscle contractions (4 sets of 10 repetitions)<br>O Standing single-leg squat with slow lowering of knee into flexion (with external rotation for isolating the vastus medialis = eccentric quadriceps strengthening) up to 40°<br>-Stretching<br>O Hamstrings static stretch held 3 repetitions for 30seconds each<br>O Quadriceps static stretch held 3 repetitions for 30seconds each |

| | |
|---|---|
| | O Static stretches will be appropriate for age and fitness (eg, standing quadriceps stretch for a younger fit subject or kneeling quadriceps stretch with an older subject) |
| 8 | |
| Study | Gustavo Telles 2016 |
| Methods | Group A: manipulative therapy (grades I through V) to the local knee joints in conjunction with soft tissue (GISTM) and exercise therapy;<br>Group B: manipulative therapy (grades I through V) to the FKC. |
| Recruitment of participants | Inclusion criteria:<br>Patients were included if they had located pain in the patellar region for at least one month or pain for at least three of the following conditions: squatting, going up and / or down stairs, being seated for long periods, kneeling and pain on palpation in the patellar region and the patellar tendon.<br>Exclusion criteria:<br>Exclusion criteria were previous physiotherapy treatment for patients with these symptoms, knee surgery less than a year ago, full knee prosthesis, previous trauma, patellar fracture, patellar dislocation and rheumatoid conditions. |
| Treatment | GROUP (E)； strengthen the hip muscles and home exercises<br>GROUP (EM)： strengthening the hip muscles, myofascial techniques, stretching and home exercises<br>-treated for a maximum of 10 sessions, each session lasting about 30 minutes<br>-each week for two sessions.<br>-The maximum duration of treatment was five weeks.<br><br>TREATMENT PROTOCOL FOR THE EXERCISE GROUP (E) (3 X 10 repetitions)<br>-Strengthening exercises for hip abductor muscles<br>1- Side lying hip abduction<br>2-Patient in an orthostatic position produces a hip abduction against an elastic resistance band (Thera Band®, black color) _x0001_<br>-Strengthening exercises for lateral hip rotator muscles<br>3- Hip lateral rotation in side lying position (both knee and hip flexed at 60°)<br>4- Hip lateral rotation against elastic band (Thera Band®; black color) while sitting bedside (both hip and knee flexed at 90°)<br>-Strengthening exercises for Gluteus Maximus<br>5- Hip extension in prone position (15°) and knee flexed at 90°<br>Home Based Exercises Patient was oriented to realize all described exercise above without resistance<br>EXERCISE PLUS MYOFASCIAL TECHNIQUE GROUP (EM)<br>-Strengthening exercises:<br>The same exercises as the E Group<br>-Myofascial Release technique<br>1-Rectus femoris muscle<br>2- Tensor fasciae latae muscle |

| | |
|---|---|
| | -Muscle Stretching technique<br>1- Tensor fasciae latae muscle<br>2- Rectus femoris muscle<br>3- Hamstrings muscles<br>Each stretch was held for 30 seconds to the point of tightness or slight discomfort. Each stretch was performed two times, accumulating 60 seconds per stretch. |
| 9 | |
| Study | Carsten M Mølgaard 2017 |
| Methods | the control group (CG): receiving the standard knee targeted exercises;<br>the intervention group (IG): receiving the standard knee targeted exercises combined with foot targeted exercises and foot orthoses; |
| Recruitment of participants | Inclusion criteria:<br>Inclusion criteria were as follows: (a) anterior or retro patellar knee pain for more than twelve weeks; (b)excessive calcaneal eversion measured as calcaneal valgus in relaxed bilateral standing greater than 6°;(c) pain elicited at least by two of the following four tests; (i) Isometric muscle contraction with slightbent knee, (ii) palpation of the patellofemoral joint line, (iii) patellar compression against the femoral bone (iv) active resisted knee extension (d) between 18 and 60 years of age; (e) able and motivated in completing the study.<br>Exclusion criteria:<br>(I) previous knee surgery, except for diagnostic arthroscopy; (II) clinical suspicion of knee osteoarthritis or specific foot and/or knee pathologies (e.g.patellar tendinopathy, lesions of the menisci, cartilage, bone, collateral or cruciate ligaments); (III) and physically or mentally incapable of following the exercise protocol. |
| Treatment | The CG and IG both received three sessions of physiotherapy during a three-month period after inclusion.The three sessions with an experienced physiotherapist were individually adjusted to educate , manual therapy treatment of the soft tissues around the patella , tibio-fibular joint , and soft tissue mobilisation of the iliotibial tract , patellar taping with medialisation of the patella with sports tape and a home exercise program which primarily targeted neuromuscular strength with repetition maximum of 15 - 20 reps .These home -based exercises included squats , semi squat , lunges , knee extensions with rubber band sitting.<br>The IG also received one weekly , supervised , session during the three - month period(a total of 12 sessions).The content of the foot exercise program is illustrated in Figure1.The standardised program was designed with the possibility of individual adjustments in relation to pain and functional level (see foot exercise program , Supplemental Digital Content) |
| 10 | |
| Study | Liliam B Priore 2019 |
| Methods | Brace Group: Participants included in this group received a knee brace plus an educational leaflet; |

| | |
|---|---|
| | Leaflet Group (minimal intervention): The participants received an educational leaflet containing general information about PFP. |
| Recruitment of participants | Inclusion criteria:<br>unilateral anterior knee pain when performing at least two of the following: sitting for prolonged time, squatting, kneeling, running, climbing and descending stairs, and jumping and landing; insidious onset symptoms lasting at least 3 months; the worst knee pain level in the previous week higher than 30mm in a Visual Analogue Scale (VAS 0-100mm);and score lower than in the Anterior Knee Pain Scale (AKPS)<br>Exclusion criteria:<br>history of surgery on any lower limb joint; history of patellar subluxation, clinical evidence of meniscal injury (tibiofemoral joint line tenderness and Thessaly's test at 20° of knee flexion), ligament instability or patellar tendinopathy; osteoarthritis in any lower limb joint assessed clinically; patient-reported spine, hips, ankle or foot pain; presence of neurological disease; no previous physiotherapy treatment for PFP (at least 6 months prior to study commenced). |
| Treatment | ●Brace Group:<br>Participants included in this group received a knee brace plus an educational leaflet (Appendix I). Participants received instructions to use the knee brace for 2 weeks while performing activities of daily living or sports that had previously resulted in knee pain. The type of knee brace used was a commercial flexible model, with approximate weight of 160g (Figure 1). The brace is made of neoprene material and contained four flexible steel rods that offer extra reinforcement aimed at controlling knee joint during movement, and two adjustable flaps to improve compression. The participants received a diary to report the number of hours and activities they used the knee brace and if there were any adverse effects due to its use.<br><br>●Leaflet Group (minimal intervention):<br>The participants received an educational leaflet containing general information about PFP. The content included was related to (i) mechanical and psychological factors in PFP; (ii) load management and; (iii) treatment options (Appendix I). Participants included in this group were instructed to do not use any type of orthoses, brace or bandage in the lower limbs for the period they were involved in the study. The knee brace was offered for the participants of this group after studies completion |
| 11 | |
| Study | Victor M Y Lun 2005 |
| Methods | A structured home rehabilitation program only group (E group);<br>Patellar brace group only (B group);<br>A structured home rehabilitation program and a patellar brace group (EB group);<br>A structured home rehabilitation program and a knee sleeve group (ES group); |

| | |
|---|---|
| Recruitment of participants | ❖Inclusion criteria:<br><br>●History Criteria<br>1. Atraumatic unilateral and/or bilateral peripatellar or retropatellar knee pain for at least 3 weeks but no greater than 2 years<br>2. Patellofemoral knee pain with and/or after activity<br>3. Inactivity patellofemoral pain and/or stiffness, especially with sitting with knees in a flexed position<br>4. No prior history of any significant knee injury (including but not limited to patellar subluxations/dislocations/fractures and ligament or meniscal injuries, and so forth) or knee surgery<br>5. No previous treatment with physiotherapy<br>●Physical Examination<br>1. No or minimal articular or soft-tissue periarticular effusion or bursitis<br>2. No significant joint line tenderness<br>3. No intra-articular ligamentous instability<br>4. Peripatellar tenderness 6 mild inferior patellar pole tendernes<br>●Radiologic Investigation<br>Subjects had standing anterior posterior, lying lateral (or decubitus lateral), and supine skyline plain film x-rays taken of their affected knees. The position of the knee for the lying lateral view was taken with the subject lying on the affected side with the knee bent at 30°. The position of the knee for the skyline view was taken as described by Merchant et al,14 with subjects lying supine with their legs resting over the edge of the x-ray table onto a wooden frame that positioned the knee at 45° of flexion. With the edge of the film cassette placed on the lower leg, the x-ray beam was directed caudally at a 30° angle relative to the femur. The distance of the x-ray tube from the film cassette was about 183 cm.<br><br>❖Exclusion criteria :<br><br>Subjects previously treated with NSAIDs, off-the-shelf knee sleeves or braces, and/or in-shoe orthotics were included in the study. Patients with any bony abnormalities including bony fracture, osteochondritis dissecans, bipartite patella, or osteoarthritis were excluded from participating in the study. |
| Treatment | A structured home rehabilitation program only group(E group)<br>Patellar brace group only (B group)<br>A structured home rehabilitation program and a patellar brace group (EB group)<br>A structured home rehabilitation program and a knee sleeve group (ES group)<br>the E,EB, and ES groups：a standardized lower limb strengthening and stretching home program<br><br>❖Strengthening program：<br><br>The strengthening component of the program consisted of a 6-stage progression of 2-leg eccentric drop squats, then single leg lunges, and finally 1-leg eccentric |

| | |
|---|---|
| | drop squats.<br>●Progressive 6-Stage Drop Squat Program：<br>Stage1：day1–5，Slow drop squat (3-s descent and 5-s ascent)<br>Stage2：day6–10，Fast drop squat (1-s descent and 3-s ascent)<br>Stage3：day11–40，Fast drop squat with 2.5-lb hand weights in each hand, with an addition of 2.5 lb every5 days until 15 lb is reached<br>Stage4：day41–45，Lunge squat with 2.5-lb hand weights in each hand<br>Stage5：day46–50，One-legged fast drop squat<br>Stage6：day51+，One-legged fast drop squat × 2–3 d/wk<br>●the 2-leg drop squats by standing with feet shoulder-width apart and toes pointed forward； then quickly allowed their knees to collapse momentarily to 35°of knee flexion. The quadriceps were then rapidly contracted to stop further collapse； then slowly stood straight.Subjects progressed through the program by initially doing 2 leg squats slowly with body weight;<br>●Then quickly as described above with body weight; then quickly with 5-lb weights in each hand; then quickly with 10-lb weights in each hand; and finally, quickly with 15-lb weights in each hand.<br>●The next progression of the program was single-leg lunges. Stepping forward with 1 leg until the forward knee was flexed to about 90°. Subjects performed the lunges alternating between both legs. Single-leg squats were the final progression of the program.<br>●Single-leg squats were performed like the 2-leg drop squats except standing only on 1 leg. Subjects performed single-leg squats initially with body weight and then with 5-lb weights in each hand.<br>●Subjects progressed through each stage of the program every 5 days. Three sets of 20 repetitions of each exercise on a daily basis. There was a rest of 1 to 2 minutes between each set. This comprised a 40-day build-up phase of the program.<br>●The subjects then maintained their strengthening in a maintenance program by doing the single-leg squats with 5-lb weights in each hand on alternating days.<br>●If there was significant worsening of symptoms at any stage of progression, subjects were instructed to return to the previous step for another 5 days.<br><br>❖stretching program:<br>The stretching component of the rehabilitation program consisted of seated spinal rotations, supine hip external rotation, standing quadriceps stretch, and sitting hamstring stretch. Stretches were performed daily prior to and after the strengthening component of the program. Each stretch was performed passively 3 times, with each stretch held for 30 seconds. |
| 12 | |
| Study | Mastour S Alshaharani 2019 |
| Methods | Group1: the Protonics™ knee brace group<br>Group2: the sport cord group |

| | |
|---|---|
| Recruitment of participants | Inclusion criteria:<br>male or female18-45 years of age; has exhibited patellofemoral pain symptoms for more than 1 month and have a pain level ≥ 3 on the NPRS; has experienced pain during at least 2 functional activities, such as squatting, ascending/descending stairs, and/or running.<br>Exclusion criteria:<br>Individuals who had experienced traumatic injuries to the knee joint or lower extremity, displayed signs or symptoms of a meniscus lesion or ligamentous-related pathology, had been diagnosed with a neurological disorder, diabetes, osteoarthritis, osteoporosis, or rheumatoid arthritis, or reported taking any over-the-counter pain medications during the study period were excluded from the study. |
| Treatment | All subjects completed three study visits, and a total of four measurements were taken at baseline, immediately following the first session, at two weeks, and at 4 weeks.<br>The Protonics™ knee brace group: warm up exercises, specific therapeutic exercises that are part of the Protonic Therapy Program (PTP).<br>❖The warm-up ：<br>● The subject wearing the Protonics™ knee brace set at a moderate resistance level and flexing the knees while sitting, standing, and reclining in the supine and prone positions.<br>●The exercises, were done in sets of 10-15 repetitions, 3 sets per day, 3 times per week, for 4 weeks.<br>●Each set took about 5 minutes for subjects to complete, or 15 minutes per day.<br>❖The PTP has three phases：<br>Phase I：1st Day of Week 1；1 Session；Education、Warm-up、Walking<br>Phase II：Weeks 1-2；3 Sessions per Week；Walking、Hamstring curl in supine, prone, and sitting positions<br>Phase III：Weeks 3-4；3 Sessions per Week；Walking、Hamstring curl in standing position<br>●At the start of each phase, subjects were given detailed instructions on how to perform the warm up and therapeutic exercises and instructed to perform them at home 3 times per week.<br>●During PhaseI or day 1, subjects were asked to walk for 5 minutes or as tolerated at varying speeds while wearing the brace. Subjects also performed the Protonics™ gait and Protonics™ neuromuscular repositioning techniques.<br>●During Phase II or weeks 1 and 2, subjects performed the same Protonics™ techniques, and were asked to walk for 8 minutes or as tolerated at varying speeds and inclines. Subjects were also instructed to perform10-15 repetitions of the hamstring curl in the prone, supine, and seated positions at home.<br>●During Phase III or weeks 3 and 4, subjects were once again asked to perform |

| | |
|---|---|
| | the aforementioned Protonics™ techniques, but this time they were also asked to walk forwards and backwards at varying speeds and inclines for 10 minutes or as tolerated. They were also instructed to do 10-15 repetitions of the standing hamstring curl.<br><br>The sport cord group:<br>❖The same warm-ups and exercises using the sport cord in the supine, standing, sitting, and prone positions.<br>❖The only difference is that subjects were asked to only walk backwards instead of forwards in order to avoid activation of the hip flexor muscle. The appropriate level of resistance for each subject was calculated by multiplying their weight in pounds by 0.3. Subjects were then given either light, medium, or heavy resistance cords according to the following classification scheme: light (pink color) with resistance 3 (R3), 0-30 lbs.; medium (orange color) with resistance 5 (R5) 0-50 lbs.; heavy (yellow color) with resistance 7 (R7) 0-70 lbs.<br><br>All subjects completed three study visits, and a total of four measurements were taken at baseline, immediately following the first session, at two weeks, and at 4 weeks. |
| 13 | |
| Study | Serdar Demirci 2017 |
| Methods | Randomized controlled trial<br>Group1: MWM group(n1=18)<br>Group2: KT group(n2=17) |
| Recruitment of participant | Included criteria:<br>(i) durations lasting longer than two months, (ii) pain scoring three or more according to Visual Analogue Scale (VAS) during at least two activities (prolonged sitting, ascending-descending stairs, squatting, kneeling and jumping-running), (iii) age between 20 and 45 (to reduce the risk of osteoarthritic changes in patellofemoral joint).<br>Excluded criteria:<br>The patients who had meniscus tear, bursitis, ligament injury, patellar tendon lesions, joint degeneration, patellofemoral dislocation and/or recurrent subluxation as well as those who had undergone lower extremity surgery were excluded. Patient with knee pain caused by the hip, lumbal spine or ankle joint were also excluded. |
| Treatment | All the patients were evaluated before the treatment, 45 min after the initial treatment, at the end of the 4-session-treatment during 2-week period and 6 weeks later.<br>Group1:<br>Straight Leg-Raise with Traction: The extremity on which the practice would be performed in supine position was grasped from the ankle level and was, then, |

| | |
|---|---|
| | subjected to traction longitudinally. Afterward, the knee was lifted up passively while in extension and was kept for waiting for a few seconds at the point where tension was felt and was, then, returned to its initial position. The practice was repeated 10 times, and 3 sets of practice at 1-min-intervals were performed. Tibial Gliding: The patients were asked, in the first place, whether or not they felt any pain in the course of the active knee flexioneextension movement while in supine position. In the patients who had pain, the treatment was started on in the position in which no load was transferred onto the knee joint. Each patient was tested in every direction in the course of the active knee flexioneextension movement so as to find out the best pain-free gliding direction (medialelateral part of the tibia, anterioreposterior, internaleexternal rotation). While a hand femur was being fixated in accordance with the treatment direction selected by the therapist, the other hand was subjected to gliding towards tibia, and at that moment, the patient was asked to perform 10 repetitive active knee fexioneextension. The practice was performed by doing 10 repetitions for 3 sets and by providing 1-min-resting time between the sets. Throughout the treatment process, particular attentionwas paid to allowing the position of the hands, the gliding direction and force to remain the same all through the movement process.25,37 If the patient felt no pain in supine position both during and after the practice, the position in which weight/load was conveyed was started to be performed (Fig. 3). This group of patients was also given an additional home exercise program specific to the technique and in the direction selected for the treatment.<br>Group2:<br>To maintain proprioceptive stimulation in the quadriceps (from origo towards insertio) and to alleviate the tension of hamstring muscle, a'Y'-shaped kinesiotape was applied by using the muscle technique. Afterward, 2 pieces of 'I'-shaped tapes were stretched by 75% through the mechanical correction technique and were applied around the patellar circumference in the way that it would allow the patella to move naturally in the femoral cavity while the knee was in flexion. |
| 14 | |
| Study | Eda Akbaş 2011 |
| Methods | Group1: kinesio taping (KT group， n1=15)<br>Group2: control group (n2=16) |
| Recruitment of participants | Inclusion criteria:<br>(1)with a diagnosis of unilateral PFPS participated in this study<br>(2) aged between 17 and 50 years and female<br>Excluded criteria:<br>(1) tendonitis, Osgood-Schlatter syndrome；<br>(2) known articular cartilage；<br>(3) meniscus or ligament damage;<br>(4) history of patellar subluxation or disloca- tion and previous knee surgery. |
| Treatment | Both groups received the same muscle strengthening and soft tissue stretching |

| | |
|---|---|
| | exercises for six weeks.<br>Group 1(KT group):<br>muscle strengthening and soft tissue stretching exercises for six weeks and KT group additionally received kinesio taping at four-day intervals for six weeks.<br>Group 2(Control group):<br>muscle strengthening and soft tissue stretching exercises for six weeks. |
| 15 | |
| Study | Lucas Simões Arrebola 2019 |
| Methods | Group1: kinesio taping (KT group, n1=13)<br>Group2: using KT® for lateral rotation of the femur and tibia (KT-LRFT group, n2=14)<br>Group3: control group (CG group, n3=16). |
| Recruitment of participants | Inclusion criteria:<br>(1)women between the ages of 18 and 45 years who were irregularly active according to the International Physical Activity Questionnaire criteria (Matsudo et al., 2001)<br>(2)The participants each had a history of knee pain for at least 3 months, reported increased pain in at least 3 PFPS-related activities, such as climbing stairs, jumping, kneeling, knee flexion for a long period, or pain on palpation of the medial or lateral facet of the patella, and had a body mass index (BMI) < 29.9 kg/m2<br>Excluded criteria:<br>(1) knee osteoarthritis;<br>(2) previous surgery on the lumbar spine or lower limbs;<br>(3) patellar or quadriceps tendinopathy;<br>(4) patellofemoral instability;<br>(5) ligament or meniscal lesion;<br>(6) and sensitivity to the material used in the application of the KT method;<br>(7) If they presented with other types of pain symptoms not associated with PFPS, that interfered with their performance during the exercise protocol period. |
| Treatment | All groups underwent the same muscle strengthening and motor control procedures for 12 weeks.<br>Rehabilitation protocol:<br>●1-4 weeks:<br>Strengthening exercises (80% 1RM): hip abductors,quadriceps in closed kinetic chain, and tricepssurae (3 sets of 12 repetitions)<br>Strengthening with elastic resistance: quadriceps in open kinetic chain and lateral hip rotators (3 sets of 12 repetitions each)<br>●4-8 weeks:<br>1-4 weeks strengthening plus motor control exercises and core training (3 sets of 30 s each of planks and lateral planks)<br>●8-12 weeks:<br>1-4 weeks strengthening plus progression of the motor control exercises to unstable planes and core training (3 sets of 1 min each of planks and lateral |

| | |
|---|---|
| | planks)<br><br>Group 1(KT group):<br>The KT method applications were performed once a week throughout the 12-week treatment protocol period. the symptomatic leg was chosen for the KT® method.<br>Group2(KT-LRFT group):<br>the taping was performed with the initial anchorage on the suprapatellar region under 0% tension. The therapeutic zone had a tension of >50% involving the entire lateral region of the patella. The final anchorage was on the tibial tuberosity with 0% tension. In the KT-LRFT group, the participant's lower limb was initially in the lateral rotation position before the taping application. The initial anchorage was on the posterior superior iliac spine under 0% tension. The therapeutic zone was in a spiral format throughout the extension of the patients' lower limbs with tension of<50%. The tape involved the greater trochanter, the medial femoral condyle, and the lateral region of the leg, ending at the lateral malleolus region, with the final anchorage placed with 0% tension<br>Group 3(Control group):<br>muscle strengthening and motor control procedures for 12 weeks |
| 16 | |
| Study | Martin Whittingham 2014 |
| Methods | Group1: patella taping combined with a standardized exercise program (Taping and Exercise group, n1=10)<br>Group2: placebo patella taping and exercise program (Placebo Taping and Exercise group, n2=10)<br>Group3: exercise program alone (Exercise Alone group, n3=10) |
| Recruitment of participants | Inclusion criteria:<br>(1) pain on ascending and/or descending stairs, squat- ting, sitting for extended periods of time, or associated with an increase in physical activity.<br>(2) subjects were aged 17 to 25 years (reflecting the age of recruits), men and women, and able to give informed consent.<br>Excluded criteria:<br>(1) a history of subluxation or dislocation of the patella, anterior or posterior cruciate ligament insufficiency, previous knee surgery or meniscal damage, or any other underlying musculoskeletal problems that would have prevented the subject from performing the exercises. |
| Treatment | All subjects attended the physiotherapy department at 08:00 hours each day for the application of tape and performance of exercises, and were advised to remove the tape at the end of the day.<br>The performance of all exercises was completed wearing standard-issue military clothing and training shoes. The program was designed to enhance VMO activation and was graduated so that subjects only progressed to the next exercise when 3 sets of 10 repetitions of the previous exercise could be performed without pain. Non-weight-bearing isometric, inner-range isotonic (from approximately |

| | |
|---|---|
| | 10° flexion to full extension), and straight leg raise quadriceps exercises were included.<br>A variety of weight-bearing exercises were also completed. These included isometric quadriceps contractions in sitting (knees at 90° flexion), bilateral quarter squats (in standing), and unilateral quarter squats (all with a rolled towel between the knees). Further progressions included 506 unilateral quarter-squats (without a rolled towel), controlled step-downs (backwards, sideways, and for-wards), and hip external rotation exercises (standing with the lateral aspect of the affected leg against the wall, hip and knee flexed to 90°, and pushing the leg into the wall). Thirty repetitions of each exercise were completed except for the final exercise, where only 10 repetitions were performed (with a 20-second hold).<br>Stretches for the quadriceps, hamstrings, gastrocnemius, and iliotibial band were also included (20-second hold, 4 repetitions).<br><br>Group 1(Taping and Exercise group):<br>muscle strengthening and soft tissue stretching exercises for six weeks and KT group additionally received kinesio taping at four-day intervals for six weeks.<br>Group2(Placebo Taping and Exercise group):<br>tape was placed across the surface of the patella without patella alignment correction.<br>Group3(Exercise Alone group）:<br>did not have any tape applied,only performed the exercises |
| 17 | |
| Study | Jehoon Lee 2014 |
| Methods | Group1= elastic band exercise group (n=13, EBG)<br>Group2=a sling exercise group (n=11, SEG)<br>Group3= control group (n=10, CG) |
| Recruitment of participants | Inclusion criteria:<br>The criteria used for the diagnosis of PFPS were based on those used in other PFPS studies: diagnosis of PFPS by a medical doctor17); and at least two of the following activities exacerbated their symptoms: prolonged sitting, ascending or descending stairs, squatting, and kneeling.<br>Exclusion criteria:<br>unregulated neurological impairment, knee surgery in the past 2 years, or acquired structural or functional lower limb failures, such as systemic arthritis ligamentous knee injury. |
| Treatment | ●Group 1(EBG group):<br>In the training course, the therapeutic exercises of EBG were performed 3 times a week (30 min) for 8 weeks. Each therapeutic exercise program included a warm-up (stationary bike, 10 min), exercise program (EBG with weight-bearing, 20 min), and cool-down (hamstring self-stretching, 5 min). Therapeutic exercise programs were performed one-on-one with a physical therapist.<br>●Group 2(SEG group):<br>In the training course, the therapeutic exercises of SEG were performed 3 times a |

| | |
|---|---|
| | week (30 min) for 8 weeks. Each therapeutic exercise program included a warm-up (stationary bike, 10 min), exercise program (SEG with weight-bearing, 20 min), and cool-down (hamstring self-stretching, 5 min). Therapeutic exercise programs were performed one-on-one with a physical therapist.<br>●Group 3(CG group):<br>the control group did not perform a therapeutic exercise program. |
| 18 | |
| Study | Marjon Mason 2011 |
| Methods | Group 1(infrapatellar taping group)<br>Group 2(quadriceps strengthening group): Open chain terminal extension quadriceps strengthening<br>Group 3(quadriceps stretching group): Quadriceps stretching<br>Group 4(Control group) |
| Recruitment of participants | Inclusion criteria:<br>had pain for at least one month, located behind or around the patella with two or more of the following: prolonged sitting with knees bent, squatting, kneeling, ascending or descending stairs or running. At least two of these activities should increase the pain in order to be classified as PFPS.<br>Excluded criteria:<br>(1)had meniscal symptoms；<br>(2)required surgery；<br>(3)rheumatoid symptoms;<br>(4)had a synovitis;<br>(5)had back pain. |
| Treatment | During the second week, all subjects followed an identical programme of combined taping, groups and no Day differences for the Control group were established, a series of planned comparisons across Day for each Group on each measure were conducted. Due to the number of planned comparisons, a more conservative significance level of $\alpha \le 0.01$ was adopted to control for increased Type 1 errors. Further analyses (t-test) were conducted to compare the changes in all subjects over the second week of treatment. During this period, all subjects were treated identically using all three modalities. Results quadriceps strengthening and quadriceps stretching.<br>intervention for the first week<br>Group 1(infrapatellar taping group)：<br>Subjects in the taping group had infrapatellar taping applied for one week. The taping was applied immediately distal to the patella with the patient in long-sitting and the quadriceps relaxed. On application, pressure was directed posteriorly and superiorly so as to create a supporting sling for the patella. One layer of 50 mm wide hypoallergenic adhesive non-woven, non-rigid fabric underwrap tape (Therafix, PhysioMed, Ausmedic, Australia) and three layers of 38 mm rigid zinc- oxide sports tape (PhysioMed, Ausmedic, Australia) were applied (Figure 2). Subjects were instructed to keep the tape as dry as possible and to remove the tape if any signs of allergy arose. The taping was replaced by |

| | |
|---|---|
| | the treating physiotherapist if it came off during the week.<br>Group 2(quadriceps strengthening group):<br>Open chain terminal extension quadriceps strengthening<br>Group 3(quadriceps stretching group):<br>Quadriceps stretching<br>Group 4(Control group):<br>received no taping or exercise. All subjects received an overview of knee anatomy and function, especially in relation to the loading of the PF joint and the importance of the quadriceps muscle. They were advised to avoid painful activities like lunges and squats. |
| 19 | |
| Study | Jyrki A Kettunen 2007 |
| Methods | Group1: an arthroscopy group (N = 28), knee arthroscopy + 8-week home exercise program;<br>Group2: a control group (N = 28), the same 8-week home exercise program only; |
| Recruitment of participants | Inclusion criteria:<br>Age 18-40 years Female or male. Characteristic history of PFPS and symptoms lasting at least 6 months. Patellofemoral pain during knee loading physical activity, such as jumping, running, squatting, or going up or down stairs.Patellofemoral pain when the knee was kept in flexion for a prolonged period, with relief on extension.<br><br>Exclusion criteria:<br>Disabling general illness; Reported knee ligamentous or meniscal injuries; Previous knee surgery; Physician diagnosed knee osteoarthritis; A history of patellar dislocation; however, subjects with patellar subluxation are included in the study; Other knee problems than PFPS diagnosed clinically (such as jumper's knee);Other knee problems than PFPS diagnosed radiographically (such as osteochondritis dissecans);Physical therapy for PFPS within the previous 4 weeks; Pregnancy; Competitive athlete |
| Treatment | ❖arthroscopy group:<br>During arthroscopy the following procedures were performed:<br>resection of inflamed/scarred medial plicae, abrasion of chondral lesions and shaving of excessive and inflamed synovium.<br>Minor corrections of the patellofemoral articulation were performed, such as lateral capsular discision in the case of clear lateral patellar subluxation in the beginning of knee flexion.<br>Moreover, possible meniscal tears were treated.<br><br>❖PFPS Exercise protocol:<br>The duration of each session was 30 min. |

| | |
|---|---|
| | ●First part of the program (weeks 1-4)<br>First two exercise weeks:<br>. Exercise every day and repeat the protocol twice daily.<br>. Exercises numbers 1 to 4: 10 - 20 repetitions each.<br>. Exercises numbers 5 to 7: 3 - 5 repetitions each.<br>Weeks three and four:<br>. Exercise every day and repeat the protocol four times a day.<br>. Exercises numbers 1 to 4: 10- 40 repetitions each.<br>. Exercises numbers 5 to 7: 3 -5 repetitions each.<br>●Second part of the program (weeks 5-8)<br>Weeks five and six:<br>. Exercise every day and repeat the protocol twice daily.<br>. Exercises numbers 8 to 11: 10 - 20 repetitions each.<br>. Exercises numbers 12 to 14:3 -5 repetitions each.<br>Weeks seven and eight:<br>. Exercise every day and repeat the protocol four times a day.<br>. Exercises numbers 8 to 11:10 - 40 repetitions each.<br>. Exercises numbers lpto 14:3 -5 repetitions each. |
| 20 | |
| Study | M S Rathleff 2015 |
| Methods | Group1: patient education, n1=59<br>Group2: patient education combined with exercise therapy, n2=62 |
| Recruitment of participants | Inclusion criteria:<br>insidious onset of anterior knee or retropatellar pain of more than 6 weeks duration and provoked by at least two of the following situations: prolonged sitting or kneeling, squatting,running, hopping or stair climbing; tenderness on palpation of the patella, pain when stepping down or double leg squatting; and worst pain during the previous week of more than 30 mm on a 100 mm visual analogue scale (VAS).<br><br>Exclusion criteria:<br>Exclusion criteria were concomitant injury or pain from the hip, lumbar spine or other knee structures; previous knee surgery; self-reported patellofemoral instability; knee joint effusion; use of physiotherapy for treating knee pain within the previous year; or at least weekly use of anti-inflammatory drugs. |
| Treatment | ❖group1:Patient education<br>One physiotherapist delivered the patient education in the two clusters randomised to patient education alone. The standardised patient education was held one-on-one with the adolescents and their parents.<br>It lasted for about 30 min<br>It covered:pain management; how to modify physical activity using pacing and load management strategies; information on optimal knee alignment during daily tasks; and responses to questions from the adolescent or the parents. Adolescents |

| | |
|---|---|
| | also received this information in an eight-page leaflet.<br>❖group2:Patient education + exercise therapy<br>One of two physiotherapists delivered the exercise therapy and patient education in each cluster.<br>The exercise therapy was consisted of a combination of supervised group training sessions and unsupervised home-based exercises.<br>●The supervised group training sessions consisted of neuromuscular training of the muscles around the foot, knee and hip, strength training for the knee and hip, patellofemoral soft tissue mobilisation, and stretching of the muscles around the hip and knee. To progressively match the exercise level to the performance level of each participant, all exercises were available in multiple levels of difficulty .All adolescents started with exercises at level 1 and progressed from there. The progression followed previously described rules.<br>●The unsupervised home exercises consisted of approximately 15 min of quadriceps and hip muscle retraining and stretching. Instructions were given immediately after patient education together with a five-page leaflet with pictures and descriptions of the exercises. The exercises were to be performed each day except on the days of supervised group training. The adolescents were instructed to incorporate the exercises into their normal daily routines.<br>Taping corrections were applied in a predetermined order of anterior tilt, medial tilt, glide and fat pad unloading until the participant's pain was reduced by at least 50%.Tape was only used if adolescents achieved a minimum of 50% reduction in pain measured with a 10 cm VAS during a two-leg squat immediately after application of the tape.<br>The supervised exercises were offered three times per week on school premises immediately after the end of the school day for 3 months. Exercise sessions were offered at three time points during the afternoon of each of three designated weekdays, a total of nine options weekly. The adolescents were told they should continue with the exercises on their own after the intervention period. |
| 21 | |
| Study | Lachlan Giles 2017 |
| Methods | Group1: BFR group, n=35；<br>Group2: standard group, n=34; |
| Recruitment of participants | Inclusion criteria:<br>Participants between 18 and 40 years were included if they experienced PFP as evidenced by the following: atraumatic onset of anterior knee pain for greater than 8 weeks; pain with any two activities, including running, jumping, squatting, kneeling, stair ascent/descent or prolonged sitting; pain with any two of patellar compression; palpation of the peripatellar region; and resisted isometric knee extension when sitting.<br>Exclusion criteria:<br>Participants were excluded if they had coexisting pathology around the knee, including patellar subluxation or dislocation, other sources of anterior knee pain |

| | |
|---|---|
| | (bursa, fat pad), knee surgery, or if they participated in weight training of the legs within the past 6 months (to not include previous non-responders). Participants were excluded on suspicion of patellar tendinopathy, with strong consideration of pain localised to the patellar tendon, increased symptoms with dynamic loads and pain reduction with sustained isometric contraction. Participants were excluded from the study if they were found to be at elevated risk of venous thrombosis (lower limb surgery in the past 6 months, cardiovascular conditions, including high blood pressure (>140/90), diabetes, unexplained chest pain or heart condition, fainting or dizzy spells during physical activity/exercise that causes loss of balance, pregnancy,22 or if exercise was contraindicated. |
| Treatment | BFR group:8 weeks of leg press and leg extension + at 30% of 1RM;<br>Standard group:8 weeks of leg press and leg extension + at 70% of 1 repetition maximum (1RM).<br><br>Features common to BFR and standardised quadriceps strengthening:Each group performed 5 min of 'light' intensity exercise bike to warm up, leg press between 0° and 60° knee flexion and leg extension from 90° to 45° knee flexion. Six rehabilitation sessions were performed with one-on-one supervision from the physiotherapist (three sessions in the first week, then at 2-week intervals), and the remainder of the sessions were performed under group supervision.<br>❖BFR group training:<br>Participants in the BFR group placed the cuff on the proximal thigh and inflated to the prescribed pressure in the resting position of the exercise to be performed (leg press or leg extension). The exercises were performed at approximately 30% of 1RM with the BFR cuff inflated. One set of 30 repetitions (or volitional fatigue), then three sets of 15 repetitions were performed.The cuff remained on for the 30 s rest between sets and was removed after the exercise was completed.<br>❖Standardised quadriceps strengthening group training:<br>The standardised quadriceps strengthening group performed three sets of 7–10 repetitions (approximately 70% of 1RM) with placebo BFR. The placebo was a 5 cm elastic cuff placed firmly around the proximal thigh, with enough room for two fingers between the skin and the cuff. This did not affect the amount of repetitions performed during strength training in preliminary testing. |
| 22 | |
| Study | Angel Yañez-Álvarez 2020 |
| Methods | Group1: exercise group + whole body vibration<br>Group2: control group |
| Recruitment of participants | Inclusion criteria:<br>i) insidious onset of anterior knee pain with a duration greater than 12 weeks; ii) self-reported patellofemoral pain intensity ≥30 mm on the 100 mm Visual Analogue Scale (VAS); iii) pain provoked by at least two of the following situations: prolonged sitting or kneeling, squatting, running, hopping or |

| | |
|---|---|
| | ascending or descending stairs.<br><br>Exclusion criteria:<br>Patellofemoral dislocation or subluxation;<br>knee osteoarthritis (confirmed with radiological tests);<br>knee joint effusion; concomitant injury or pain from the hip, lumbar spine, or other knee structures (meniscus, ligaments, bursa, synovial plica, infrapatellar fat);<br>traumatic lesions of soft tissues or previous orthopaedic surgery in lower limbs;<br>having received knee injections of corticosteroids or hyaluronic acid; cognition or impaired communication;<br>being involved in an ongoing medical-legal dispute. |
| Treatment | Group1：<br>The experimental group performed 12 supervised sessions of hip, knee and core strengthening exercises on a vibration platform 3 times per week during 4 weeks.<br>1.The frequency of the vibration platform was fixed at 40Hz along the study and the amplitude of the vibration platform (peak-to-peak displacement) was set at 2 mm in the first two weeks, and 4 mm during the following two.<br>2.The acceleration peak for these parameters were 3.2G and 6.4G respectively. In terms of force (Newtons) developed for the participants to perform the exercises, this ranged from 748.5 N in a neutral environment to 2395.2 N<br>3. Each session was structured following scheduled phases of warm-up, main active part and, finally, cool-down and stretching. The warm-up phase consisted of different lower limb active exercises to increase the blood flow, muscle temperature and to activate the central nervous system. All exercises in the warm-up and conditioning phases were performed considering the time on the vibration platform in sets of 30s, with 30 s of rest between repetitions.Finally, the cool-down period involved global stretching and trunk and lower limb relaxation, with exercises involving 60 s of work and 6 s of rest and 120 s of work with 12 s of rest,respectively. The total duration of the program was 22 min.<br>Group2:<br>The same protocol but without vibration stimuli. |
| 23 | |
| Study | Ebrahim Rasti 2020 |
| Methods | Group1: WBV + exercise, n1=12<br>Group2: exercise, n=12 |
| Recruitment of participants | Inclusion criteria:<br>Patients were included if they had unilateral patellofemoral pain which was aggravated during at least two of these activities: running, hopping, kneeling, squatting, prolonged sitting, and ascending and descending stairs .An additional criterion was a positive result in at least one of the following: patellar apprehension test, vastus medialis coordination test, or eccentric step-down test<br>Exclusion criteria:<br>contraindications for WBV (kidney stone disease, diabetes, cardiopulmonary |

| | |
|---|---|
| | disease, recent fractures, acute edema, acute disk herniation, using a heart pacemaker, epilepsy), as well as any history of patellar dislocation or subluxation, previous hip, knee or ankle surgery, and any other conditions that may cause anterior knee pain, such as tibiofemoral pathologies |
| Treatment | ●Group1: exercise therapy with whole body vibration training with frequency of 50 Hz and amplitude of 4 mm and for 2 sets 60 seconds with 30 second rest between sets are performed for 4 weeks and three times a week.<br>●Group2: exercise therapy is performed for 4 weeks and three times per week.<br><br>Patients of both groups received 4 weeks of exercise therapy (3 sessions of 45 to 60 min/week) in two phases：<br>●Phase 1 (1st and 2nd weeks):<br>1.3 min warm-up on a stationary bike<br>2.Quadriceps setting, supine straight-leg raises (SLR), side-lying SLR, single-leg stance<br>3.Self-stretches of the Hamstring, quadriceps, and calf muscles<br>●Phase 2 (3rd and 4th weeks)：<br>1.3 min warm-up on a stationary bike<br>2.Self-stretches of the iliotibial band, hamstring ,quadriceps , and calf muscles<br>3. Quadriceps setting<br>4.Dynamic exercises: single-leg cable machine exercises in flexion， extension and abduction directions， bilateral mini-squat , and prone-plank exercise |
| 24 | |
| Study | Mustafa Corum 2018 |
| Methods | Group1: WBV training + home exercise, n1=18<br>Group2: home exercise, n2=16 |
| Recruitment of participants | Inclusion criteria:<br>Among these patients, a total of 40 women aged between 18-40 years diagnosed with either unilateral or bilateral PFP with at least three months symptom history with an average pain during activity (previous week) equal to or greater than 3 cm on a 10 cm visual analogue scale (VAS). Diagnosis of PFP was based on clinical criteria of peri- or retropatellar pain on at least 2 of the following activities such as prolonged sitting, squatting, ascending or descending stairs, kneeling, hopping, or running and positive clinical patellar test.<br>Exclusion criteria:<br>Exclusion criteria were as follows: participation in any systematic training programs such as strengthening and/or aerobic exercises, having received any treatment for PFP within the previous three months, history of lower extremity surgery, lower extremity trauma in the past year, and/or fracture, presence of musculoskeletal diseases such as acute herniated disc or spondylolisthesis, any structural disturbances of the lower extremity (e.g. osteoarthritis in hip or knee joints, prosthesis), central or peripheral neurological pathology and any chronic disease (e.g. diabetes mellitus), presence of gall or kidney stones and intraocular lenses, smoking and excessive alcohol intake, malignancy, and pregnancy. |

| | |
|---|---|
| Treatment | Group1:<br>Exercise therapy:<br>1.WBV training was performed on a triplanar (mostly vertical, Z axis) oscillating vibration platform (Power Plate® pro5™; Power Plate North America, Inc., Northbrook, IL, USA) for 20-30 minutes per session. WBV training was supervised and performed in a clinic three days a week with at least one day between each session for eight weeks (total of 24 sessions).<br>2.Each session lasted approximately 40 minutes including a 10 minutes warm-up and flexibility training period, a 20-30 minutes period of WBV training and 5 minutes cooldown period.<br>Exercise:<br>1.Lunge-step position<br>2.Semi-squat position<br>3.Ball squeeze squat position<br>4.Dynamic squat position<br><br>Group2:<br>Exercise therapy:<br>1.The patients in the control group were instructed on how to perform the exercises at home and supervised individually once a week throughout the program<br>2.Each session was performed bilaterally and lasted approximately 40 minutes including 10 minutes warm-up, 20-30 minutes period of strength exercises with three sets of 10-15 repetitions and 5 minutes cool-down.<br>Exercise:<br>1.lower extremity stretching exercises<br>2.isometric quadriceps setting, knee extensions, double-legged wall squat<br>3.lower extremity stretching exercises |
| 25 | |
| Study | F Revelles Moyano 2012 |
| Methods | Group1(n1=35): a classic stretching group received stretching intervention;<br>Group2(n2=33): a proprioceptive neuromuscular facilitation and aerobic exercise group received proprioceptive neuromuscular facilitation stretching intervention;<br>Group3(n3=26): a control treatment received educational materials (control group) |
| Recruitment of participants | Inclusion criteria:<br>We included subjects with a pain history more than six months, with no previous history of apophysitis or osteoarthritis and with positive results in the physical examination tests: patellofemoral grinding test and patellofemoral compression test |
| Treatment | Participants attended three appointments of 20–60 minutes duration per week for 16 weeks.<br><br>❖Group1:Classic stretching group (CP) |

| | |
|---|---|
| | The classic stretching protocol consisted of active exercises and stretching exercises for hip and knee muscles. The soft tissue stretching protocol was replicated from the study published by Syme et al. including soft tissue stretches for the quadriceps, hamstrings, iliotibial band, gastrocnemius/soleus, and anterior hip structures. In the sequence, each stretch was maintained for 30 seconds and repeated three times.23 Active exercises focused on quadriceps strengthening with and without resistance.<br><br>❖Group2:PNF group<br><br>The hold relax proprioceptive neuromuscular facilitation stretching protocol consisted of passively moving the dominant leg into a position where the subjects felt mild discomfort and holding that position for 30 seconds.25 Subjects were then asked to isometrically contract the stretched muscle for 10 seconds; this was followed by muscle relaxation in the same position for 30 seconds, before being stretched to a new point of mild discomfort. The leg was then released. Additionally, 45 minutes of aerobic exercise, controlled by a personal trainer, were included in each session after the fourth week.<br><br>❖Group3:Control group<br><br>Control group participants received only health educational materials regarding patellofemoral pain as previously described in Song et al. They were advised not to perform or receive any exercise program or intervention too. After the trial they were proposed conventional physiotherapy treatment. |
| 26 | |
| Study | Nayra Deise Dos Anjos Rabelo 2017 |
| Methods | Group1: patients in the S group;<br>Group2: patients in the MC&S group |
| Recruitment of participants | Inclusion criteria:<br>This trial included women aged between 18 to 30 years who had history of anterior knee pain of at least 3 months while performing at least two of the following activities: remaining seated for a prolonged time; going up or downstairs; squatting; running, and jumping .They were also required to score at least 3pointson the Numerical Pain Rating Scale(NPRS)<br>Exclusion criteria:<br>Individuals were excluded if had history of surgery in the lower limbs, recurrent patellar instability, disorders associated with meniscal and/or ligamentous injuries, as well as cardiac or locomotor disorders that could affect the assessment and treatment. |
| Treatment | Group1:<br>Patients in the S group underwent to a set of conventional weight-bearing and non-weight-bearing exercises emphasizing knee extensor, abductor, and lateral rotator hip strengthening. Non-weight-bearing exercises were initiated using ankle weights and elastic bands and progressed to a machine (for quadriceps muscles). |

| | |
|---|---|
| | Group2:<br>Patients in the MC&S group underwent the same strengthening program as the S group, but from the beginning of the treatment were informed about movement control disorders common in women with PFP (ipsilateral trunk lean, contralateral pelvic drop, adduction, and internal rotation of the hip and foot pronation) and were instructed to correct these abnormalities during the execution of the exercises and during daily living activities.<br>All weight-bearing exercises in this group were performed in front of a mirror for the purposes of visual feedback. Also involved verbal feedback and different proprioceptive stimuli such as training in single leg balance for each of the lower limbs, which evolved progressively from stable to unstable surface. This was carried out in 3 sets of 20 seconds in the first week, 30 seconds in the second week, and 40 seconds in third week.<br>The load during training was standardized at 70% of the single maximum Repetition, Maximum load was assessed during the first session, revised on a weekly basis, and adjusted when necessary. Exercises using elastic resistance were standardized for the maximum load that each patient could support while completing 15 repetitions of the exercise, assessed on a weekly basis for adaptations. These criteria were based on the protocol described in a previous study. Patients performed 3 sets of each exercise, with 15 repetitions. Resistance was increased as soon as the exercise became easy to execute. |
| 27 | |
| Study | Alireza Motealleh 2019 |
| Methods | Group1: physical therapy exercise program<br>Group2: core neuromuscular training + physical therapy exercise program |
| Recruitment of participants | Inclusion criteria:<br>Women between 18 and 40 years old with PFPS were included if they had anterior knee pain during at least 2 functional activities including step-up and step-down, squatting, kneeling, jumping, or running, of at least 2 months' duration. The diagnosis of PFPS was confirmed by a positive patellar grind test and tenderness of the medial and lateral patellar facets.24 The patellar grind test has acceptable sensitivity (29%-49%) and specificity (67%-95%).Additional inclusion criteria were pain intensity of more than 3 on a Visual Analog Scale (VAS) and Kujala patellofemoral questionnaire score of 50 to 80 before the intervention. We included participants who did not get analgesic drugs from 2 weeks before the study commencement.<br>Exclusion criteria:<br>Participants were excluded if they had knee meniscus, ligament, or tendon pathologies; subluxation or dislocation of the patella; Sinding-Larsen-Johansson syndrome; Osgood-Schlatter disease; or plica syndrome. Patients also were excluded if they had low back pain, previous pathology, or surgery of the spine or lower limb with or without referred pain. In addition, women with any neuromuscular, rheumatologic, or metabolic diseases (such as diabetic neuropathy) that might affect the outcome measures were excluded. |

| | |
|---|---|
| Treatment | The same treatment : physical therapy exercise program for 4 weeks， 3 exercise sessions per day. Patient attendance at the clinic for supervised exercise therapy was considered 1 of 3 exercise sessions per day. The quality and frequency of home exercise was the same as clinic program. To avoid fatigue, a 3-minute rest was allowed between exercises。<br>The physical therapy exercise program focused mainly on strengthening the knee muscles (mainly quadriceps and hamstring) and flexibility exercises for the gastrocnemius, iliotibial band, and hamstring muscles.<br><br>Group1: physical therapy exercise program<br>◎First （approximately 19 min)<br>1.Hamstring, ITB and gastrocnemius stretching (30-s hold, 5 repetitions)<br>2.Quadriceps setting (10 repetitions, 10-s hold)<br>◎Second (approximately 21 min) ：<br>1.SLR (3 sets, 10 repetitions, 10-s hold)；<br>2.Forward step-up (3 sets, 10 repetitions)<br>◎Last (approximately 25 min)<br>1. Squatting with 30° knee flexion (3 sets, 10-s hold)；<br>2. Lateral step-up (3 sets, 10-s hold)；<br><br>Group2: core neuromuscular training + physical therapy exercise program<br>◎First （approximately 19 min):<br>1.Hamstring, ITB, and gastrocnemius stretching (30-s hold, 5 repetitions);<br>2. Quadriceps setting (10 repetitions, 10-s hold);<br>3.Bridging while holding a small ball between knees (3 sets, 10-s hold);<br>4. Side-lying hip abduction (clam exercise) (6 repetitions, 10-s hold)<br>◎Second (approximately 21 min)：<br>1. SLR (3 sets, 10 repetitions, 10-s hold);<br>2. Forward step-up (3 sets, 10 repetitions);<br>3. Lateral SLR (3 sets, 10 s hold);<br>4. Curl-up while holding a small ball between bent knees (5 repetitions, 10-s hold);<br>◎Last (approximately 25 min):<br>1. Squatting with 30° knee flexion (3 sets, 10-s hold);<br>2. Lateral step-up (3 sets, 10-s hold);<br>3. Isometric hip abduction in standing position (15 repetitions, 5-s hold)；<br>4. Intermittent shoulder flexion/extension while standing on affected limb (15 repetitions, 5-s hold); |

| | |
|---|---|
| | 5. Trunk rotation toward healthy side while maintaining hip internal rotation in standing position on the afflicted leg (15 repetitions, 5-s hold);<br>6. Curl-up while holding a small ball between straight knees (6 repetitions, 10-s hold);<br>7.Lateral curl-up while holding a small ball between straight knees (6 repetitions, 10-s hold). |
| 28 | |
| Study | Benjamin T Drew 2017 |
| Methods | Group1: MT group<br>Group2: UC group |
| Recruitment of participants | Inclusion criteria:<br>Aged 18–40 years<br>•Reported insidious (non-traumatic) onset of anterior or retropatellar knee pain<br>•Pain on two or more of the following activities: prolonged sitting, kneeling, squatting, running, patella palpation, hopping, stair walking, stepping down or isometric quadriceps contraction<br>•Peak hip abduction torque values: Females [18–29 years] ≤ 94.1 Nm; Females [30–39 years] ≤ 75.8 Nm; Males [18–29 years] ≤ 144.1 Nm; Males [30–39 years] ≤ 139 Nm<br>Exclusion criteria:<br>•Presence of inflammatory arthritis; knee pain referred from the hip or lumbar spine; any history of significant knee surgery; other causes of knee pain such as, but not restricted to: meniscal pathologies, quadriceps tendon injuries, patella tendinopathy, tibial tubercle apophysitis; bursitis<br>•Received any treatment within the last 3 months including physiotherapy, podiatry etc |
| Treatment | MT group:<br>attend six supervised sessions of approximately 30 min in duration once per week for 6 weeks at a local hospital.<br>Each week they also performed two additional sessions on non-consecutive days independently at home, with the intervening days allowing adequate rest.<br>During these sessions, participants were given education and justification of the treatment followed by three exercises aimed at targeting coronal, sagittal and transverse strength of the hip using resistance bands. Each week at least one of the exercises would change with the aim of providing variation and minimising tedium. Fidelity was ensured by checking the exercise technique and making corrections to performance prior to these being performed independently at home. Subsequent visits ensured this instruction had been correctly applied or not. Tailoring the intervention based on progressive loading was in line with current recommendations.<br>Participants were issued yellow (least resistance), red or green (most resistance) resistance tubing (66fit Ltd. ™) and were allowed to take it home. To progress the load and resistance, a Borg Rate of Perceived Exertion scale (RPE) was used based on the recommendations when using resistance band. An RPE of >6 was |

| | |
|---|---|
| | considered desirable and participants were monitored after a few repetitions to ensure this was what was being achieved.<br>As participants were stratified for strength, the intervention required participants to perform 10 repetitions within three sets as recommended for strength training. Participants were advised to ensure the time under tension was 8 s (3 s concentric, 2 s isometric hold and 3 s eccentric contraction). Strengthening was performed on each leg alternatively providing a standardised rest between sets.<br>Exercise diaries were issued to participants to provide a reminder of the exercises and to allow a measure of adherence. Participants were asked to document each time each exercise was performed on their diary sheet and return these at each visit.<br>UC group：<br>continued with the same management of their condition as they were planning to receive prior to the commencement of the study. This included planned physiotherapy, podiatry or no intervention, depending upon participant preference.<br>Of the participants in the UC group, 55% received formal physiotherapy treatment, which may or may not have included a strengthening component. The remaining UC participants reported continuing with their normal self-management. |
| 29 | |
| Study | G Syme 2008 |
| Methods | Group A: Selective “vastus medialis oblique activation” group (n1= 21);<br>Group B: General “quadriceps femoris strengthening” group (n2= 22);<br>Group C: Control “no treatment” group (n3= 20); |
| Recruitment of participants | Inclusion criteria:<br>Males and females to be included.<br>Age range 16e40 years.<br>Unilateral or bilateral patellofemoral pain longer than three months.<br>Willing to complete an eight-week rehabilitation program and attend the hospital clinic for assessments.<br>Anterior or retropatellar pain reported on at least two of the following activities: prolonged sitting, ascending or descending stairs, squatting, running, kneeling, and hopping/jumping.<br>In addition to the above, at least two of the following clinical examination findings: Patellar pain with manual compression of the patella against the femur. Patellar tenderness with palpation of the posteromedial and posterolateral borders of the patella. Patellar pain during resisted dynamic knee extension. Patellar pain with manual compression of the patella against the femur during isometric knee extension contraction.<br>Exclusion criteria:<br>Previous knee surgery or trauma.<br>Ligamentous instability and/or internal derangement. (Subjects were referred for arthroscopy or Magnetic Resonance Imaging based on the criteriaoutlined by |

| | |
|---|---|
| | Acton and Craig (2000).)<br>History of patella subluxation or dislocation or patella laxity.<br>Traumatic lesions.<br>Joint effusion when the midpatellar girth was 105% or more than the non-involved knee, where applicable.<br>True knee joint locking and/or giving way.<br>Concurrent medical illness.<br>Inflammatory joint pathology.<br>Infection.<br>Confirmed osteoarthritis of tibiofemoral and/or patellofemoral joints.<br>Knee radiograph abnormalities.<br>Circulatory or neurological abnormalities.<br>Pre/infera or pes anserine bursitis, patella tendonitis, iliotibial tract tendonitis, Osgood Schlatter's disease, Sinding-Larsen Johansson Syndrome,<br>muscle tears or knee plica.<br>Subjects unable or unwilling to give informed to written consent.<br>Subjects awaiting surgery for another lower limb joint problem(s).<br>History of low back, sacroiliac or ankle/feet problems longer than 3 days duration.<br>Subjects already involved in active lower limb training programs.<br>Malignancy.<br>Pregnancy or breast feeding.<br>Ongoing litigation related to lower limb injuries.<br>No patient could be under the care of or have been under the care of another physiotherapist out with the study in the previous one-year prior to the study commencing. |
| Treatment | Selective group:<br>Physiotherapists were instructed that they could use all components of the 'McConnell' approach, such as VMO muscle re-education, flexibility stretching exercises, patella mobilization, patella taping, electromyographic biofeedback and commercially available prefabricated orthotics.<br><br>◎Exercises<br><br>Lower limb exercises emphasising selective activation and retraining of the VMO muscle relative to the VL muscle was undertaken by using a dual channel surface electromyographic biofeedback unit, suggested that a minimum of six sessions should be given during the eight-week treatment period<br>Correction of any dynamic lower limb malalignment and gluteus medius muscle retraining was also encouraged<br>Participants were prescribed daily home exercises and provided with standardised home exercise information sheets 。<br><br>◎Taping<br><br>Patella taping for pain relief in Group A was as advocated by McConnell. |

| | |
|---|---|
| | Non-rigid hypoallergic tape was used to provide skin protection and rigid zinc oxide tape was used for taping corrections. The aim, if possible, was to achieve an immediate reduction in pain intensity of at least 50% 。 Participants were taught to independently apply the taping corrections and were instructed to reapply the tape daily and wear the tape during all waking hours until the pain subsided and exercises could be undertaken pain free.<br><br>◎Stretching<br><br>Soft tissue stretches were included for the quadriceps, hamstrings, iliotibial band, gastrocnemius/soleus and anterior hip structures， The aim was to maintain the stretches for 30 s and repeat each three times over.<br>The patella was mobilised by the physiotherapist and combined with deep frictional massage where necessary. The aim was three repetitions of 60 s each per treatment session 。<br><br>◎Restrictions<br><br>Physiotherapists were informed not to use isokinetic training, electrotherapy, acupuncture or place the subject on a regular gymnasium based on training program for the lower limb(s).<br><br>◎Advice<br><br>All patients were supplied with an advice sheet about patellofemoral pain prior to the start of their treatment.<br><br>General group:<br>Lower limb exercises were based on widely accepted concentric, eccentric and proprioceptive rehabilitation principles 。<br><br>◎Exercises<br><br>The strengthening protocol aimed for 3-5 exercises consisting of 1-3 exercise sets of 10 repetitions, at 60-70% of the one repetition maximum intensity。 This strengthening protocol was suggested as a guideline and physiotherapists were advised to instruct patients to stop any prescribed exercise if pain intensity exceeded 5 on a 0-10 verbal rating scale,(0 ¼ ‘no pain’ and 10 ¼ ‘worst possible pain’) and to adjust training loads accordingly.<br>Correction of any dynamic lower limb malalignment was encouraged during the exercises.<br><br>◎Taping<br><br>A ‘knee sling’ U shaped strapping was applied if necessary, which uses 2.5 cm wide zinc oxide Elastoplast to support the knee and patellofemoral joints. The strapping was only applied during the initial pain control stages.<br><br>◎Stretching |

| | |
|---|---|
| | Soft tissue stretches were included for the quadriceps, hamstrings, iliotibial band, gastrocnemius/soleus and anterior hip structures. The aim was to maintain the stretches for 30s and repeat each three times over. The patella was mobilised by the physiotherapist and combined with deep frictional massage where necessary. The aim was three repetitions of 60 s each per treatment session.<br>◎Restrictions<br>Physiotherapists were informed not to use isokinetic training, electrotherapy, acupuncture, electromyographic biofeedback training or to specifically try and rehabilitate the VMO muscle during the treatment of this group.<br>◎Advice<br>All patients were supplied with an advice sheet about patellofemoral pain prior to the start of their treatment<br>Control group: no treatment |
| 30 | |
| Study | Farzin Halabchi 2015 |
| Methods | Group1: n=26 in the intervention (according to the identified risk factors);<br>Group2: n=27 in the control group |
| Recruitment of participants | Inclusion criteria:<br>Inclusion criteria were (1) age between 18 and 40 years,(2) presence of at least 3 of the following symptoms and signs: peri-/retropatellar pain by walking up or down the stairs, squatting, running, cycling, sitting with knees flexed for a prolonged period, grinding of the patella, and a positive clinical patellar test (such as Clarke test or patellar femoralgrinding test), and (3) pain persisted for more than 2 months.<br>Exclusion criteria:<br>The exclusion criteria, however, were (1) known severe knee osteoarthritis, diagnosed based on clinical and/or radiographic assessment, (2) known rheumatological disease,(3) defined pathological conditions of the knee including patellar tendinopathy, Osgood-Schlatter disease, SindingLarsen-Johansson disease, knee ligamentous injury/instability, (4) a history of knee injuries or knee surgery (ligament reconstruction or medial patellofemoral ligament/other patellar instability surgeries), (5)intake of psychotherapeutic drugs, and (6) previous treatment with exercise therapy, orthosis, and taping in the past 12 months. |
| Treatment | Both groups:<br>Stage 1 (first 6 wk):<br>CKC exercises:<br>Semisquat with 408 knee flexion; 3 sets, 10 repetitions, daily<br>Progressive step-up (10 cm high); 3 sets, 10 repetitions, daily<br>OKC exercises:<br>Straight leg rising in leg external rotation (supine); 3 sets, 10 repetitions, daily<br>Sitting leg extension (108 flexion to full extension); 3 sets, 10 repetitions, daily |

| | |
|---|---|
| | Stage 2 (second 6 wk):<br>CKC exercises<br>Squat with 908 knee flexion; 3 sets, 10 repetitions, daily<br>Progressive step-up (20 cm high); 3 sets, 10 repetitions, daily<br>OKC exercises<br>Straight leg rising in leg external rotation (supine); 3 sets, 10 repetitions, daily<br>Sitting leg extension (908 flexion to full extension); 3 sets, 10 repetitions, daily<br><br>Intervention group (according to the identified risk factors):<br>◎Hamstring tightness:<br>Passive static stretching exercises (unilateral standing, bilateral standing, bilateral sitting) Three alternating repetitions, held for 15 s, daily; continue until the end of the 12-wk program<br>◎Hip flexor tightness:<br>Passive modified lunge stretch and the active prone leg lifts with the knee bent and then straight 10 repetitions, held for 30s, with up to a 30s rest period between repetitions, daily.<br>◎Iliotibial band tightness:<br>Standing stretches in 3 positions of upright standing, overhead clasped hands, and diagonally lowered arms Three alternating repetitions, held for 30 s, daily.<br>◎Gastrocsoleus tightness :<br>Static stretching in forward lunge position 2 repetitions, held for 60 s, daily.<br>◎Excessive foot pronation:<br>Custom-made foot orthoses with the use of foot scan technology.<br>◎Limb length inequality:<br>Correction with lifts no greater than one-half of the difference between limb lengths an interval of 2 weeks between lift therapy adjustments, each lift increment not more than 3-6 mm.<br>◎Patellar malalignment:<br>Patellar taping using the Grelsamer and McConnell technique Nonrigid, hypoallergenic underwrap for skin protection and rigid poly cotton zinc-oxide tape for taping corrections.<br>◎Patellar hypermobility:<br>Patellar taping: The patella will be usually tilted first. A medial glide tape will be then applied to the patella followed by an external rotation tape applied superiorly and inferiorly to improve the seating of the patella in the trochlea. |

| 31 | |
|---|---|
| Study | R van Linschoten 2009 |
| Methods | Group1: n1=65 in the intervention group: received as standardized exercise program;<br>Group2:n2=66 in the control group: usual care, which comprised a“wait and see”approach of res; |
| Recruitment of participants | Inclusion criteria:<br>Inclusion criteria comprised the presence of at least three of the following symptoms: pain when walking up or down stairs; pain when squatting; pain when running; pain when cycling; pain when sitting with knees flexed for a prolonged period of time; grinding of the patella; and a positive clinical patellar test (such as Clarke’s test or patellar femoral grinding test). Symptoms had to have persisted for longer than 2 months but not longer than 2 years.<br>Exclusion criteria:<br>Patients were excluded if they had knee osteoarthritis, patellar tendinopathy, Osgood-Schlatter disease, or other defined pathological conditions of the knee, or had previous knee injuries or surgery. Patients were also excluded if they had already been treated with supervised exercise therapy. |
| Treatment | The intervention group： received as standardised exercise programme for 6 weeks tailored to individual performance and supervised by a physical therapist, and were instructed to practice the tailored exercises at home for 3 months.<br>-The programme consisted of a general warm up on a bicycle ergometer followed by static and dynamic muscular exercises for the quadriceps, adductor, and gluteal muscles.<br>-The programme also included balance exercises and flexibility exercises for major thigh muscles.<br>-Patients exercised for 25 minutes supervised by the physical therapist.<br>-The load of the exercise programme was increased every 2 weeks during the first 6 weeks by increasing the number of repetitions or the intensity of the exercises. The increment of the exercise protocol was monitored by the physical therapist who was guided by pain reaction on exertion.<br>-Patients visited the therapist nine times in 6 weeks.<br>-In addition, they were instructed to practice the exercises daily for 25 minutes over a period of 3 months.<br><br>The control group： were assigned usual care, which comprised a“wait and see”approach of rest during periods of pain and refraining from pain provoking activities.<br><br>Both the intervention group and the control group received standardised information and advice from their GP or sport physician about the background of patellofemoral pain syndrome and its good prognosis as well as advice to refrain from all sports activities that provoke pain. Patients were recommended to use a simple analgesic such as paracetamol when pain was severe and to find |

| | |
|---|---|
| | alternative ways to keep in shape.<br><br>Other interventions—like the use of bandages or braces, insoles, or ice applications, or consumption of medication other than simple analgesics—were allowed in both groups. |
| 32 | |
| Study | Erik Witvrouw 2004 |
| Methods | Group1: CKC group: only closed kinetic chain exercises<br>Group2: OKC group: only open kinetic chain exercises |
| Recruitment of participants | Inclusion criteria:<br>To be eligible for the study, subjects had to experience anterior knee pain for more than 6 weeks and exhibit 2 of the following criteria on initial assessment: pain on direct compression of the patella against the femoral condyles with the knee in full extension, tenderness on palpation of the posterior surface of the patella, pain on resisted knee extension, and pain with isometric quadriceps contraction against suprapatellar resistance with the knee in slight flexion.<br>Exclusion criteria:<br>Patients with knee problems other than patellofemoral pain were excluded from the study. Patients with visually marked cartilage damage on MRI were excluded from this study. Also excluded from this study were patients with a history of a knee operation. None of our patients had a history of trauma, nor did they have a history of subluxation or dislocation |
| Treatment | Prior to the beginning of the OKC and CKC exercise program, a 10-repetition maximum (10 RM) was determined.<br>On that information, patients were instructed to train at 60% of the 10 RM. A new 10 RM was established at the end of a week of training. Each exercise in both training groups was repeated for 3 sets of 10 repetitions. The patient rested 1 minute after the conclusion of each set.<br>In the OKC exercise protocol, each exercise was held isometrically for a count of 6 seconds with a 3-second rest between repetitions.<br>Each exercise in the CKC protocol was performed dynamically with a 3-second rest between repetitions.<br><br>The exercise protocols were as follows:<br>Therapeutic OKC Exercise Program<br>•Maximal static quadriceps contractions (quadriceps setting) with the knee in full extension.<br>•Straight leg raisings with the patient in the supine position.<br>•Short arc movements from 10° of knee flexion to terminal extension.<br>•Leg adduction exercises in the lateral decubitus position.<br><br>Therapeutic CKC Exercise Program<br>•Seated leg press.<br>•Double or single one-third knee bend. |

| | |
|---|---|
| | •Stationary biking.<br>•Rowing machine exercise.<br>•Step up and down exercise.<br>•Progressive jumping exercises on mini trampoline.<br>In both training protocols, the patients were instructed to perform the conventional static quadriceps, hamstrings, and gastrocnemius stretching exercises after each training session. All subjects were instructed to perform 3 repetitions of a 30-second static stretch of these muscle groups |
| 33 | |
| Study | Alexandra Hott 2020 |
| Methods | Group1: The hip exercise group<br>Group2: The knee exercise group<br>Group3: The control group receive no prescribed exercise regime but are encouraged to be physically active according to their own wishes. |
| Recruitment of participants | Inclusion criteria:<br>patients should be 16-40 years of age and have at least three months history of peri- or retropatellar pain with worst pain intensity during previous week of VAS 3 or more. The pain should be provoked by at least two of the following activities: Stair ascent or descent, hopping, running, prolonged sitting, squatting or kneeling. On clinical exam, pain should be present during one of the following: Compression of the patella, palpation of the patellar facets. In patients with bilateral pain the worst knee will be included, and presence of bilateral pain will be documented. One specialist in Physical Medicine and Rehabilitation (PM&R) will perform clinical examinations of all patients. Possible candidates will have a plain x-ray and MRI of the knee joint performed, if this has not already been performed within the previous six months.<br>Exclusion criteria:<br>clinical, x-ray and MRI findings indicative of meniscal or other intra-articular injury, injury to or increased laxity of cruciate or collateral ligaments, or other pathology including: osteoarthritis, Osgood-Schlatter or Sinding-Larsen-Johanssen syndrome, jumpers knee, or of significant knee joint effusion, significant pain from hip or lumbar spine on clinical evaluation, with potential for causing referred pain to the knee, or hindering the patient's ability to perform the prescribed exercises, recurrent patellar subluxation or dislocation, previous surgery to the knee joint, NSAID or cortisone use over an extended period of time, having suffered trauma to the knee joint judged during clinical evaluation to have a significant effect on the presenting clinical condition. Patients having received physiotherapy or other similar treatment for patellofemoral pain syndrome within the previous three months will also be excluded. |
| Treatment | ❖Guiding principles<br>🎬 Dosage is chosen in which the last repetitions are challenging but quality of movement is |

| | |
|---|---|
| | maintained.<br>🎬 Dosage individually adjusted once per week by physiotherapist.<br><br>❖Progression (all exercises):<br><br>🎬 Number of repetitions is increased from 3 sets of 10 repetitions to a maximum of 3 sets of 20 repetitions.<br>🎬 Thereafter resistance is increased using weight cuff or resistance tubing (see individual exercise).<br>o Weight cuffs are available in 0.5 kg increments.<br>o Resistance tubing[b] is selected from 3 possible variants. In order of increasing resistance: red (medium), green (heavy), black (special heavy)<br><br>❖Other details:<br><br>🎬 Repetitions performed dynamically over 2-3 seconds<br>🎬 2-second pause between repetitions.<br>🎬 30 second pause between sets.<br>🎬 Minimum one rest day between sessions<br><br>❖Hip exercises:<br><br>●Hip abduction:<br>Side-lying; Abduct hip, lifting the straight leg upward. Pelvic stabilization emphasized. Weight cuff fastened at ankle;<br>●Hip external rotation (clam-shell):<br>Side-lying; Clam-shell position, hip flexed approx. 60°. Pelvic stabilization emphasized. Weight cuff fastened directly below knee;<br>●Hip extension:<br>Prone; Extend hip, lifting the straight leg upward. Weight cuff fastened at ankle.<br><br>❖Knee exercises:<br><br>●Straight leg raising:<br>Supine Pelvic stabilization emphasized. Weight cuff fastened at ankle.<br>●Terminal knee extension:<br>Supine Knee supported over a cylinder (ø15cm). Knee extends from 10° to 0°. Weight cuff fastened at ankle<br>●Mini-squat to 45°:<br>Standing Back supported against low-friction wall to reduce stabilizing requirements from hip muscles.<br>Feet placed shoulder-width apart and 1 foot-length from wall.<br>Bend knees to 45°.<br>Elastic tubing (length: 2 times distance from lateral femoral epicondyle to medial malleolus) held with one end in each hand, passing beneath both feet. |

| | |
|---|---|
| 34 | |
| Study | Marcelo Camargo Saad 2018 |
| | Group1: Quadriceps strengthening group (QG);<br>Group2: HIP strengthening group (HG);<br>Group3: Stretching group (SG)<br>Group4: Control group (CG) |
| Recruitment of participants | Inclusion criteria:<br>Subjects were included in the study if they were female and had anterior knee pain with a minimum intensity of 3 or greater on the 10-cm visual analog scale (VAS) for at least three months before the study assessment.<br>(1) insidious onset of symptoms; (2) retropatellar or peripatellar pain with at least 2 of the following activities (ascending/descending stairs,<br>running, kneeling, squatting, prolonged sitting or jumping).<br>Exclusion criteria:<br>(1) previous history of knee surgery; (2) history of back, hip, or ankle joint injury or pain; (3) patellar instability; (4) lesion or pain during palpation or test of any structure of knee and (6) any neurological involvement that would affect gait. |
| Treatment | Patients included in the treatment groups participated in two sessions per week for eight weeks with a minimum break of 24 h between sessions. Each treatment session was approximately 50 min in duration, and all sessions were performed individually and supervised by the same physical therapist. For all groups of treatment the weights were increasing as the patient's reported changes in their following the Rating of Perceived Exertion (RPE) scale based on Borg's Scale of effort.<br>❖Quadriceps strengthening group (QG).<br>The exercises in this group focused specifically on quadriceps strengthening.<br>❖HIP strengthening group (HG).<br>This group performed exercises to strengthen hip stabilizing muscles.<br>❖Stretching group (SG).<br>In this group, the physical therapist monitored and stabilized the patients during the stretching exercises for all muscles involved in knee and hip stabilization.<br>❖Control group (CG).<br>Patients included in this group did not have any kind of intervention for eight weeks, but they were tested at the start of the program & at the end like the other 3 groups. At the end of the experiment, a rehabilitation program was made available to all patients. |
| 35 | |
| Study | Alexandra Hott 2019 |
| Methods | Group1: education combined with isolated hip-focused exercise (n = 39);<br>Group2: traditional knee-focused exercise (n = 37); |

| | |
|---|---|
| | Group3: free physical activity (n = 36) |
| Recruitment of participants | Inclusion Criteria:<br>Patients were considered eligible if they were 16 to 40 years old with a minimum 3-month history of PFP (pain, 3 of 10) reproduced by at least 2 activities (stair ascent/descent, hopping, running, prolonged<br>sitting, squatting, kneeling) and present on at least 1 clinical test (compression of the patella, palpation of the patellar facets). For patients with bilateral pain, the worst knee was included.<br><br>Exclusion criteria:<br>(1) clinical, radiographic, or MRI findings indicative of other specific pathology, including meniscal, ligament, or cartilage injury, as well as osteoarthritis, epiphysitis, significant knee joint effusion, or recurrent patellar subluxation or dislocation; (2) significant pain from hip or back hindering the ability to perform the prescribed exercises; (3) previous<br>surgery to the knee joint; (4) nonsteroidal anti-inflammatory drug or cortisone use over an extended period; (5) previous trauma to the knee joint with an effect on the presenting clinical condition; and (6) physiotherapy or other similar exercises for PFP syndrome within the previous 3 months. |
| Treatment | Three sessions per week were performed for 6 weeks: 1 under supervision of the physiotherapist and 2 home sessions, with at least 1 day between sessions. Initial dosage was 3 sets of 10 repetitions for each exercise, with progression to a maximum 3 3 20 repetitions. Each repetition was performed dynamically over 2 to 3 seconds, with a 2-second pause between repetitions and a 30-second pause between sets. Additional resistance thereafter was achieved through weights or elastic tubing depending on the exercise.<br><br>❖Hip-Focused Exercises:<br><br>The hip-focused exercises were based on previous studies and consisted of side-lying hip abduction, hip external rotation (clam shell), and prone hip extension. These exercises were intended to maximally isolate the hip abductors, extensors, and external rotators without stimulating the quadriceps muscles.<br><br>❖Knee-Focused Exercises.:<br><br>The knee-focused exercise regimens were based on previous studies and was intended to maximally isolate the quadriceps muscles. The exercises consisted of straight-leg raises in the supine position, supine terminal knee extensions (from 10of flexion to full extension), and a mini-squat (45of flexion) with the back supported against the wall (to reduce stabilizing requirements from the hip muscles). |

| | |
|---|---|
| | ❖Control Group (Free Physical Activity):<br><br>At randomization, the control group was encouraged by the study physiotherapist to be physically active in accordance with standardized information. |
| 36 | |
| Study | Reed Ferber 2014 |
| Methods | Group1:HIP treatment group(n=111)<br>Group2:KNEE treatment group(n=88) |
| Recruitment of participants | Inclusion Criteria<br>1. Visual analog score rating of pain during activities of daily living during the previous week at a minimum of 3 cm on a 10-cm scale<br>2. Insidious onset of symptoms unrelated to trauma and persistent for at least 4 wk<br>3. Pain in the anterior knee associated with at least 3 of the following:<br>a. During or after activity<br>b. Prolonged sitting<br>c. Stair ascent or descent<br>d. Squatting<br>4. Pain with palpation of the patellar facets or pain during step down from a 20-cm box or during a double-legged squat<br>5. Recreationally active (30 min/d, 3–4 d/wk for the past 6 mo and exclusive of pain)<br><br>Exclusion Criteria<br>1. Meniscal or other intra-articular injury<br>2. Cruciate or collateral ligament laxity or tenderness<br>3. Patellar tendon, iliotibial band, or pes anserine tenderness<br>4. Positive patellar-apprehension sign<br>5. Osgood-Schlatter or Sinding-Larsen-Johansson syndrome<br>6. Evidence of effusion<br>7. Hip or lumbar referred pain<br>8. History of recurrent patellar subluxation or dislocation<br>9. History of surgery to the knee joint<br>10. Nonsteroidal anti-inflammatory drug or corticosteroid use within 24 hours before testing<br>11.History of head injury or vestibular disorder within the last 6 months<br>12.Pregnancy |
| Treatment | For rehabilitation progression, each patient with PFP visited the AT up to 3 times/wk during the 6-week period. The AT asked all patients with PFP to perform their prescribed exercises a minimum of 6 d/wk (including the visits with the AT) for 6 weeks. Compliance was monitored and recorded within the home-exercise rehabilitation booklet.<br>Group1: HIP treatment group(n=111) |

| | |
|---|---|
| | Patients with PFP in the HIP treatment group initially performed non-weight-bearing, muscle-strengthening exercises that focused on activating the hip musculature. Those exercises progressed to weight-bearing exercises, including core-strengthening and balance exercises that were designed to target the core musculature,with specific emphasis placed on stabilizing the core musculature before initiating any of the movements.<br>Week1:<br>Hip abduction—standing, 3 Sets, 10 Repetitions<br>Hip external rotator—standing, 3 Sets, 10 Repetitions<br>Hip external rotator—seated, 3 Sets, 10 Repetitions<br>Week2:<br>Hip abduction—standing, 3 Sets, 10 Repetitions<br>Hip internal rotator—standing, 3 Sets, 10 Repetitions<br>Hip external rotator—standing, 3 Sets, 10 Repetitions<br>Week3:<br>Hip abduction—standing,3 Sets,10 (w/ stronger band) Repetitions<br>Hip internal rotator—standing,3 Sets,10 (w/ stronger band) Repetitions<br>Hip external rotator—standing,3 Sets,10 (w/ stronger band) Repetitions<br>Balancing 2 feet–Airex[a] pad, 3 Sets, 30–45s Seconds.<br>Week4-6:<br>Hip extension at 458—standing,3 Sets,10-15 Repetitions<br>Hip internal rotator—standing,3 Sets,10-15 Repetitions<br>Hip external rotator—standing,3 Sets,10-15 Repetitions<br>Balancing 1 foot—Airex[a] pad,3 Sets, 45-60s Seconds.<br><br>Group2: KNEE treatment group(n=88)<br>Patients in the KNEE treatment group initially performed non-weight -bearing quadriceps strengthening and then progressed to weight -bearing quadriceps-strengthening exercises.<br>Week1:<br>Isometric quadriceps setting, 3 Sets, 10 Repetitions<br>Knee extensions—standing, 3 Sets, 10 Repetitions<br>Double-legged, one-quarter squats, 3 Sets, 10 Repetitions<br>Week2:<br>Isometric quadriceps setting, 3 Sets, 15 Repetitions<br>Double-legged, one-half squats, 3 Sets, 15 Repetitions<br>Terminal knee extension w/ TheraBanda, 3 Sets,15 Repetitions<br>Double-legged, one-quarter squats, 3 Sets, 30 Seconds.<br>Week3:<br>Double-legged, one-half squats, 3 Sets, 10 Repetitions<br>Single-legged, one-quarter squat, 3 Sets, 10 Repetitions<br>Double-legged, one-quarter wall squats, 3 Sets, 10 Repetitions<br>Terminal-knee extension w/ TheraBand, 3 Sets, 10 (w/ stronger band) Repetitions |

| | |
|---|---|
| | Week4:<br>Single-legged, one-half squats, 3 Sets, 10 Repetitions<br>Forward, one-quarter lunge, 3 Sets, 10 Repetitions<br>Lateral step-down (4-inch [3.6 cm] step), No. 3 Sets,10 Repetitions<br>Forward step-down (4-inch [3.6 cm] step), No. 3 Sets,10 Repetitions<br>Double-legged, one-half wall squats, 3 Sets, 30 Seconds.<br>Week5-6:<br>Double-legged wall squat (to max 908 knee flexion), 3 Sets, 30 Seconds.<br>Lateral step-down (6–10 in [5.6–9.6 cm] step), 3 Sets, 15 Repetitions<br>Forward step-down (6–10 in [5.6–9.6 cm] step), 3 Sets, 15 Repetitions<br>Forward one-half full lunge (to maximum 908 of knee flexion), 3 Sets, 15 Repetitions<br>Single-legged one-half full squat (to maximum 908 of knee flexion), 3 Sets, 15 Repetitions |
| 37 | |
| Study | Kimberly L Dolak 2011 |
| Methods | Group1: initial hip strengthening (hip group n1 = 17);<br>Group2: initial quadriceps strengthening (quad group n2=16) |
| Recruitment of participants | Inclusion criteria:<br>The inclusion criteria were that participants needed to exhibit or report (1) anterior or retropatellar knee pain during at least 2 of the activities of stair climbing, hopping, running, squatting, kneeling, and prolonged sitting, (2) an insidious onset of symptoms not related to trauma, (3) pain with compression of the patella, and (4) pain on palpation of patellar facets.<br>Exclusion criteria:<br>Participants were excluded if they had (1) symptoms present for less than 1 month, (2) self-reported other knee pathology, such as cartilage injury or ligamentous tear, (3) a history of knee surgery within the last year, (4) a self-reported history of patella dislocations or subluxations, and (5) any other concurrent significant injury affecting the lower-extremity |
| Treatment | Following the initial testing session, all women were taught and supervised on the first phase of rehabilitation, based on their assignment to either the hip group or quad group. Both groups received the same flexibility exercises. A seated hamstring stretching, standing quadriceps stretch, and standing wall stretch for the triceps were performed throughout the 8-week program. Flexibility exercises were performed 3 times for 30 seconds each, prior to strengthening exercises. Participants performed rehabilitation exercises 1 day a week with an investigator and 2 days a week at home, for a total of 3 exercise sessions each week.<br><br>❖Group1:The hip group<br><br>Week1:<br>Sidelying combination hip abduction and external rotation,3 sets of 10 repetitions<br>Standing hip abduction,3 sets of 10 repetitions<br>Seated hip external rotation,3 sets of 10 repetitions |

| | |
|---|---|
| | Week2:<br>Standing hip abduction with 3% body weight,3 sets of 10 repetitions<br>Sidelying hip abduction with 3% body weight,3 sets of 10 repetitions<br>Seated hip external rotation with 3% body weight,3 sets of 10 repetitions<br>Week3:<br>Sidelying hip abduction with 5% body weight,3 sets of 10 repetitions<br>Seated hip external rotation with 5% body weight,3 sets of 10 repetitions<br>Quadruped hydrant (combined hip abduction and external rotation)， 3 sets of 10 repetitions<br>Week4：<br>Sidelying hip abduction with 7% body weight,3 sets of 10 repetitions<br>Seated hip external rotation with 7% body weight,3 sets of 10 repetitions<br>Quadruped hydrant with 3% body weight,3 sets of 10 repetitions<br>Week5:<br>Single-leg balance with front pull,3 sets of 30 seconds<br>Wall slides with resistance,3 sets of 10 repetitions<br>Lateral step-downs off a 10-cm step,3 sets of 10 repetitions<br>2-leg calf raises,3 sets of 10 repetitions<br>Week6:<br>Single-leg balance with diagonal pull,3 sets of 30 seconds<br>Single-leg mini-squats,3 sets of 10 repetitions<br>Lateral step-downs off a 15.25-cm step,3 sets of 10 repetitions<br>Single-leg calf raises,3 sets of 10 repetitions<br>Week7:<br>Single-leg standing on Airex pad,3 sets of 30 seconds<br>Lunges to a 20.3-cm step,3 sets of 10 repetitions<br>Lateral step-downs off a 15.25-cm step with resistance,3 sets of 10 repetitions<br>Single-leg calf raises off a step,3 sets of 10 repetitions<br>Week8:<br>Single-leg standing on Airex pad with diagonal pull,3 sets of 30 seconds<br>Lunges to a 10-cm step,3 sets of 10 repetitions<br>Lateral step-downs off a 20.3-cm step,3 sets of 10 repetitions<br>Single-leg calf raises on Airex pad,3 sets of 10 repetitions<br><br>❖Group2:The knee group<br><br>Week1:<br>Quad sets,3 sets of 10 repetitions<br>Short-arc quads,3 sets of 10 repetitions<br>Straight leg raises,3 sets of 10 repetitions<br>Week2:<br>Short arc quads with 3% body weight,3 sets of 10 repetitions<br>Straight leg raises with 3% body weight,3 sets of 10 repetitions<br>Terminal knee extensions with 3% body weight,3 sets of 10 repetitions<br>Week3: |

| | |
|---|---|
| | Short-arc quads with 5% body weight,3 sets of 10 repetitions<br>Straight leg raises with 5% body weight,3 sets of 10 repetitions<br>Terminal knee extensions with 5% body weight 3 sets of 10 repetitions<br>Terminal knee extensions with 7% body weight 3 sets of 10 repetitions<br>Week4:<br>Short-arc quads with 7% body weight 3 sets of 10 repetitions<br>Straight leg raises with 7% body weight 3 sets of 10 repetitions<br>Terminal knee extensions with 7% body weight 3 sets of 10 repetitions<br>Week5:<br>Single-leg balance with front pull,3 sets of 30 seconds<br>Wall slides with resistance,3 sets of 10 repetitions<br>Lateral step-downs off a 10-cm step,3 sets of 10 repetitions<br>2-leg calf raises,3 sets of 10 repetitions<br>Week6:<br>Single-leg balance with diagonal pull,3 sets of 30 seconds<br>Single-leg mini-squats,3 sets of 10 repetitions<br>Lateral step-downs off a 15.25-cm step,3 sets of 10 repetitions<br>Single-leg calf raises,3 sets of 10 repetitions<br>Week7:<br>Single-leg standing on Airex pad,3 sets of 30 seconds<br>Lunges to a 20.3-cm step,3 sets of 10 repetitions<br>Lateral step-downs off a 15.25-cm step with resistance,3 sets of 10 repetitions<br>Single-leg calf raises off a step,3 sets of 10 repetitions<br>Week8:<br>Single-leg standing on Airex pad with diagonal pull,3 sets of 30 seconds<br>Lunges to a 10-cm step,3 sets of 10 repetitions<br>Lateral step-downs off a 20.3-cm step,3 sets of 10 repetitions<br>Single-leg calf raises on Airex pad,3 sets of 10 repetitions |
| 38 | |
| Study | Mehtap Şahin 2016 |
| Methods | Group1: knee-only exercise (Group A) programs;<br>Group2: hip-and-knee exercises (group B) |
| Recruitment of participants | Inclusion Criteria:<br>1) sedentary female patients ranging from age 20 to 45;<br>2) patients with a full range of motion of the knee joints;<br>3) presence of anterior or retropatellar knee pain during at least 3 of the following activities: ascending/descending stairs, squatting, hopping/running, and prolonged sitting;<br>4) insidious onset of symptoms unrelated to a traumatic incident and persistence of symptoms for at least 4 weeks;<br>5) a score of at least 3 on the visual analog scale (VAS);<br>6) presence of pain on palpation of the patellar facets;<br>7) presence of pain on stepping down from a 25-cm step or double-legged squat |

| | |
|---|---|
| | Exclusion Criteria:<br>1) current significant injury affecting lower limb joints;<br>2) surgery of the knee joint;<br>3) signs or symptoms or MRI findings of intraarticular pathologic conditions such as effusion, meniscal, or cruciate or collateral ligament involvement;<br>4) tenderness of the patellar tendon or iliotibial band or pesanserinus tendon;<br>5) patellar subluxation or dislocation;<br>6) signs of patellar apprehension;<br>7) referred pain with hip pain, or back pain, or sacroiliac joint pain;<br>8) acute strain or sprain;<br>9) current use of nonsteroid anti-inflammatory drugs or corticosteroids. |
| Treatment | ❖Patient education of both groups<br><br>Patient education consisted of recommendations to both groups, such as avoiding prolonged sitting, low-chair sitting, cross-legged sitting, kneeling, stair-climbing, and squatting. Only a cold pack was prescribed for pain control. Other pain medications were restricted.<br><br>❖Exercise program for both groups<br><br>A therapist-supervised exercise program of thirty sessions (5 days a week for 6 weeks) was given to both groups. Each session started with a 5-min warm-up, continued with 20 min of lower extremity stretching and strengthening exercises, and concluded with a 5-min cool-down. Elastic resistance exercises were performed using green<br>TheraBand latex exercise bands (TheraBand, USA). Exercises utilizing elastic resistance were standardized to the maximum resistance at which each patient was able to perform 10 repetitions of the exercise. The maximum load and resistance for all strengthening exercises were evaluated during the first treatment session.<br><br>❖Exercise program for the knee-only exercise group :<br><br>Group 1 = knee exercise (n=25)<br>1. Lower extremity stretches<br>Patients were asked to perform 3 repetitions of supine hamstring stretching and standing quadriceps, iliotibial band, and gastrocnemius stretching exercises twice a day. Patients were asked to hold the muscle in contraction for 10 s in each exercise.<br>2. Isometric quadriceps-strengthening exercise<br>A towel was placed under the knees in the supine position. Patients were asked to repeat the exercise twice a day with 20 initial repetitions, after which 5 repetitions were added every following week. Patients were asked to hold the muscle in contraction for 10 s in each exercise.<br>3. Straight leg raise exercise<br>Patients were instructed to perform the exercise twice a day with 10 repetitions. |

| | |
|---|---|
| | Patients were asked to hold the muscle in contraction and work up to holding the contraction for 3.5 s in each exercise.<br>❖Exercise program for the hip-and-knee exercise group<br>Group 2= hip plus knee exercises(n=25)<br>1. Hip abductor-strengthening exercises<br>Patients were instructed to perform 5 repetitions of 30°–35 standing hip abductions with an elastic resistance exercise twice a day ; 5 repetitions were added every following week. Patients were asked to hold the muscle in contraction for 3.5 s in each exercise.<br>2. Hip external rotator-strengthening exercises<br>A towel was placed between the thighs. Patients were instructed to externally rotate the hip to approximately 30° and then hold the contraction for 3.5 s. Patients were instructed to perform this exercise twice a day with an initial 5 repetitions; 5 repetitions were added every<br>following week. After five sessions a week for 6 weeks (30 sessions) with the supervised exercise program at the clinic, patients were instructed to continue with 6 weeks of an at-home exercise program and follow-up visits. |
| 39 | |
| Study | Thiago Yukio Fukuda 2012 |
| Methods | Group1: KE group<br>Group2: KHE group |
| Recruitment of participants | Inclusion Criteria:<br>The study sample included women 20 to 40 years of age who had a history of anterior knee pain for at least 3 months and reported increasing pain in 2 or more activities that commonly provoke PFPS.<br>These activities included ascending and descending stairs, squatting, kneeling, jumping, long sitting, isometric knee extension contraction at 60° of knee flexion, and pain on palpation of the medial and/or lateral facet of the patella.All patients included in the trial were sedentary, defined as not having practiced physical activity (aerobic and strengthening exercises) any day of the week for at least 6 months previously.<br><br>Exclusion Criteria:<br>Participants were excluded if they had a neurological disorder; injury to the lumbosacral region, hip, or ankle; rheumatoid arthritis, a heart condition, or previous surgery involving the lower extremities; or were pregnant or using corticosteroids or anti-inflammatory medication. Women who had other knee pathologies, such as patellar instability, patellofemoral dysplasia, meniscal or ligament tears, osteoarthritis, or tendinopathies, were also excluded. |
| Treatment | The KE and KHE groups completed 12 treatment sessions, provided 3 times per week for 4 weeks. The patients performed exercises solely during physical therapy and did not perform exercises at home. After the 4-week treatment program, the patients were instructed to maintain their normal daily activities |

| | |
|---|---|
| | without performing a home exercise program.<br>KE group:<br>• Stretching (hamstrings, plantar flexors, quadriceps, and iliotibial band), 3 repetitions of 30 s<br>• Seated knee extension from 90° to 45°, 3 sets of 10 repetitions*<br>• Leg press from 0° to 45°, 3 sets of 10 repetitions*<br>• Squatting from 0° to 45°, 3 sets of 10 repetitions*<br>• Single-leg calf raises, 3 sets of 10 repetitions*<br>• Prone knee flexion,†3 sets of 10 repetitions*<br><br>KHE group:<br>• Same protocol as the KE group<br>• Hip abduction with weights (sidelying), 3 sets of 10 repetitions*<br>• Hip abduction against elastic band (standing), 3 sets of 10 repetitions‡<br>• Hip lateral rotation against elastic band (sitting), 3 sets of 10 repetitions‡<br>• Hip extension (machine), 3 sets of 10 repetitions* |
| 40 | |
| Study | Thiago Yukio Fukuda 2011 |
| Methods | Group1: knee group;<br>Group2: knee + hip group<br>Group3:no-treatment group |
| Recruitment of participants | Inclusion Criteria:<br>All the females included in this trial were sedentary, defined as individuals who had not practiced physical activity any day of the week, both aerobic and strengthening exercises, for at least the past 6 months.<br><br>Exclusion Criteria:<br>Females were excluded if they were pregnant or had any neurological disorders, hip or ankle injuries, low back or sacroiliac joint pain, rheumatoid arthritis, used corticosteroids and/or anti-inflammatory drugs, a heart condition that precluded performing the exercises, or previous surgery involving the lower extremities. We also excluded females who had other knee pathologies such as patellar instability, patellofemoral dysplasia, meniscal or ligament tears, osteoarthritis, tendinopathies, and epiphysitis. |
| Treatment | The females of the KE and KHE groups completed 3 treatment sessions per week for 4 weeks, totaling 12 sessions. The treatment for the individuals in the KE group emphasized stretching and strengthening of the knee musculature. Individuals in the KHE group were treated using the same protocol, with the addition of performing exercises to strengthen the hip abductor and lateral rotator muscles.<br><br>Group1: knee group<br>Stretching (HM, PF, quadriceps, and ITB), 3*30 s<br>Iliopsoas strengthening in non-weight bearing, 3*10 repetitions* |

| | |
|---|---|
| | Seated knee extension 90°-45°, 3 *10 repetitions*<br>Leg press 0°-45°, 3*10 repetitions*<br>Squatting 0°-45°, 3 *10 repetitions*<br><br>Group2: knee + hip group<br>Same protocol as the knee exercise group<br>Hip abduction against elastic band (standing), 3*10 repetitions*<br>Hip abduction with weights (sidelying), 3*10 repetitions†<br>Hip external rotation against elastic band (sitting), 3*10 repetitions*<br>Side-stepping against elastic band, 3*1 min |
| 41 | |
| Study | Theresa Helissa Nakagawa 2008 |
| Methods | Group1: knee exercise<br>Group2: hip + knee exercise |
| Recruitment of participants | The inclusion criteria were anterior or retropatellar knee pain during at least three of the following activities: ascending/descending stairs,<br>squatting, running, kneeling, hopping/jumping and prolonged sitting; the insidious onset of these symptoms being unrelated to a traumatic<br>incident and persistent for at least four weeks; and the presence of pain on palpation of the patellar facets, on stepping down from a 25-cm step, or during a double-legged squat.<br><br>The participants were excluded if they showed signs or symptoms of any of the following: meniscal or other intra-articular pathologic conditions; cruciate or collateral ligament involvement; tenderness over the patellar tendon, iliotibial band, or pesanserinus tendons; sign of patellar apprehension; Osgood-Schlatter or Sinding-Larsen-Johansson syndromes; hip or lumbar referred pain; a history of patellar dislocation; evidence of knee joint effusion; or previous surgery on the patellofemoral joint |
| Treatment | The exercise protocol for the control group consisted of patellar mobilization, stretching of the quadriceps, gastrocnemius, iliotibial band and hamstrings and open and closed kinetic chain exercises for quadriceps strengthening.<br>The intervention group received the same exercise protocol as the control group as well as additional time for strengthening and functional training exercises focused on the transversus abdominis muscle, hip abductors and lateral rotator muscles.<br>All the patients performed the rehabilitation exercises once a week under the supervision of the principal investigator and four times a week at home, for a total of five sessions a week for six weeks.<br><br>❖Intervention group:<br><br>Weeks 1 and 2 exercises:<br>●Transversus abdominis muscle contraction in the quadruped position,2 sets of 15 repetitions/10-second hold |

| | |
|---|---|
| | ●Isometric combined hip abduction–lateral rotation in sidelying with the hips and knees slightly flexed elastic resistance,2 sets of 15 repetitions/10-second hold<br>●Side-lying isometric hip abduction with extended knee,2 sets of 15 repetitions/10-second hold<br>●Isometric combined hip abduction–lateral rotation in the quadruped position,2 sets of 15 repetitions/10-second hold<br><br>Weeks 3 and 4 exercises:<br>●Pelvic drop exercise on a 20-cm step,2 sets of 15 repetitions/<br>10-second hold<br>●Upper extremity extension of the contralateral arm with elastic resistance performed in a single-leg stance,3 sets of 10 repetitions<br>●Rotation of the body in the direction of the contralateral side,<br>holding an elastic resistance with the ipsilateral arm while maintaining the lower extremity static,2 sets of 15 repetitions/10-second hold<br><br>Weeks 5 and 6 exercises, as for weeks 3 and 4<br>Additional elastic resistance around the affected leg in the forward lunges to encourage lateral rotation and abduction of the hip<br><br>❖Control group:<br><br>●Stretches (all exercise sessions),3 repetitions/30-second hold<br>Sitting hamstring stretch<br>Sitting patellar mobilization<br>Standing quadriceps stretch<br>Standing calf stretch<br>Standing iliotibial band stretch<br><br>Weeks 1 and 2 exercises:<br>●Isometric quadriceps contractions while sitting with 90 °of knee flexion,2 sets of 10 repetitions/10-second hold<br>●Straight-leg raise in supine position,3 sets of 10 repetitions<br>●Mini squats to 408 of knee flexion,4 sets of 10 repetitions<br><br>Weeks 3 and 4 exercises:<br>●Wall slides (0–60 of knee flexion),3 sets of 10 repetitions<br>●Steps-up and steps-down from a 20-cm step,3 sets of 5 repetitions<br>●Forward lunges (0–45 of knee flexion),3 sets of 10 repetitions<br><br>Weeks 5 and 6 exercises, as for weeks 3 and 4 plus:<br>●Balance exercises: unilateral stance on the floor and on an air-filled disc, with opened and closed eyes,3 sets of 30-second hold each exercise |

| | |
|---|---|
| | ●Progressive walking or running programme,3 sets of 30-second hold each exercise |
| 42 | |
| Study | Khalil Khayambashi 2012 |
| Methods | Randomized controlled trial<br>Group1: exercise group (n1=14)<br>Group2: no-exercise control group (n2=14) |
| Recruitment of participants | Included criteria:<br>To be considered for the study, patients had to be female and have a diagnosis of PFP. The diagnosis of PFP was based on the location of symptoms (peripatellar and/ or retropatellar) and the reproduction of pain with activities commonly association with this condition, such as stair descent, squatting, kneeling, and prolonged sitting. Patients were screened by physical examination to rule out ligamentous laxity, meniscal injury, pes anserine bursitis, iliotibial band syndrome, and patellar tendinitis as possible causes of current symptoms. Patients were invited to participate in the study if they had a diagnosis of bilateral PFP lasting at least 6 months (both knees), and had not previously received physical therapy.<br>Excluded criteria:<br>Patients were excluded from participation if they reported a history of previous patella dislocation, patellar fracture, or knee surgery. |
| Treatment | Twenty-eight females with PFP were sequentially assigned to an exercise (n = 14) or a no-exercise control group (n = 14). The exercise group completed bilateral hip abductor and external rotator strengthening 3 times per week for 8 weeks.<br>Group1:<br>The exercise group completed supervised hip-strengthening exercises 3 times per week for 8 weeks. Each session consisted of a 5-minute warm-up (walking around the gym at a self-selected pace), 20 minutes of hip-strengthening exercises, and a 5-minute cool-down (walking at a self-selected pace). All strengthening exercises were completed bilaterally. Each participant in the exercise group followed a standardized exercise program.<br>Group2:<br>As with the control group, individuals in the exercise group were asked to refrain from exercise-related activity beyond that of the supervised program and were allowed to take over-the-counter pain and/or anti-inflammatory medication as needed. |
| 43 | |
| Study | Chen-Yi Song 2009 |
| Methods | randomized controlled trial<br>Group1: hip adduction combined with leg-press exercise (LPHA group, n1=29)<br>Group2: leg-press exercise only (LP group, n2=30)<br>Group3:no exercise (control group, n3=30) |
| Recruitment of | Inclusion criteria |

| | |
|---|---|
| participants | (1) experience of anterior or retropatellar knee pain after performing at least 2 of the following activities: prolonged sitting, stair climbing, squatting, running, kneeling, hopping and jumping, and deep knee flexing; (2) insidious onset of symptoms unrelated to traumatic accident; (3) presence of pain for more than 1 month; and (4) age of 50 years and under (to eliminate the possibility of osteoarthritis). In addition, participants had to exhibit at least 2 of the following positive signs of anterior knee pain during the initial physical examination: (1) patellar crepitus, (2) pain following isometric quadriceps femoris muscle contraction against suprapatellar resistance with the knee in slight flexion (Clarke's sign), (3) pain following compression of the patella against the femoral condyles with the knee in full extension (patellar grind test), (4) tenderness upon palpation of the posterior surface of the patella or surrounding structures, and (5) pain following resisted knee extension.<br>Excluded criteria<br>(1) self-reported clinical evidence of other knee pathology; (2) patellar tendinitis or knee plica; (3) a history of knee surgery; (4) central or peripheral neurological pathology; (5) knee radiographic abnormalities (eg, knee osteoarthritis) or lower-extremity malalignment (eg, foot pronation); (6) severe knee pain (visual analog scale [VAS] score of > 8); or (7) received nonsteroidal anti-inflammatory drugs, injections, or physical therapy intervention in preceding 3 months. |
| Treatment | Participants were randomly assigned to the LP group, LPHA group, or control group and participated in 3 weekly exercise sessions for 8 weeks.<br>Simple LP exercise:<br>Leg-press exercise was performed unilaterally starting from 45 degrees of knee flexion to full extension using an ENDynamic Track machine. Exercise within the functional range was considered safe for patients with PFPS. A blue Thera-Band†was tied to each patient's thigh (without resistance) to maintain consistent tactile stimulation among groups. Prior to the beginning of exercise training, the unilateral 1-repetition maximum (RM) strength of the lower extremity was determined by Odvar Holten Pyramid diagram with repetition-tofatigue testing. Patients were unilaterally trained at 60% of 1 RM for 5 sets of 10 repetitions. The 1 RM was re-measured every 2 weeks, and the exercise intensity was adjusted accordingly. A 60-Hz metronome was used to control the exercise pace at 2-second concentric and eccentric contractions from 45 degrees of knee flexion to full extension. There were 2-second breaks between repetitions and 2-minute breaks between sets. Limbs were alternatively trained between exercise sets.<br>LPHA:<br>This exercise was performed as per the LP, except that a 50-N hip abduction force was applied to the distal one third of the thigh. This force was achieved by tying a blue Thera-Band to an arm of the EN-Dynamic Track machine (Fig. 2). Therefore, this exercise was a combination of LP and 50-N isometric hip adduction.<br>A hot pack was applied to the quadriceps femoris muscle for 15 minutes before |

| | |
|---|---|
| | exercise was commenced. After exercise completion, participants were asked to stretch the quadriceps, hamstring, iliotibial band, and calf muscle groups and were given a cold pack to apply for 10 minutes. Stretches were maintained for 30 seconds and were repeated 3 times for each muscle group. All study participants were asked not participate in any form of sport or exercise during the intervention period.<br>Control group：<br>Control group participants did not receive any exercise intervention, but were provided with health educational material regarding patellofemoral pain. They were advised not to perform or receive any exercise program or intervention. Neither tape nor brace was used. Exercise training was implemented after the 8-week control period. |
| 44 | |
| Study | Mahsa Emamvirdi 2019 |
| Methods | Group1： VCI exercise training<br>Group2： Written instructions + heat or ice treatment |
| Recruitment of participants | Inclusion criteria:<br>Patients were included in the study if they had anterior knee pain of 3 or greater on a 10-cm visual analog scale (VAS) for a minimum of 8 weeks before the assessment or anterior or retropatellar knee pain during at least 3 of the following activities: ascending/descending stairs, squatting, running, kneeling, jumping, and prolonged sitting. Patients also must have presented with an insidious onset of symptoms unrelated to trauma and positive Clark test.<br>Exclusion Criteria:<br>intra-articular pathology, patellar instability, Osgood-Schlatter or Sinding-Larsen-Johansson syndrome, hip pain, knee joint effusion, and previous surgery in the lower limb. Patients were also excluded if palpation of the patellar tendon, iliotibial band, or pes anserinus tendons reproduced the pain. |
| Treatment | Group1： VCI intervention<br>1.feedback methods and neuromuscular training：use verbal and visual (a mirror) feedback methods to control movement of the pelvis and the knee in the frontal plane. Feedback was eliminated during the last 4 sessions.<br>2.Each training session included 15 minutes of simple aerobic movements to warm up and cool down and about 45 minutes of prescribed exercise time. 3 times per week for 6 weeks, with at least 24 hours between intervention sessions.<br>3.The intensity of exercise was increased every 2 weeks. Each exercise was performed in 3 sets, and for the first week, each new exercise was repeated 6, 8, and 4 times.<br>Following exercise were done：<br>·Squat in front of mirror (0°-60° of knee flexion, performed in front of mirror to ensure the knee does not exceed the midfoot)<br>·Squat (0°-60° of knee flexion)<br>·Lateral walk with elastic resistance around the forefoot<br>·Strengthening the hip abductors with weightbearing (Trendelenburg) |

| | |
|---|---|
| | ·Squat with elastic resistance (0°-60° of knee flexion, resistance placed around the knees, stimulating the constant activation of the hip abductors and lateral rotators during task execution; relatively stable terrain)<br>·Squat on BOSU ball (BOSU) (0°-60° of knee flexion)<br>·Forward lunge in front of mirror (exercise performed in front of the mirror, single-leg balance at 30° of knee flexion on stable terrain)<br>·Forward lunge (single-leg balance at 30°of knee flexion on stable terrain)<br>·Balance exercise on BOSU ball<br>·Single-leg balance at 30° of knee flexion (performed on stable terrain)<br>·Squat with elastic resistance around the knees<br>·Unipodal squat on BOSU ball (keep the pelvis balanced and avoid excessive pronation of the foot)<br>·Modified forward lunge with elastic around the knee that is ahead (constant muscle activation of abductors and lateral rotators of the hip and training of motor control during the execution of the activity, performed on stable terrain)<br>·Romanian deadlift<br>·Lateral sliding without jumping<br>·Hip lateral rotation<br><br>Group2： control group<br>1.Received written instructions that included postural corrections and tips for improving general health.<br>2.They were asked to come to the clinic once or twice a week and received heat or ice treatment according to their needs. |
| 45 | |
| Study | Lori A Bolgla 2016 |
| Methods | Group1: hip/core rehabilitation program, n1=105;<br>Group2: knee rehabilitation program, n2=80 |
| Recruitment of participants | Inclusion criteria:<br>Briefly, subjects were recreationally-active (exercised a minimum of 30 minutes three times a week for at least 6 months prior) and between the ages of 18 and 35 years. Additional inclusion criteria were an insidious onset of PFP for at least 1 month, self-reported pain during activity of at least 3-cm on a 10-cm VAS, and pain during activities that required loading on a flexed knee (e.g., running, jumping, squatting, or stair ambulation).<br>Exclusion Criteria:<br>Exclusion criteria included a history of back or lower extremity pathology (including patella tendinopathy, patella instability, and/or iliotibial band stress syndrome) other than PFP. |
| Treatment | Group 1: The hip/core program + the rehabilitation specialist supervised exercises sessions<br>Group 2: The knee program + the rehabilitation specialist supervised exercises sessions<br>The rehabilitation specialist supervised exercises sessions： |

| | |
|---|---|
| | All subjects met with a trained rehabilitation specialist up to three times a week over a six-week period.<br>Subjects were instructed to perform the exercises at least six times a week (e.g.,a subject who attended three supervised sessions completed at least three additional sessions independently at home) and used Theraband® (The Hygenic Corp, Akron, OH) for resistance. Subjects performed all exercises bilaterally.<br><br>The hip/core program：<br>·non-weight bearing exercises.<br>·weight bearing exercises.<br>1 week:<br>Hip abduction-standing ； Hip external rotator-standing ； Hip external rotator-seated<br>2 week:<br>Hip abduction-standing ； Hip internalrotator-standing ； Hip external rotator-standing<br>3 week:<br>Hip abduction-standing; Hip internal rotator-standing; Hip external rotator-standing; Balancing 2 feet-Airex pad<br>4 week:<br>Hip extension@45°-standing; Hip internal rotator-standing; Hip external rotator-standing; Balancing 1 foot-Airex" pad<br><br>The knee program:<br>·non-weight bearing knee extensor exercises<br>·weight bearing.<br>1 week:<br>Isometric quadriceps setting; Knee extensions -standing; Double-legged, one-quarter squats<br>2 week:<br>Isometric quadriceps setting; Double-legged, one-half squats; Terminal knee extension with Theraband; Double-legged, one-quarter squats<br>3 week:<br>Double-legged, one-half squats; Single-legged, one-quarter squats Double-legged, one-quarter squats; Terminal-knee extension with Theraband.<br>4 week:<br>Single-legged, one-half squats; Forward, one-quarter lunges; Lateral step-down (4-in [3.6-cm] step); Forward step-down (4-in [3.6-cm] step) ; Double-legged, one-half wall squats<br>5-6 week:<br>Double-legged wall squats (to maximum 90°knee flexion); Lateral step-down (6-10-in [5.6-9.6-cm] step); Forward step-down (6-10-in [5.6-9.6-cm] step); Forward one-half full lunge (to maximum 90°knee flexion); Single-legged one-half full lunge (to maximum 90°knee flexion) |

### WEB APPENDIX 6. RISK OF BIAS FINDINGS

#### Contents:

**- Table 1. Domain-based risk of bias judgements for each outcome per study**
**- Table 2. Support for risk of bias judgement for each outcome per study**

**Table 1: domain-based risk of bias assessment**

| Study | Selection bias | | Performance bias | | Detection bias | Attrition bias | Reporting bias | Other bias |
|---|---|---|---|---|---|---|---|---|
| | Sequence generation | Allocation concealment | Participants | Personnel | | | | |
| Jean-Francois Esculier 2017 | Low risk | Low risk | High risk | High risk | Low risk | Low risk | High risk | Unclear risk |
| Jason Bonacci 2017 | Low risk | Low risk | Unclear risk | Unclear risk | Unclear risk | Low risk | Unclear risk | Unclear risk |
| Jenevieve L Roper 2016 | Low risk | Low risk | High risk | Unclear risk | Unclear risk | Low risk | Low risk | Unclear risk |
| Henrik Riel 2018 | Low risk | Low risk | Low risk | High risk | Unclear risk | Low risk | Low risk | Unclear risk |
| Selina L M Yip 2006 | Low risk | Unclear risk | Unclear risk | Unclear risk | Low risk | Low risk | Unclear risk | Unclear risk |
| N Dursun 2001 | Unclear risk | Unclear risk | Unclear risk | Unclear risk | Unclear risk | Unclear risk | Unclear risk | Unclear risk |
| Gustavo Telles 2016 | Low risk | Unclear risk | Unclear risk | Unclear risk | Unclear risk | Unclear risk | Unclear risk | Unclear risk |
| James W Brantingham 2009 | Low risk | Low risk | Unclear risk | Unclear risk | Low risk | High risk | Unclear risk | Unclear risk |
| Carsten M Mølgaard 2017 | Low risk | Low risk | High risk | High risk | Low risk | Low risk | Unclear risk | Unclear risk |
| Liliam B Priore 2019 | Low risk | Low risk | High risk | High risk | Low risk | Low risk | Unclear risk | Unclear risk |

| | | | | | | | | |
|---|---|---|---|---|---|---|---|---|
| Victor M Y Lun 2005 | Low risk | Low risk | Unclear risk | Low risk | Unclear risk | Low risk? | Unclear risk | Unclear risk |
| Mastour S Alshaharani 2019 | Unclear risk | Unclear risk | Unclear risk | Unclear risk | Unclear risk | Low risk | Unclear risk | Unclear risk |
| Serdar Demirci 2017 | Low risk | Unclear risk | Unclear risk | Unclear risk | Unclear risk | Unclear risk | Unclear risk | Unclear risk |
| Eda Akbaş 2011 | Low risk | Unclear risk | Unclear risk | Unclear risk | Low risk | Unclear risk | Unclear risk | Unclear risk |
| Lucas Simões Arrebola 2019 | Low risk | Low risk | Low risk | Low risk | Low risk | Low risk | Unclear risk | Unclear risk |
| Martin Whittingham 2004 | Low risk; | Low risk | Low risk | High risk | Low risk | Unclear risk | Unclear risk | Unclear risk |
| Jehoon Lee 2014 | Low risk | Unclear risk | Unclear risk | Unclear risk | Unclear risk | Unclear risk | Unclear risk | Unclear risk |
| Marjon Mason 2011 | Low risk | Low risk | Low risk | Low risk | Low risk | Unclear risk | Unclear risk | Unclear risk |
| Jyrki A Kettunen 2007 | Low risk | Low risk | Unclear risk | Unclear risk | High risk | Low risk | Unclear risk | Unclear risk |
| M S Rathleff 2015 | Low risk | Low risk | High risk | Low risk | Low risk | Unclear risk | Unclear risk | Unclear risk |
| Lachlan Giles 2017 | Low risk | Low risk | Low risk | Low risk | Unclear risk | Low risk | Unclear risk | Unclear risk |
| Angel Yañez-Álvarez 2020 | Low risk | Unclear risk | Unclear risk | Unclear risk | Low risk | Low risk | Unclear risk | Unclear risk |

| | | | | | | | | |
|---|---|---|---|---|---|---|---|---|
| Ebrahim Rasti 2020 | Low risk | Low risk | Low risk | Unclear | Low risk | Low risk | Unclear | Unclear risk |
| Mustafa Corum 2018 | Low risk | Unclear risk | Unclear risk | High risk | Low risk | Low risk | Unclear risk | Unclear risk |
| F Revelles Moyano 2012 | Low risk | Low risk | Unclear risk | Unclear risk | Low risk | Unclear risk | Unclear risk | Unclear risk |
| Nayra Deise Dos Anjos Rabelo 2017 | Low risk | Low risk | Low risk | High risk | Unclear risk | Low risk | Unclear risk | Unclear risk |
| Alireza Motealleh 2019 | Low risk | Low risk | Low risk | Unclear risk | Low risk | Low risk | Unclear risk | Unclear risk |
| Benjamin T Drew 2017 | Low risk | Low risk | Unclear risk | High risk | Low risk | Low risk | Unclear risk | Unclear risk |
| G Syme 2008 | Low risk | Low risk | High risk | Unclear risk | Low risk | Low risk | Unclear risk | Unclear risk |
| Farzin Halabchi 2015 | Low risk | Low risk | Unclear risk | Unclear risk | Unclear risk | Low risk | Unclear risk | Unclear risk |
| R van Linschoten 2009 | Low risk | Low risk | High risk | Unclear risk | Low risk | Low risk | Unclear risk | Unclear risk |
| Erik Witvrouw 2004 | Low risk | Low risk | Unclear risk | Unclear risk | Low risk | Low risk | Unclear risk | Unclear risk |
| Alexandra Hott 2020 | Low risk | Low risk | High risk | High risk | Low risk | Low risk | Low risk | Unclear risk |
| Marcelo Camargo Saad 2018 | Low risk | Low risk | Unclear risk | High risk | Low risk | Low risk | Low risk | High risk |
| Alexandra Hott | Low risk | Low risk | Low risk | High risk | Low risk | Low risk | Unclear risk | Low risk |

| | | | | | | | | |
|---|---|---|---|---|---|---|---|---|
| 2019 | | | | | | | | |
| Reed Ferber 2014 | Low risk | Unclear risk | Unclear risk | Unclear risk | Low risk | High risk | Unclear risk | Unclear risk |
| Kimberly L Dolak 2011 | Low risk | Low risk | Unclear risk | High risk | High risk | Unclear risk | Unclear risk | Unclear risk |
| Mehtap Şahin 2016 | Low risk | Low risk | High risk | Unclear risk | Unclear risk | Unclear risk | Unclear risk | Unclear risk |
| Thiago Yukio Fukuda 2012 | Low risk | Low risk | Unclear risk | Unclear risk | Low risk | Low risk | Unclear risk | Unclear risk |
| Thiago Yukio Fukuda 2011 | Low risk | Low risk | Unclear risk | Unclear risk | Low risk | Low risk | Unclear risk | Unclear risk |
| Theresa Helissa Nakagawa 2008 | Low risk | Low risk | Low risk | High risk | Low risk | Low risk | Unclear risk | Unclear risk |
| Khayambashi, Khalil 2012 | High risk | Unclear risk | Unclear risk | Unclear risk | Low risk | Low risk | Unclear risk | Unclear risk |
| Song, Chen-Yi 2009 | Low risk | Low risk | Unclear risk | Unclear risk | Low risk | Unclear risk | Unclear risk | Unclear risk |
| Emamvirdi, Mahsa 2019 | Low risk | Low risk | Low risk | Unclear risk | High risk | Low risk | Unclear risk | Unclear risk |
| Bolgla, Lori A 2016 | Low risk | Unclear risk | Low risk | Unclear risk | Low risk | Low risk | Unclear risk | Unclear risk |

**Table2: Risk of bias judgements + support for their judgements**

| Study: Jean-Francois Esculier 2017 | | |
|---|---|---|
| **Domain** | **Support for judgement** | **Review authors' judgement** |
| **Selection bias** | | |
| Random sequence generation | A scientist not involved in data collection generated randomisation lists using a random number generator (block randomisation; block size of 3–12). | Low risk |
| Allocation concealment | Group allocations were concealed in sequentially numbered sealed opaque envelopes, which were opened by one member of the research team not involved in data collection following baseline assessment. | Low risk |
| **Performance bias** | | |
| Blinging of participants and personnel | "A single-blind (evaluator only) parallel-group RCT was conducted." | High risk |
| **Detection bias** | | |
| Blinding of outcome assessment | "Participants were instructed not to reveal the content of their programme to the evaluator." | Low risk |
| **Attrition bias** | | |
| Incomplete outcome date | "Seven participants dropped out of the study before week 8 (follow-up rate=89.9%), and three additiona,l runners failed to return their follow-up questionnaires at week 20 (follow-up rate=85.5%)."At 20-week, the numbers of the three groups were20/23; 21/23; 18/23. Intention-to-treat and per-protocol analyses were used for symptoms and function outcomes. | Low risk |
| **Reporting bias** | | |
| Selective reporting | "Trial registration number ClinicalTrials. gov (NCT02352909). secondary outcome Global rating of change (GRC) were not reported." | High risk |
| **Other bias** | | |

| | | |
|---|---|---|
| Other sources of bias | Unclear, insufficient information. | Unclear risk |

| **Study: Jason Bonacci 2017** | | |
|---|---|---|
| **Domain** | **Support for judgement** | **Review authors' judgement** |
| **Selection bias** | | |
| Random sequence generation | "Allocation was done according to a computer-generated randomisation schedule. | Low risk |
| Allocation concealment | Group allocation was sealed in opaque, consecutively numbered envelopes by an independent researcher and stored in a central locked location. " | Low risk |
| **Performance bias** | | |
| Blinging of participants and personnel | Unclear, no information. | Unclear risk |
| **Detection bias** | | |
| Blinding of outcome assessment | Unclear, no information. | Unclear risk |
| **Attrition bias** | | |
| Incomplete outcome date | "Sixteen participants were randomised and 14 (7 (88%) gait retraining and 7 (88%) foot orthoses) completed follow-up assessment. One participant in the gait retraining group withdrew due to an ankle sprain and one participant in the foot orthoses group was no longer interested.Neither of the participants who withdrew had started treatment. " | Low risk |
| **Reporting bias** | | |
| Selective reporting | Unclear, insufficient information. | Unclear risk |
| **Other bias** | | |
| Other sources of bias | Unclear, insufficient information | Unclear risk |

| Study: Jenevieve L Roper 2016 | | |
|---|---|---|
| **Domain** | **Support for judgement** | **Review authors' judgement** |
| **Selection bias** | | |
| Random sequence generation | "Protocol Research personnel randomized subjects using a random numbers generator to either the control group or experimental group using blocked randomization so that there were an equal number (n=8) of subjects in each group. " | Low risk |
| Allocation concealment | "The sequence was kept in a locked filing cabinet in the Gait Analysis lab until interventions were assigned. " | Low risk |
| **Performance bias** | | |
| Blinging of participants and personnel | "Protocol Research personnel randomized subjects to either the control group or experimental group, subjects were not blinded to what group they were in." | High risk |
| **Detection bias** | | |
| Blinding of outcome assessment | Unclear, insuffcient information. | Unclear risk |
| **Attrition bias** | | |
| Incomplete outcome date | "All randomized subjects completed all trials. Analyses were performed on all subjects in both the experimental (n=8) and control (n=8) groups." | Low risk |
| **Reporting bias** | | |
| Selective reporting | This trial was registered at the US National Institutes of Health (Clinicaltrials.gov) #NCT02567123. | Low risk |
| **Other bias** | | |
| Other sources of bias | Unclear, insufficient information. | Unclear risk |

| Study: Henrik Riel 2018 | | |
|---|---|---|
| **Domain** | **Support for judgement** | **Review authors' judgement** |
| **Selection bias** | | |
| Random sequence generation | "Randomisation Adolescents were block randomised using a random number generator on www.random.org. A researcher not involved in the data collection or analysis generated the allocation sequence and was the only person who knew the block sizes. " | Low risk |
| Allocation concealment | "After all baseline measurements were made, the assessor took a sequentially numbered opaque sealed envelope in which allocation was indicated." | Low risk |
| **Performance bias** | | |
| Blinging of participants and personnel | "Participant-blinded. The primary investigator was not blinded towards group allocation. The adolescents attended separate group training sessions based on their randomisation and had no contact with the participants of the opposite group." | Low risk<br>High risk |
| **Detection bias** | | |
| Blinding of outcome assessment | Unclear, insufficient information. | Unclear risk |
| **Attrition bias** | | |
| Incomplete outcome date | "Two adolescents failed to participate in the follow-up. Statistical analysis the primary intention-to-treat analysis tested the between-group difference of mean deviation from the prescribed contraction time per repetition using an independent t-test." | Low risk |
| **Reporting bias** | | |

| | | |
|---|---|---|
| Selective reporting | superiority trial (NCT02674841) with a 2-group parallel design conducted in Aalborg, Denmark.Recruitment The trial was approved by the Ethics committee of North Denmark Region (project ID: N20150070). | Low risk |
| **Other bias** | | |
| Other sources of bias | Unclear, insufficient information. | Unclear risk |

| **Study: Selina L M Yip 2006** | | |
|---|---|---|
| **Domain** | **Support for judgement** | **Review authors' judgement** |
| **Selection bias** | | |
| Random sequence generation | "The subjects were then randomly assigned into two groups by drawing lots." | Low risk |
| Allocation concealment | Unclear, insufficient information. | Unclear risk |
| **Performance bias** | | |
| Blinging of participants and personnel | "This study involved repeated measurements with a double-blinded design." Insufficient information about the blinging of participants and personnel. | Unclear risk |
| **Detection bias** | | |
| Blinding of outcome assessment | "the assessor was blinded to the subject grouping throughout the whole study." | Low risk |
| **Attrition bias** | | |
| Incomplete outcome date | "All subjects completed the home programme and the assessment sessions.Assessments were performed at weeks 0, 4 and 8, respectively . A study period of eight weeks was chosen so as to study the effect of a training programme." | Low risk |

| | | |
|---|---|---|
| **Reporting bias** | | |
| Selective reporting | Unclear, insufficient information. | Unclear risk |
| **Other bias** | | |
| Other sources of bias | Unclear, insufficient information. | Unclear risk |

| **Study: N Dursun 2001** | | |
|---|---|---|
| **Domain** | Support for judgement | Review authors' judgement |
| **Selection bias** | | |
| Random sequence generation | "Patients were randomized to biofeedback and control groups, each consisting of 30 patients." | Unclear risk |
| Allocation concealment | Unclear, insufficient information. | Unclear risk |
| **Performance bias** | | |
| Blinging of participants and personnel | Unclear the blinging of participants.The conventional exercise program for both groups was supervised by the same physical therapist | Unclear risk |
| **Detection bias** | | |
| Blinding of outcome assessment | Unclear, insufficient information. | Unclear risk |
| **Attrition bias** | | |
| Incomplete outcome date | "No dropouts occurred during the trial and all subjects in both groups regularly participated in the treatment."<br>No sufficient information about follow-up. | Unclear risk |
| **Reporting bias** | | |
| Selective reporting | Unclear, insufficient information. | Unclear risk |
| **Other bias** | | |

| | | |
|---|---|---|
| Other sources of bias | Unclear, insufficient information. | Unclear risk |

| **Study: Gustavo Telles 2016** | | |
|---|---|---|
| **Domain** | **Support for judgement** | **Review authors' judgement** |
| **Selection bias** | | |
| Random sequence generation | "Randomization was performed electronically on http://graphpad.com/quickcalcs/index.cfm using simple random sampling." | Low risk |
| Allocation concealment | "A physiotherapist was responsible for the screening of eligible patients and the random allocation of participants." | Unclear risk |
| **Performance bias** | | |
| Blinging of participants and personnel | "……participants were referred for an initial evaluation consisting of medical history and physical examination performed by a blinded physiotherapist." No sufficient information about blinging of participants. | Unclear risk |
| **Detection bias** | | |
| Blinding of outcome assessment | Unclear, insufficient information. | Unclear risk |
| **Attrition bias** | | |
| Incomplete outcome date | "22 met the eligibility criteria of the study after the evaluation. Among these, four participants were excluded because they started the treatment and then dropped out. All 18 subjects who continued in the study." No information on reasons for loss to follow-up. | Unclear risk |
| **Reporting bias** | | |
| Selective reporting | No protocol/analysis plan could be retrieved in trial registers. | Unclear risk |
| **Other bias** | | |
| Other sources of bias | Unclear, insufficient information. | Unclear risk |

| **Study: James W Brantingham 2009** | | |
|---|---|---|
| **Domain** | **Support for judgement** | **Review authors' judgement** |
| **Selection bias** | | |
| Random sequence generation | "A computer-generated list of random numbers was allocated into either group "A" or "B." | Low risk |
| Allocation concealment | "These letters were written on slips of paper, folded over, and sealed in envelopes that had no marking on them on the outside and the folded numbers inside were undetectable. These envelopes placed in a locked cabinet were requested for assignment only after a patient was fully accepted into the study and given out by a researcher who had no contact with the patients." | Low risk |
| **Performance bias** | | |
| Blinging of participants and personnel | **Unclear, insufficient information.** | **Unclear risk** |
| **Detection bias** | | |
| Blinding of outcome assessment | "If the blind assessor became aware of the subject's group assignment (ie, became unblinded), the assessor was to, without delay, leave the room and immediately report to the project coordinator/or another supervising clinical faculty member. Another blind assessor was then located to make the assessment." | Low risk |
| **Attrition bias** | | |
| Incomplete outcome date | A total of 47 people was included, including 25 in group A and 22 in group | High risk |

| | | |
|---|---|---|
| | B. In the end, 22 people in group A received the intervention, and 21 people in group B received the intervention. Finally, 13 people in group A and 18 people in group B were followed up. | |
| **Reporting bias** | | |
| Selective reporting | Unclear, insufficient information. | Unclear risk |
| **Other bias** | | |
| Other sources of bias | Unclear, insufficient information. | Unclear risk |

| **Study: Carsten M Mølgaard 2017** | | |
|---|---|---|
| **Domain** | **Support for judgement** | **Review authors' judgement** |
| **Selection bias** | | |
| Random sequence generation | "The randomisation was managed by an independent secretary not involved in assessment of participants, who generated a simple randomisation sequence a priori. " | Low risk |
| Allocation concealment | "Allocation was sealed in opaque and consecutively numbered envelopes held in a central location. Envelopes were opened in sequence by a person not involved in the study after recruitment and baseline testing of participants." | Low risk |
| **Performance bias** | | |
| Blinging of participants and personnel | "The participants and the physiotherapists responsible for delivering the interventions were not blinded." | High risk |
| **Detection bias** | | |
| Blinding of outcome assessment | "Assessor-blinded. The physiotherapist, responsible for collecting outcome measures at follow-up was blinded to the randomisation." | Low risk |

| Attrition bias | | |
|---|---|---|
| Incomplete outcome date | “Between-group comparison was analyzed on an intention-to-treat basis and included all individuals who responded to the self-report questionnaire at that time point.” | Low risk |
| Selective reporting | Unclear, insufficient information. | Unclear risk |
| **Other bias** | | |
| Other sources of bias | Unclear, insufficient information. | Unclear risk |

| **Study: Liliam B Priore 2019** | | |
|---|---|---|
| **Domain** | **Support for judgement** | **Review authors’ judgement** |
| **Selection bias** | | |
| Random sequence generation | “Randomization codes were generated in blocks of 4 and 6 using a custom list at the website (https://www.sealedenvelope.com/).” | Low risk |
| Allocation concealment | “Sealed opaque envelopes, sequentially numbered, were used to hide the allocation. ” | Low risk |
| **Performance bias** | | |
| Blinging of participants and personnel | “As the participants were informed about the type of intervention they were receiving, the study was not considered double-blind.” | High risk |
| **Detection bias** | | |
| Blinding of outcome assessment | “The assessments were performed by a third investigator who was blinded to group allocation.” | Low risk |
| **Attrition bias** | | |
| Incomplete outcome date | “Intention-to-treat analyses were used for all outcomes.” | Low risk |

| Reporting bias | | |
|---|---|---|
| Selective reporting | Trial Registration: ensaiosclinicos.gov.br/: RBR-2DY25R.<br>Unclear. No protocol/analysis plan could be retrieved in trial registers | Unclear risk |
| **Other bias** | | |
| Other sources of bias | Unclear, insufficient information. | Unclear risk |

| **Study: Victor M Y Lun 2005** | | |
|---|---|---|
| **Domain** | **Support for judgement** | **Review authors' judgement** |
| **Selection bias** | | |
| Random sequence generation | "a random number generator with block design" | Low risk |
| Allocation concealment | "a second research assistant used a random number generator with block design to assign subjects to 1 of 4 treatment groups" | Low risk |
| **Performance bias** | | |
| Blinging of participants and personnel | "The investigators were blinded to the treatment group of each subject." | Low risk |
| **Detection bias** | | |
| Blinding of outcome assessment | Insufficient information:<br>"The study coordinator then obtained background demographic data and measured subject. Baseline assessment of the study's outcome measurements was then performed."<br>"These journals were submitted to the second research assistant on a monthly basis." | Unclear risk |
| **Attrition bias** | | |
| Incomplete outcome date | Reasons for missing outcome data unlikely to be related to true outcome: | Low risk |

| | | |
|---|---|---|
| | "Subjects who withdrew from the study were not followed (non–intention-to-treat analysis)."<br>"Three hundred five subjects were initially screened for the study. Of these, 152 met the study inclusion criteria. Twenty-one subjects withdrew from the study because of a job transfer or lack of interest. Two subjects were crossed over to another treatment group before 3 months and considered to be withdrawals from the study."<br>"Therefore, a total of 129 subjects (76 females and 53 males with a total of 186 affected knees; mean age, 35; range, 18–60) were included in the final analysis (Fig. 1). A summary of the baseline characteristics of these 129 subjects, within their respective treatment group, is seen in Table 2. It can be seen that subjects in each treatment group were very similar with respect to age, height, weight, leg length difference, Q angle, affect side, and duration of symptoms." | |
| **Reporting bias** | | |
| Selective reporting | Insufficient information | Unclear risk |
| **Other bias** | | |
| Other sources of bias | Insufficient information | Unclear risk |

| **Study: Mastour S Alshaharani 2019** | | |
|---|---|---|
| **Domain** | **Support for judgement** | **Review authors' judgement** |
| **Selection bias** | | |
| Random sequence generation | There is not enough information to determine whether a random sequence is at high or low risk. | Unclear risk |

| | | |
|---|---|---|
| Allocation concealment | There is not enough information. | Unclear risk |
| **Performance bias** | | |
| Blinging of participants and personnel | This was not described in the study. | Unclear risk |
| **Detection bias** | | |
| Blinding of outcome assessment | This was not described in the study. | Unclear risk |
| **Attrition bias** | | |
| Incomplete outcome date | The generation of missing data is unlikely to be related to the true outcome. "One subject was excluded as they had a meniscus lesion that prohibited them from participating in the study, and another subject voluntarily left the study due to personal time constraints, therefore only data from the 41 remaining subjects who completed the prescribed intervention was analyzed." | Low risk |
| **Reporting bias** | | |
| Selective reporting | No protocol/analysis plan could be retrieved in trial registers. | Unclear risk |
| **Other bias** | | |
| Other sources of bias | There is insufficient information to assess whether there are other significant risks of bias. | Unclear risk |

| **Study: Serdar Demirci 2017** | | |
|---|---|---|
| **Domain** | **Support for judgement** | **Review authors' judgement** |
| **Selection bias** | | |
| Random sequence generation | "a computer-generated randomization" | Low risk |

| | | |
|---|---|---|
| Allocation concealment | Insufficient information | Unclear risk |
| **Performance bias** | | |
| Blinging of participants and personnel | Insufficient information | Unclear risk |
| **Detection bias** | | |
| Blinding of outcome assessment | Insufficient information | Unclear risk |
| **Attrition bias** | | |
| Incomplete outcome date | Insufficient information | Unclear risk |
| **Reporting bias** | | |
| Selective reporting | Insufficient information | Unclear risk |
| **Other bias** | | |
| Other sources of bias | Insufficient information | Unclear risk |

| **Study: Eda Akbaş 2017** | | |
|---|---|---|
| **Domain** | **Support for judgement** | **Review authors' judgement** |
| **Selection bias** | | |
| Random sequence generation | "Using a random number generator." | Low risk |
| Allocation concealment | Insufficient information | Unclear risk |
| **Performance bias** | | |
| Blinging of participants and personnel | Insufficient information to determine low or high risk. | Unclear risk |
| **Detection bias** | | |
| Blinding of outcome assessment | "Evaluations were performed by two experienced physiotherapists. | Low risk |

| | | |
|---|---|---|
| | Examiner 1 was blinded and positioned the patient. While taking the measurements, it was not possible for Examiner 2 to remain blinded. ... There are some limitations to this study. As in other researches in this area, the examiners were not blinded to the participants' group status. However, with the aim of reducing any bias this might cause, Examiner 1 was blinded. The subjects were measured without warm-up or pre-stretching, which may have affected flexibility. However, as this was standardized, any effect would be spread across all participants." | |
| **Attrition bias** | | |
| Incomplete outcome date | Insufficient information | Unclear risk |
| **Reporting bias** | | |
| Selective reporting | Insufficient information | Unclear risk |
| **Other bias** | | |
| Other sources of bias | Insufficient information | Unclear risk |

| **Study: Lucas Simões Arrebola 2019** | | |
|---|---|---|
| **Domain** | **Support for judgement** | **Review authors' judgement** |
| **Selection bias** | | |
| Random sequence generation | "A draw of opaque and sealed envelopes containing a group number." | Low risk |
| Allocation concealment | "The participants were randomized into three groups by a draw of opaque and sealed envelopes containing a group number." | Low risk |
| **Performance bias** | | |
| Blinging of participants and personnel | "Design: Double-blind, randomized, controlled pilot study." | Low risk |

| | | |
|---|---|---|
| **Detection bias** | | |
| Blinding of outcome assessment | "all the evaluations were conducted with the participant wearing a long T-shirt, opaque gym pants, 3/4-length stockings or gaiters, and sneakers. The clothing was selected to cover the entire trunk and lower limb region, thereby making it impossible for the physiotherapist to see the participants KT® placement."<br><br>"The evaluations were performed by a physiotherapist blinded to the randomization." | Low risk |
| **Attrition bias** | | |
| Incomplete outcome date | "During the treatment protocol, 21% of the sample dropped out after 6 weeks and 35% did not attend the final assessment at 12 weeks. Also, 63% did not attend the 12-weeks follow-up. We hypothesized that the high drop-out rate was related to the improvement of pain and function observed at 6 weeks. Also, these patients had a low socioeconomic condition and could not afford the transportation to the ambulatory clinic where the study took place, through the 12 weeks." | Low risk |
| **Reporting bias** | | |
| Selective reporting | Insufficient information | Unclear risk |
| **Other bias** | | |
| Other sources of bias | "Other limitations to this pilot study include the fact that the first evaluation after baseline was performed late, the fact that a PROM and a quality of life evaluation were not performed, and the fact that only women were recruited. Perhaps the KT method® has a greater short-term effect on decreasing pain and improving function compared to the methods | Unclear risk |

| | |
|---|---|
| | administered to the CG (Morris et al., 2013; Freedman et al., 2014) that were not detected on a late 6-week evaluation. In the full RCT, weekly NPRS at rest and during effort measurements will be performed to detect this short-term effect, and the PROM and the brief World Health Organization Quality of Life Assessment (WHOQOL-BREF) (The WHOQOL Group, 1998) will be added to the outcome measures. The reason for only including women is related to the higher incidence of PFPS in the female sex (Rothermich et al., 2015). ” |

| **Study: Martin Whittingham 2004** | | |
|---|---|---|
| **Domain** | **Support for judgement** | **Review authors' judgement** |
| **Selection bias** | | |
| Random sequence generation | "3 labeled envelopes." | Low risk |
| Allocation concealment | "A block randomization process was used, where subjects randomly chose 1 of 3 labeled envelopes to determine their group allocation. The next subject chose 1 of the remaining 2 envelopes and the third subject was then assigned to the remaining group before the process was repeated." | Low risk |
| **Performance bias** | | |
| Blinging of participants and personnel | "All subjects remained in the group to which they were originally assigned. All subjects were placed on restricted duties (similar for all individuals) throughout the treatment period."<br><br>Personnels were not blind:<br>"A second physiotherapist (therapist 2), who was aware of group | Low risk<br><br>High risk |

| | | |
|---|---|---|
| | allocation, applied adhesive tape to the affected knee of subjects in the taping group and placebo taping group." | |
| **Detection bias** | | |
| Blinding of outcome assessment | "Another physiotherapist (therapist 3), who was blinded to group allocation, took all outcome measures. Subjects wore tracksuit bottoms to hide the presence of tape during assessment."<br>"Outcome measures were visual analog scales for pain and the functional index questionnaire, recorded at weekly intervals by a therapist who was blinded to group allocation." | Low risk |
| **Attrition bias** | | |
| Incomplete outcome date | Insufficient information | Unclear risk |
| **Reporting bias** | | |
| Selective reporting | Insufficient information | Unclear risk |
| **Other bias** | | |
| Other sources of bias | "There was also a limited age range and the study sample had a majority of men. The external validity of the findings should, therefore, be considered within these limitations. Outcomes were taken over a 4-week period in the present investigation, so it is not possible to comment on the long-term success of treatment." | Unclear risk |

| **Study: Jehoon Lee 2014** | | |
|---|---|---|
| **Domain** | **Support for judgement** | **Review authors' judgement** |
| **Selection bias** | | |
| Random sequence generation | "A computer using a basic random number generator." | Low risk |

| | | |
|---|---|---|
| Allocation concealment | Insufficient information to determine low or high risk. | Unclear risk |
| **Performance bias** | | |
| Blinging of participants and personnel | Insufficient information to determine low or high risk. | Unclear risk |
| **Detection bias** | | |
| Blinding of outcome assessment | Insufficient information to determine low or high risk. | Unclear risk |
| **Attrition bias** | | |
| Incomplete outcome date | Insufficient information to determine low or high risk. | Unclear risk |
| **Reporting bias** | | |
| Selective reporting | Insufficient information to determine low or high risk. | Unclear risk |
| **Other bias** | | |
| Other sources of bias | Insufficient information to determine low or high risk. | Unclear risk |

| **Study: Marjon Mason 2011** | | |
|---|---|---|
| **Domain** | **Support for judgement** | **Review authors' judgement** |
| **Selection bias** | | |
| Random sequence generation | "a selected, sealed and prenumbered envelope" | Low risk |
| Allocation concealment | "The latter was blind to the treatment grouping of the subjects who were randomly allocated to one of four groups according to a selected, sealed and prenumbered envelope." | Low risk |
| **Performance bias** | | |
| Blinging of participants and personnel | "A prospective double-blind randomized control study was designed"<br>"Two physiotherapists were involved in this study, namely a treating and | Low risk |

| | | |
|---|---|---|
| | an assessing physiotherapist. The latter was blind to the treatment grouping of the subjects” | |
| **Detection bias** | | |
| Blinding of outcome assessment | “All subjects were asked to wear long pants for the end of the first week assessment so that the assessing physiotherapist remained blinded towards the single modality treatment during that week.”<br>“the assessor was blind to the subject groupings.” | Low risk |
| **Attrition bias** | | |
| Incomplete outcome date | Insufficient information | Unclear risk |
| **Reporting bias** | | |
| Selective reporting | Insufficient information | Unclear risk |
| **Other bias** | | |
| Other sources of bias | Insufficient information | Unclear risk |

| **Study: Jyrki A Kettunen 2007** | | |
|---|---|---|
| **Domain** | **Support for judgement** | **Review authors’ judgement** |
| **Selection bias** | | |
| Random sequence generation | “The randomization process was carried out using a computer-generated randomization list stratified by gender.” | Low risk |
| Allocation concealment | “Sealed, sequentially numbered envelopes containing information on the treatment group were prepared and given to the assisting nurse, who opened the envelopes in numerical order after recruitment so that concealment of allocation was successful in all cases.” | Low risk |
| **Performance bias** | | |

| | | |
|---|---|---|
| Blinging of participants and personnel | Insufficient information | Unclear risk |
| **Detection bias** | | |
| Blinding of outcome assessment | The data collector was not blinded:<br>"Outcome measures were collected using self-administered questionnaires. This data collection was organized by the study coordinator (JAK). As the coordinator did not have any presuppositions as to which of the groups would show better results and because he was not a treatment provider, the data collector was not, for practical reasons, blinded to the treatment groups." | High risk |
| **Attrition bias** | | |
| Incomplete outcome date | "In addition, we carried out 'a worst-case scenario' analysis of the data. In this analysis, we assumed that the Kujala score would have been the same as the baseline score (no change), if follow-up data were not available."<br>"One patient in the arthroscopy group and three in the control group were lost to the follow-up."<br>Figure 1 illustrates the details of lost follow-up. | Low risk |
| **Reporting bias** | | |
| Selective reporting | Insufficient information | Unclear risk |
| **Other bias** | | |
| Other sources of bias | Insufficient information | Unclear risk |

| **Study: M S Rathleff 2015** | | |
|---|---|---|
| **Domain** | **Support for judgement** | **Review authors' judgement** |

| | | |
|---|---|---|
| **Selection bias** | | |
| Random sequence generation | "computer-generated sequence developed by the main investigator (MSR)." | Low risk |
| Allocation concealment | "The four schools were randomised either to patient education or patient education and exercise therapy using a computer-generated sequence developed by MSR. Cluster randomisation was chosen to minimise the contamination between individuals, which could occur if more than one adolescent in each class were diagnosed with PFP, but randomised to different treatment groups." | Low risk |
| **Performance bias** | | |
| Blinging of participants and personnel | Participants were blinded:<br>"Cluster randomisation was chosen to minimise the contamination between individuals, which could occur if more than one adolescent in each class were diagnosed with PFP, but randomised to different treatment groups."<br><br>Physiotherapists were blinded:<br>"One physiotherapist delivered the patient education in the two clusters randomised to patient education alone."<br>"One of two physiotherapists delivered the exercise therapy and patient education in each cluster." | High risk<br><br><br><br><br>Low risk |
| **Detection bias** | | |
| Blinding of outcome assessment | "Self-report questionnaires were completed at baseline, 3, 6, 12 and 24 months after inclusion and collected by blinded project personnel."<br>"The first author and a statistician not involved in the study performed all analyses. They were not blinded to group allocation during the analyses." | Low risk |
| **Attrition bias** | | |
| Incomplete outcome date | "Follow-up rate ranged from 73% to 91% with a 91% follow-up rate at the | Unclear risk |

| | | |
|---|---|---|
| | primary endpoint at 12 months."<br>Appendix 2a illustrates the number of adolescents available for follow-up at each time-point at the four clusters. But the reason is not given for the loss of follow-up. | |
| **Reporting bias** | | |
| Selective reporting | Insufficient information | Unclear risk |
| **Other bias** | | |
| Other sources of bias | Insufficient information | Unclear risk |

| **Study: Lachlan Giles 2017** | | |
|---|---|---|
| **Domain** | **Support for judgement** | **Review authors' judgement** |
| **Selection bias** | | |
| Random sequence generation | The randomisation was performed by a person independent to the study in lots of 20 at a 1:1 ratio by drawing group allocation from a concealed box; the box was replenished before each lot had been used. | Low risk |
| Allocation concealment | Assessments conducted by a physiotherapist blinded to treatment allocation. | Low risk |
| **Performance bias** | | |
| Blinging of participants and personnel | Participants conducted by a physiotherapist blinded to treatment allocation. Participants were blinded to group allocation, and both groups were informed they were receiving BFR. | Low risk |
| **Detection bias** | | |
| Blinding of outcome assessment | unknown | Unclear risk |
| **Attrition bias** | | |

| | | |
|---|---|---|
| Incomplete outcome date | 87%participants all of whom completed follow-up at 6 months. Of the non-completers. The last reported scores of the non-completers were carried forward. | Low risk |
| **Reporting bias** | | |
| Selective reporting | This randomised controlled trial was registered with the Australian New Zealand Clinical Trials Registry Trial Number 12614001164684 | Unclear risk |
| **Other bias** | | |
| Other sources of bias | unknown | Unclear risk |

| **Study: Angel Yañez-Álvarez 2020** | | |
|---|---|---|
| **Domain** | **Support for judgement** | **Review authors' judgement** |
| **Selection bias** | | |
| Random sequence generation | Those participants who met the inclusion criteria were randomly assigned by a member of the research team to the experimental or control group following simple randomization procedures using a random-number generator website http://www.randomization.com, and considering a 1:1 ratio distribution of participants in the study groups" | Low risk |
| Allocation concealment | Unknow | Unclear risk |
| **Performance bias** | | |
| Blinging of participants and personnel | Unknow | Unclear risk |
| **Detection bias** | | |
| Blinding of outcome assessment | This study was a single-blind prospective randomized controlled clinical trial. A blind evaluator performed all measurements. | Low risk |

| | | |
|---|---|---|
| **Attrition bias** | | |
| Incomplete outcome date | All patients who received the intervention were followed up | Low risk |
| **Reporting bias** | | |
| Selective reporting | Trial registration: ClinicalTrials.gov (NCT04031248). | Unclear risk |
| **Other bias** | | |
| Other sources of bias | unknown | Unclear risk |

| **Study: Ebrahim Rasti 2020** | | |
|---|---|---|
| **Domain** | **Support for judgement** | **Review authors' judgement** |
| **Selection bias** | | |
| Random sequence generation | Then an online randomization application (www.randomization.com) was used to randomly assign the patients to two group. | Low risk |
| Allocation concealment | For group concealment we used a sealed envelope. | Low risk |
| **Performance bias** | | |
| Blinging of participants and personnel | Patients were aware of the existence of two different groups, but did not know whether they had been assigned to the treatment or control group. | Low risk |
| **Detection bias** | | |
| Blinding of outcome assessment | The outcome assessor was unaware of the group allocation of the participants. | Low risk |
| **Attrition bias** | | |
| Incomplete outcome date | All patients who received the intervention were analyzed. | Low risk |
| **Reporting bias** | | |
| Selective reporting | this research was registered in the Iranian Registry of Clinical Trials | Unclear |

| | | |
|---|---|---|
| | (IRCT20090831002391N39). | |
| **Other bias** | | |
| Other sources of bias | unknown | Unclear risk |

| **Study: Mustafa Corum 2018** | | |
|---|---|---|
| **Domain** | **Support for judgement** | **Review authors' judgement** |
| **Selection bias** | | |
| Random sequence generation | Patients were randomly assigned equally to a WBV plus home exercise (intervention) group or a home exercise only (control) group using a computer-generated programme. | Low risk |
| Allocation concealment | unknown | Unclear risk |
| **Performance bias** | | |
| Blinging of participants and personnel | Insufficient information<br><br>"informed the non-blinded administering physician (C. Basoglu) about the allocation of the study patients to the WBV or control groups." | Unclear risk<br><br>High risk |
| **Detection bias** | | |
| Blinding of outcome assessment | All assessments and data collection were performed by a single assessor (M. Corum) who was blind to the groups. | Low risk |
| **Attrition bias** | | |
| Incomplete outcome date | The final study sample consisted of 34 patients (n=18 in the WBV and n=16 in the control groups). 6 patients were excluded from the final analysis due to attendance failure. An intention-to-treat analysis was not performed because the dropout patients were not included in the statistical | Low risk |

| | | |
|---|---|---|
| | analysis. | |
| **Reporting bias** | | |
| Selective reporting | unknown | Unclear risk |
| **Other bias** | | |
| Other sources of bias | unknown | Unclear risk |

| **Study: F Revelles Moyano 2012** | | |
|---|---|---|
| **Domain** | **Support for judgement** | **Review authors' judgement** |
| **Selection bias** | | |
| Random sequence generation | The randomization sequence was drawn up and kept off-site by an independent body, using a random number generator in blocks of eight with no stratification. | Low risk |
| Allocation concealment | To ensure concealment of allocation, eligibility was determined by a blinded assessor not involved in the randomization process. | Low risk |
| **Performance bias** | | |
| Blinging of participants and personnel | unknown | Unclear risk |
| **Detection bias** | | |
| Blinding of outcome assessment | "To ensure concealment of allocation, eligibility was determined by a blinded assessor not involved in the randomization process" | Low risk |
| **Attrition bias** | | |
| Incomplete outcome date | a total of 74 patients were enrolled in the study and 2 patients lost to follow up. Lost to follow-up（N=2）， No specific reason | Unclear risk |
| **Reporting bias** | | |

| | | |
|---|---|---|
| Selective reporting | unknown | Unclear risk |
| **Other bias** | | |
| Other sources of bias | unknown | Unclear risk |

| **Study: Nayra Deise Dos Anjos Rabelo 2017** | | |
|---|---|---|
| **Domain** | **Support for judgement** | **Review authors' judgement** |
| **Selection bias** | | |
| Random sequence generation | The randomization codes were generated using the RAND function of Excel. | Low risk |
| Allocation concealment | Patients were randomized to the two groups using opaque, sealed, sequentially numbered envelopes. | Low risk |
| **Performance bias** | | |
| Blinging of participants and personnel | "Participants were informed that they would receive one of two different forms of<br>treatment but were unaware of the exercises performed by the other group.<br><br>Due to the nature of the interventions, it was not possible to blind the physiotherapists who carried out the interventions." One of the two therapists, who carried out the treatment, opened the envelopes with the random codes on the first day of treatment. | Low risk<br><br>High risk |
| **Detection bias** | | |
| Blinding of outcome assessment | unknown | Unclear risk |
| **Attrition bias** | | |
| Incomplete outcome date | Statistical analysis was conducted on an intention-to-treat basis. Patients | Low risk |

| | | |
|---|---|---|
| | were treated and assessed after the 4 weeks of intervention (0% loss to post-intervention). All patients from both groups completed the 12 sessions without adverse effects. | |
| **Reporting bias** | | |
| Selective reporting | ClinicalTrials.gov (NCT01804608). | Unclear risk |
| **Other bias** | | |
| Other sources of bias | unknown | Unclear risk |

| **Study: Alireza Motealleh 2019** | | |
|---|---|---|
| **Domain** | **Support for judgement** | **Review authors' judgement** |
| **Selection bias** | | |
| Random sequence generation | "The participants then were assigned randomly to the intervention or the control group with a block randomization method (14 blocks, 2 block sizes), using a random number generator in randomization software." | Low risk |
| Allocation concealment | The results of group allocation were concealed via drawing from a sealed opaque envelope. | Low risk |
| **Performance bias** | | |
| Blinging of participants and personnel | The participants in each group were not aware of the treatment plan of the other group.<br>Insufficient information about personnel. | Low risk<br><br>Unclear risk |
| **Detection bias** | | |
| Blinding of outcome assessment | The physiotherapist who measured the outcome measures was blinded to the patient's allocation group. | Low risk |
| **Attrition bias** | | |

| | | |
|---|---|---|
| Incomplete outcome date | A total of 28 women participated in this study. All patients who received the intervention were analyzed. | Low risk |
| **Reporting bias** | | |
| Selective reporting | This study was registered in the Iranian Registry of Clinical Trials (IRCT2014021315932N2). | Unclear risk |
| **Other bias** | | |
| Other sources of bias | unknown | Unclear risk |

| **Study: Benjamin T Drew 2017** | | |
|---|---|---|
| **Domain** | **Support for judgement** | **Review authors' judgement** |
| **Selection bias** | | |
| Random sequence generation | Randomisation The random allocation sequence was made according to the output from a random number generator. | Low risk |
| Allocation concealment | The random allocation sequence was concealed within pre-sealed, opaque envelopes. | Low risk |
| **Performance bias** | | |
| Blinging of participants and personnel | Insufficient information<br><br>Intervention blinding is not possible for a physiotherapeutic intervention of this nature. | Unclear risk<br><br>High risk |
| **Detection bias** | | |
| Blinding of outcome assessment | "The outcome assessor was unblinded, however, patient reported outcome measures (PROMs) were completed in a separate room with no input from the assessor. The biomechanical outcomes were acquired in accordance to a | Low risk |

| | | |
|---|---|---|
| | strict study protocol to minimise variation and bias. Furthermore, the biomechanical outputs are automated so the lack of blinding is less of an issue.” | |
| **Attrition bias** | | |
| Incomplete outcome date | “At post-treatment follow up, two participants did not complete the study, an attrition rate of 8%. In the MT group, one participant did not attend their second treatment session and was then lost to contact. In the UC group, one participant was unable to complete the post-treatment analysis due to work commitments.”<br>Conversion to consent (100%), missing data (0%), attrition rate (8%) and adherence to both treatment and appointments (>90%) were deemed successful endpoints. | Low risk |
| **Reporting bias** | | |
| Selective reporting | This study was registered retrospectively. ISRCTN74560952. | Unclear risk |
| **Other bias** | | |
| Other sources of bias | unknown | Unclear risk |

| **Study: G Syme 2008** | | |
|---|---|---|
| **Domain** | **Support for judgement** | **Review authors' judgement** |
| **Selection bias** | | |
| Random sequence generation | One independent physiotherapist carried out the individual randomisation procedure out of view of the researcher. | Low risk |
| Allocation concealment | Block randomisation (in blocks of three) was employed through assigning numbers to permutations of the ABC sequence with each letter representing | Low risk |

| | | |
|---|---|---|
| | an arm of the study (Altman and Bland, 1999). These sequence blocks were placed in opaque sealed envelopes and randomly shuffled. | |
| **Performance bias** | | |
| Blinging of participants and personnel | Blinding of participants was not possible owing to the nature of the study.<br><br>Insufficient information | High risk<br><br>Unclear risk |
| **Detection bias** | | |
| Blinding of outcome assessment | The researcher (blind assessor) was not involved in the management of the study participants until their involvement in the study had been completed. | Low risk |
| **Attrition bias** | | |
| Incomplete outcome date | “69 patients were randomised into three groups (Selective n = 23, General n =23 and Control n =23).Two （withdrew owing to work Commitments ） from the Selective group, one （withdrew no further contact with department ） from the General group and three from the Control group failed to complete the study for various reasons（1 withdrew no further contact with department； 1 withdrew owing to pregnancy； 1 withdrawn owing to undisclosed insurance claim）” | Low risk |
| **Reporting bias** | | |
| Selective reporting | unknown | Unclear risk |
| **Other bias** | | |
| Other sources of bias | unknown | Unclear risk |

| **Study: Farzin Halabchi 2015** | | |
|---|---|---|
| **Domain** | **Support for judgement** | **Review authors’ judgement** |

| | | |
|---|---|---|
| **Selection bias** | | |
| Random sequence generation | Patients were allocated to the intervention or control group using the balanced block randomization with random block sizes of 4, 6, and 8. The sequence was generated by a data collector using opaque envelopes and the other researcher, who was not aware of this sequence. | Low risk |
| Allocation concealment | the other researcher, who was not aware of this sequence, enrolled the participants and assigned them to intervention or control groups. | Low risk |
| **Performance bias** | | |
| Blinging of participants and personnel | unknown | Unclear risk |
| **Detection bias** | | |
| Blinding of outcome assessment | unknown | Unclear risk |
| **Attrition bias** | | |
| Incomplete outcome date | "During the 12-week period, 7 participants lost to follow-up (3 in the control and 4 in the intervention groups) and 53 patients completed the trial." | Low risk |
| **Reporting bias** | | |
| Selective reporting | Study protocol was registered in the Iranian Trial Registry (http://www.irct.ir, trial No. 201010305050N1). | Unclear risk |
| **Other bias** | | |
| Other sources of bias | unknown | Unclear risk |

| **Study: R van Linschoten 2009** | | |
|---|---|---|
| **Domain** | **Support for judgement** | **Review authors' judgement** |
| **Selection bias** | | |

| | | |
|---|---|---|
| Random sequence generation | "The randomisation was done by an independent researcher who used a computer-generated list in which patients were stratified by age (14-17 years or 18 years and older) and by recruiting physician (GP or sport physician)" | Low risk |
| Allocation concealment | "…patients were randomly allocated to the intervention (exercise therapy) or the control (usual care). The randomisation was done by an independent researcher who used a computer-generated list…" | Low risk |
| **Performance bias** | | |
| Blinging of participants | Patients could not be blinded. "…patients in the intervention group cannot be blinded for the exercise therapy and, therefore, may be biased for positive outcome (placebo effect)" | High risk |
| Blinging of personnel | This was not described in the study. | Unclear risk |
| **Detection bias** | | |
| Blinding of outcome assessment | "…a blinded external observer could be used to provide objective and observational measures of functional outcomes." | Low risk |
| **Attrition bias** | | |
| Incomplete outcome date | "Outcomes at 3 and 12 months were missing for some patients, but available data from other time points were included in the analyses. This approach meant that the number of patients was not always equal for the different outcome measures." For continuous outcome data, plausible effect size (difference in means or standardized difference in means) among missing outcomes enough to induce clinically relevant bias in observed effect size. | Low risk |
| **Reporting bias** | | |

| | | |
|---|---|---|
| Selective reporting | Trial registration ISRCTN83938749.<br>Secondary outcome measures, as cost-utility (economical evaluation), was not reported.<br>ISRCTN - ISRCTN83938749: Exercise therapy for patello-femoral pain syndrome (PFPS): a randomised clinical trial in general practice and sports medicine | Unclear risk |
| **Other bias** | | |
| Other sources of bias | "…the use of an exercise diary in the intervention group to assess compliance may have caused a bias owing to awareness of being involved in a study (Hawthorne effect)" | Unclear risk |

| **Study: Erik Witvrouw 2004** | | |
|---|---|---|
| **Domain** | **Support for judgement** | **Review authors' judgement** |
| **Selection bias** | | |
| Random sequence generation | "60 PFPS patients…were randomized (by opening a sealed and numbered envelope) into a 5-week conservative rehabilitation protocol." | Low risk |
| Allocation concealment | "…by opening a sealed and numbered envelope" | Low risk |
| **Performance bias** | | |
| Blinging of participants | This was not described in the study. | Unclear risk |
| Blinging of personnel | This was not described in the study. | Unclear risk |
| **Detection bias** | | |
| Blinding of outcome assessment | "In this study, all tests at all evaluation periods were performed by the same examiners, who were blinded to the study and familiar with the different tests." | Low risk |

| Attrition bias | | |
|---|---|---|
| Incomplete outcome date | The number of missing data was similar in all intervention groups, and the causes of missing data were similar in each group. "Fifty-one of these 60 patients (85%) attended the 5-year follow-up evaluation. Two (1 from each treatment group) of these 51 patients underwent surgical intervention (1 patellar release and 1 shaving of the patellar surface). Therefore, the results of these patients were not used in the statistical analysis of this follow-up study. Of the 9 patients who were not evaluated at the 5-year follow-up, 2 were injured (1 had a fracture of the tibia, and 1 had an ear operation) at the time of the evaluation and were unable to attend the evaluation, 1 had died in a car accident, 3 had moved (1 to the United States, 2 to France), and 3 could not be traced. Twenty-five of 49 included patients (16 women and 9 men) who were initially treated by a CKC exercise program, whereas 24 (16 women and 8 men) followed the OKC exercise protocol. The mean age of the patients was 24.8 years (range, 19-36 years)." | Low risk |
| **Reporting bias** | | |
| Selective reporting | This was not described in the study. | Unclear risk |
| **Other bias** | | |
| Other sources of bias | This was not described in the study. | Unclear risk |

| Study: Alexandra Hott 2020 | | |
|---|---|---|
| **Domain** | **Support for judgement** | **Review authors' judgement** |

| | | |
|---|---|---|
| **Selection bias** | | |
| Random sequence generation | "The randomization sequence was computer-generated, stratified by sex and consisted of blocks of a variable size, unknown to any of the research team." | Low risk |
| Allocation concealment | "The sequence was concealed in opaque envelopes and stored by an independent nurse, who delivered them sequentially to the study physiotherapist at randomization." | Low risk |
| **Performance bias** | | |
| Blinging of participants | "It was not possible to blind the patients or physiotherapists who provided the interventions." | High risk |
| Blinging of personnel | "The physiotherapists providing the interventions were blinded to baseline measures…It was not possible to blind the patients or physiotherapists who provided the interventions." | High risk |
| **Detection bias** | | |
| Blinding of outcome assessment | "All outcome measures were collected by blinded observers." | Low risk |
| **Attrition bias** | | |
| Incomplete outcome date | "There were generally few missing data (<5% for all patient-reported outcomes), with the exception of muscle strength testing at 12 months, which 14 patients (Hip, n=6; knee, n=3; control, n=5) were unable to attend and thus only submitted patient-reported outcomes. Missing data analysis found no systematic differences between those attending muscle strength testing compared to those who did not. Thus, no imputation was performed." | Low risk |
| **Reporting bias** | | |
| Selective reporting | "All outcome measures are explained in detail in the published | Unclear risk |

| | | |
|---|---|---|
| | protocol."<br>"…registered with the ClinicalTrials.gov database (reference number: NCT02114294)" | |
| **Other bias** | | |
| Other sources of bias | This was not described in the study. | Unclear risk |

| **Study: Marcelo Camargo Saad 2018** | | |
|---|---|---|
| **Domain** | **Support for judgement** | **Review authors' judgement** |
| **Selection bias** | | |
| Random sequence generation | "The randomization schedule was generated using R 2.7.2 statistical software." | Low risk |
| Allocation concealment | "The allocation was concealed by the use of consecutively numbered, sealed and opaque envelopes." | Low risk |
| **Performance bias** | | |
| Blinging of participants | This was not described in the study. | Unclear risk |
| Blinging of personnel | "Only LVOM, MSBM and RFL (authors) were blinded to the group allocation, because MCS (author) provided all treatments to the groups." | High risk |
| **Detection bias** | | |
| Blinding of outcome assessment | "All kinematic assessments were performed by RFL (author)… LVOM, MSBM and RFL (authors) were blinded to the group allocation… LVOM and MSBM were also responsible for the administration of the questionnaires, intensity of pain assessment and muscle strength assessment tests." | Low risk |
| **Attrition bias** | | |

| | | |
|---|---|---|
| Incomplete outcome date | The generation of missing data is unlikely to be related to the true outcome. "lost at end of intervention, n=1" | Low risk |
| **Reporting bias** | | |
| Selective reporting | This study was approved by Brazilian Clinical Trials Registry registration number: RBR-6tc7mj (http://www.ensaiosclinicos.gov.br/rg/RBR-6tc7mj/). | Low risk |
| **Other bias** | | |
| Other sources of bias | "…participants' baseline characteristics seemed to be different, especially for pain, probably as a consequence of low number of participants." | High risk |

| **Study: Alexandra Hott 2019** | | |
|---|---|---|
| **Domain** | **Support for judgement** | **Review authors' judgement** |
| **Selection bias** | | |
| Random sequence generation | "The randomization sequence was computer generated with blocks of a variable size, stratified by sex, and unknown to anyone in the research team. " | Low risk |
| Allocation concealment | "The sequence was concealed in opaque envelopes, stored by a nurse not otherwise involved in the study, and delivered sequentially to the study physiotherapist at randomization." | Low risk |
| **Performance bias** | | |
| Blinging of participants | "Although blinding of the participants to group allocation was not possible, expectations of the effects of the different interventions were investigated at inclusion and were not significantly different among groups." | Low risk |

| | | |
|---|---|---|
| Blinging of personnel | “Physiotherapists providing the interventions were blinded to baseline measures.” “It was not possible to blind the participants or the physiotherapists who provided the interventions.” | High risk |
| **Detection bias** | | |
| Blinding of outcome assessment | “Members of the research team who handled outcome measures were blinded to treatment allocation.” | Low risk |
| **Attrition bias** | | |
| Incomplete outcome date | “Missing values in the AKPS, Tampa Scale for Kinesiophobia, Knee Self-efficacy Scale, and Hopkins Symptom Checklist were treated as follows: If $< 25\%$ of items were missing, the values were substituted with the arithmetic mean of values from the available items. If $\geq 25\%$ of items were missing,<br>the outcome was regarded as missing for the patient.”<br>“The trial was adequately powered, with good compliance and a low rate of dropout (7%) and missing data.” | Low risk |
| **Reporting bias** | | |
| Selective reporting | “NCT02114294 (ClinicalTrials.gov identifier).”<br>Secondary outcome measures are incomplete. | Unclear risk |
| **Other bias** | | |
| Other sources of bias | “The standardization of written and oral communication with all patients regardless of group allocation, including presentation of the control<br>group as an active treatment group, also likely reduced bias.”<br>“Data analysis and the first draft of the manuscript were performed blinded to group allocation to avoid bias at this stage of the study.” | Low risk |

| Study: Reed Ferber 2014 | | |
|---|---|---|
| **Domain** | **Support for judgement** | **Review authors' judgement** |
| **Selection bias** | | |
| Random sequence generation | "The randomization sequence was developed and kept at the University of Calgary (Alberta, Canada) by the research coordinator, using a random number generator, and the same sequence of randomization was used at each site." | Low risk |
| Allocation concealment | Insufficient information to permit judgement of 'Low risk' or 'High risk'. "Next, the patients with PFP were randomly assigned to receive 1 of 2 treatment protocols (HIP or KNEE)." | Unclear risk |
| **Performance bias** | | |
| Blinging of participants | This was not described in the study. | Unclear risk |
| Blinging of personnel | This was not described in the study. | Unclear risk |
| **Detection bias** | | |
| Blinding of outcome assessment | "Only the athletic trainer (AT) at each site communicated with the research coordinator to ensure that the investigators, who were responsible for outcomes measurement and data analysis, remained blinded to group allocation." | Low risk |
| **Attrition bias** | | |
| Incomplete outcome date | The rate of lost follow-up is high. "At the post rehabilitation follow-up, we collected outcomes and clinical data from 146 of the 199 patients (73.4%) with PFP." | High risk |
| **Reporting bias** | | |

| | | |
|---|---|---|
| Selective reporting | Insufficient information to permit judgement of 'Low risk' or 'High risk'. | Unclear risk |
| **Other bias** | | |
| Other sources of bias | Insufficient rationale or evidence that an identified problem will introduce bias. | Unclear risk |

| **Study: Kimberly L Dolak 2011** | | |
|---|---|---|
| **Domain** | **Support for judgement** | **Review authors' judgement** |
| **Selection bias** | | |
| Random sequence generation | "…group allocation for each participant was made with a random-number generator in Microsoft Excel (Microsoft Corporation, Redwood, WA)." | Low risk |
| Allocation concealment | Insufficient information to permit judgement of 'Low risk' or 'High risk'.<br>"Prior to initiation of the study, group allocation for each participant was made with a random-number generator in Microsoft Excel (Microsoft Corporation, Redwood, WA)...participants were randomly assigned to a hip strengthening program (hip group) or a quadriceps-strengthening program (quad group) for 4 weeks." | Low risk |
| **Performance bias** | | |
| Blinging of participants | This was not described in the study. | Unclear risk |
| Blinging of personnel | "The researcher responsible for setup and testing was blinded to participants' group assignment during the initial testing session."<br>"Testers were not blinded to participants' group assignment after | High risk |

| | | |
|---|---|---|
| | baseline testing, mostly due to the large number of patient exercise sessions supervised.” | |
| **Detection bias** | | |
| Blinding of outcome assessment | “The researcher responsible for setup and testing was blinded to participants’ group assignment during the initial testing session.”<br>“Blinding of the investigators after initial testing was a further limitation of the study.” | High risk |
| **Attrition bias** | | |
| Incomplete outcome date | No missing data processing instructions.<br>“Three women did not complete this phase. Two removed themselves from the study due to time constraints and 1 withdrew because of injuries sustained during an unrelated motor vehicle accident.”<br>“Three women did not complete this phase. Two removed themselves for unknown reasons and 1 was withdrawn by investigators for increased pain.”<br>“One woman (hip group) did not complete this phase due to time constraints.” | Unclear risk |
| **Reporting bias** | | |
| Selective reporting | Trial registration: ClinicalTrials.gov NCT00445224. | Unclear risk |
| **Other bias** | | |
| Other sources of bias | Insufficient rationale or evidence that an identified problem will introduce bias. | Unclear risk |

| **Study: Mehtap Şahin 2016** |
|---|

| Domain | Support for judgement | Review authors' judgement |
|---|---|---|
| **Selection bias** | | |
| Random sequence generation | "At the beginning of the program, a nurse prepared 56 small pieces of opaque paper numbered as 1 or 2, which were folded with the treatment on the inside, and patients picked these small papers from the box." | Low risk |
| Allocation concealment | "Group allocation was hidden from observers using empty patient files prepared by a nurse. The success of masking was checked by asking verbal questions about group allocation." | Low risk |
| **Performance bias** | | |
| Blinging of participants | "Patients were not blind to the treatment group because of the nature of the exercise programs." | High risk |
| Blinging of personnel | This was not described in the study. | Unclear risk |
| **Detection bias** | | |
| Blinding of outcome assessment | This was not described in the study. | Unclear risk |
| **Attrition bias** | | |
| Incomplete outcome date | No missing data processing instructions.<br>"Losses (N3): 2, 1: transport problems, 1: surgery; Losses (N3) : 3, 1: transport problems, 2: trauma" | Unclear risk |
| **Reporting bias** | | |
| Selective reporting | This was not described in the study. | Unclear risk |
| **Other bias** | | |
| Other sources of bias | Insufficient rationale or evidence that an identified problem will introduce bias. | Unclear risk |

| Study: Thiago Yukio Fukuda 2012 | | |
|---|---|---|
| **Domain** | **Support for judgement** | **Review authors' judgement** |
| **Selection bias** | | |
| Random sequence generation | "The assignment of subjects to the 2 groups was performed randomly using opaque, sealed envelopes, each containing the name of one of the groups (KE or KHE)." | Low risk |
| Allocation concealment | "The assignment of subjects to the 2 groups was performed randomly using opaque, sealed envelopes, each containing the name of one of the groups (KE or KHE). The envelopes were picked by an individual not involved in the study." | Low risk |
| **Performance bias** | | |
| Blinging of participants | Insufficient information to permit judgement of 'Low risk' or 'High risk'. | Unclear risk |
| Blinging of personnel | Insufficient information to permit judgement of 'Low risk' or 'High risk'. | Unclear risk |
| **Detection bias** | | |
| Blinding of outcome assessment | "The examiner was blind to the group assignment of the patients and did not participate in the intervention." | Low risk |
| **Attrition bias** | | |
| Incomplete outcome date | "An intention-to-treat analysis was performed using the last value-carried-forward method to impute values for all missing data." | Low risk |
| **Reporting bias** | | |
| Selective reporting | Insufficient information to permit judgement of 'Low risk' or | Unclear risk |

| | | |
|---|---|---|
| | 'High risk'. | |
| **Other bias** | | |
| Other sources of bias | Insufficient information to assess whether an important risk of bias exists. | Unclear risk |

| **Study: Thiago Yukio Fukuda 2011** | | |
|---|---|---|
| **Domain** | **Support for judgement** | **Review authors' judgement** |
| **Selection bias** | | |
| Random sequence generation | "The assignment of patients in the 3 groups was performed randomly using<br>opaque and sealed envelopes containing the names of the groups: CO, KE, and KHE." | Low risk |
| Allocation concealment | "The assignment of patients in the 3 groups was performed randomly using<br>opaque and sealed envelopes containing the names of the groups: CO, KE, and KHE. The envelopes were picked by an individual not involved in this study." | Low risk |
| **Performance bias** | | |
| Blinging of participants | Insufficient information to permit judgement of 'Low risk' or 'High risk'. | Unclear risk |
| Blinging of personnel | Insufficient information to permit judgement of 'Low risk' or 'High risk'. | Unclear risk |
| **Detection bias** | | |
| Blinding of outcome assessment | "This examiner was blind to the group assignment of the patients and did not participate in the intervention." | Low risk |

| | | |
|---|---|---|
| **Attrition bias** | | |
| Incomplete outcome date | "We performed an intention-to-treat analysis, 24 and the results were consistent with the per-protocol analysis as presented above. This parallel method was based on the imputation of the group mean to each missing value for each of the 3 groups (CO, KE, and KHE)." | Low risk |
| **Reporting bias** | | |
| Selective reporting | Insufficient information to permit judgement of 'Low risk' or 'High risk'. | Unclear risk |
| **Other bias** | | |
| Other sources of bias | Insufficient information to assess whether an important risk of bias exists. | Unclear risk |

| **Study: Theresa Helissa Nakagawa 2008** | | |
|---|---|---|
| **Domain** | **Support for judgement** | **Review authors' judgement** |
| **Selection bias** | | |
| Random sequence generation | "…14 preprinted cards in opaque sealed envelopes from a box (seven labelled 'intervention group' and seven labelled 'control group') ..." | Low risk |
| Allocation concealment | "After baseline assessment, participants blindly drew one of 14 preprinted cards in opaque sealed envelopes from a box (seven labelled 'intervention group' and seven labelled 'control group') and were placed in the intervention or control group in accordance with the card drawn." | Low risk |

| | | |
|---|---|---|
| **Performance bias** | | |
| Blinging of participants | "Participants were blind to treatment allocation; both groups received therapeutic exercises as intervention in individual sessions." | Low risk |
| Blinging of personnel | "The principal investigator remained blind to treatment allocation until all baseline assessment had been completed. After this point, blinding of the principal investigator was not feasible." | High risk |
| **Detection bias** | | |
| Blinding of outcome assessment | "…two blind assessors conducted the assessments to minimize any communication between the participant and the researcher that might interfere with the results." | Low risk |
| **Attrition bias** | | |
| Incomplete outcome date | "All the participants completed the rehabilitation protocol and assessment sessions." | Low risk |
| **Reporting bias** | | |
| Selective reporting | Insufficient information to permit judgement of 'Low risk' or 'High risk'. | Unclear risk |
| **Other bias** | | |
| Other sources of bias | Insufficient information to assess whether an important risk of bias exists. "It cannot be ignored that a larger sample size could have altered some of the results of the study; and therefore further research is required before definite conclusions can be drawn." | Unclear risk |

| Study: Khayambashi, Khalil 2012 | | |
|---|---|---|
| **Domain** | **Support for judgement** | **Review authors' judgement** |
| **Selection bias** | | |
| Random sequence generation | 28 participants were sequentially assigned in an alternating fashion to the exercise or control group | High risk |
| Allocation concealment | unknown | Unclear risk |
| **Performance bias** | | |
| Blinging of participants and personnel | Participants were aware of an alternative treatment group in the study but had no knowledge of intervention details.<br>Insufficient information to judge. | Unclear risk<br><br>Unclear risk |
| **Detection bias** | | |
| Blinding of outcome assessment | the investigator who was responsible for obtaining the functional outcome and strength measures was not blinded to the participants group assignment. While this lack of blinding would have had no influence on the self-reported outcomes, potential bias must be acknowledged regarding the strength testing。 | Low risk |
| **Attrition bias** | | |
| Incomplete outcome date | No patients dropped out of the study。 | Low risk |
| **Reporting bias** | | |
| Selective reporting | unknown | Unclear risk |
| **Other bias** | | |
| Other sources of bias | unknown | Unclear risk |

| Study: Song, Chen-Yi 2009 |
|---|

| Domain | Support for judgement | Review authors' judgement |
|---|---|---|
| **Selection bias** | | |
| Random sequence generation | A single physical therapist, unaware of the purpose of the study, was responsible for randomization and interventions. Stratified allocation was carried out with regard to the number of affected sides (unilateral or bilateral) and symptom severity (Lysholm scale scores ≥65 or <65). | Low risk |
| Allocation concealment | chosen through numbered opaque envelopes | Low risk |
| **Performance bias** | | |
| Blinging of participants and personnel | unknown | Unclear risk |
| **Detection bias** | | |
| Blinding of outcome assessment | Two assessment sessions were performed by another physical therapist (blinded to each patient s grouping) before and after the 8-week intervention. | Low risk |
| **Attrition bias** | | |
| Incomplete outcome date | Ten participants later dropped out of the study due to personal factors (not knee pain) or work. LPHA Group: dropped out 2; LP Group: dropped out 3; Control Group: dropped out 5. Data were subjected to an intention-to-treat analysis and included all dropouts.<br>Unclear is whether the missing data were estimated using appropriate methods. | Unclear risk |
| **Reporting bias** | | |
| Selective reporting | unknown | Unclear risk |
| **Other bias** | | |
| Other sources of bias | unknown | Unclear risk |

| **Study:** Emamvirdi, Mahsa 2019 | | |
|---|---|---|
| **Domain** | **Support for judgement** | **Review authors' judgement** |
| **Selection bias** | | |
| Random sequence generation | a computer-generated table of random numbers | Low risk |
| Allocation concealment | Randomization was performed in blocks of 4. Consecutively numbered, opaque envelopes were randomly assigned by a computer-generated table of random numbers. | Low risk |
| **Performance bias** | | |
| Blinging of participants and personnel | An individual blinded to patient data performed the randomization and provided the group assignment to a physical therapist. To ensure that participants were unaware of the exercises performed by the other group, the groups were given program instructions separately. | Low risk |
| **Detection bias** | | |
| Blinding of outcome assessment | No | High risk |
| **Attrition bias** | | |
| Incomplete outcome date | The VCI and control groups had a participation rate of 100% during the study period. | Low risk |
| **Reporting bias** | | |
| Selective reporting | unknown | Unclear risk |
| **Other bias** | | |
| Other sources of bias | unknown | Unclear risk |

| **Study: Bolgla, Lori A** 2016 | | |
|---|---|---|
| **Domain** | **Support for judgement** | **Review authors' judgement** |
| **Selection bias** | | |
| Random sequence generation | A random number generator was used to assign subjects to either the hip/core or knee program | Low risk |
| Allocation concealment | unknown | Unclear risk |
| **Performance bias** | | |
| Blinging of participants and personnel | unknown | Unclear risk |
| **Detection bias** | | |
| Blinding of outcome assessment | Subjects were randomly assigned to exercise group and examiners were blinded to subject group assignment. | Low risk |
| **Attrition bias** | | |
| Incomplete outcome date | An intention-to-treat analysis, using a conservative method where missing data were replaced with the last score carried forward, was used. | Low risk |
| **Reporting bias** | | |
| Selective reporting | unknown | Unclear risk |
| **Other bias** | | |
| Other sources of bias | unknown | Unclear risk |

#### Web Appendix7. GRADE assessment for primary outcomes

The five GRADE criteria below that are deem most significant for down rating the quality in NMAs by rating -1 (serious concern) or -2 (very serious concern) for the following reasons:

1. Risk of Bias criteria: Downgraded when comparisons failed to conceal ‘random allocation’ or ‘blinding of outcome assessors’ within RoB assessment for the eligible trials.

2. Inconsistency (or heterogeneity): For heterogeneity, we looked at the common tau and compared to the expected value as reported in the literature (Turner RM et al (2012) Int J Epidemiol, 41, 818-827: https://www.ncbi.nlm.nih.gov/pmc/articles/PMC3396310/) which was reported as 75% (95% CI: 58-95%).

For inconsistency, we looked at the results of the node-splitting analysis and we downgraded the comparisons with important inconsistency ($p<0.10$).

3. Indirectness (PICO and applicability): we downgraded single connected nodes for indirectness because evaluation of transitivity for such nodes is unclear.

4. Imprecision: Using Cohen’s probably of benefit we chose the comparison that reported a wide 95% confidence interval and include or are close to null effect, otherwise they were not necessary for downgrading.

5. Publication bias: The comparison-adjusted funnel plots did not suggest presence of overall publication bias. We are confident that we have all available information that is possible to capture from the bibliographic databases. However, we cannot completely rule out the possibility that some studies are still missing. Due to this we decide to downgrade all studies for potential publication bias by one level.

| Comparison | Risk of bias | Inconsistency | Indirectness | Imprecision | Publication bias | Quality of evidence |
|---|---|---|---|---|---|---|
| Pain intensity measured with VAS | | | | | | |
| Nontreatment VS Proprioceptive Neuromuscular Facilitation + Exercise | Serious | No | No serious indirectness | Yes | ? | Low |
| Nontreatment VS Whole Body Vibration Exercise | Serious | No | No serious indirectness | Yes | ? | Low |
| Nontreatment VS Knee Exercise + Hip Exercise | Very serious | No | No | Yes | ? | Modorate |
| Nontreatment VS Foot Orthoses + Exercise | Serious | No | No serious indirectness | Yes | ? | Low |
| Nontreatment VS Hip Exercise | Very serious | No | No | Yes | ? | Modorate |
| Nontreatment VS Knee Brace + Exercise | Serious | No | No serious indirectness | Yes | ? | Low |
| Nontreatment VS Supervised Exercise | Serious | No | No serious indirectness | Yes | ? | Low |
| Nontreatment VS Gait Retraining Exercise | Serious | No | No serious indirectness | Yes | ? | Low |
| Nontreatment VS Keen Exercise | Serious | No | No serious indirectness | Yes | ? | Low |
| Nontreatment VS Blood Flow Restriction Exercise | Serious | No | No serious indirectness | Yes | ? | Low |

| | | | | | | |
|---|---|---|---|---|---|---|
| Nontreatment VS Knee Arthroscopy + Exercise | Serious | No | No serious indirectness | Yes | ? | Low |
| Nontreatment VS Target Exercise | Very serious | No | No | Yes | ? | Modorate |
| Nontreatment VS KinesioTaping + Exercise | Very serious | No | No | Yes | ? | Modorate |
| Nontreatment VS Manipulation Treatment +Exercise | Serious | No | No serious indirectness | Yes | ? | Low |
| Nontreatment VS Education + Exercise | Serious | No | No serious indirectness | Yes | ? | Low |
| Nontreatment VS Motor Control Training | Serious | No | No serious indirectness | Yes | ? | Low |
| Nontreatment VS General Exercise | Very serious | No | No | Yes | ? | Modorate |
| Nontreatment VS Kinetic Chain Exercise | Serious | No | No serious indirectness | Yes | ? | Low |
| Nontreatment VS Feedback Exercise | Serious | No | No serious indirectness | Yes | ? | Low |
| Feedback Exercise VS Proprioceptive Neuromuscular Facilitation + Exercise | Serious | No | No serious indirectness | Yes | ? | Low |
| Feedback Exercise VS Whole Body Vibration Exercise | Serious | No | No serious indirectness | Yes | ? | Low |
| Feedback Exercise VS Knee Exercise + Hip Exercise | Serious | No | No serious indirectness | Yes | ? | Low |

| | | | | | | |
|---|---|---|---|---|---|---|
| Feedback Exercise VS Foot Orthoses + Exercise | Serious | No | No serious indirectness | Yes | ? | Low |
| Feedback Exercise VS Hip Exercise | Serious | No | No serious indirectness | Yes | ? | Low |
| Feedback Exercise VS Knee Brace + Exercise | Serious | No | No serious indirectness | Yes | ? | Low |
| Feedback Exercise VS Supervised Exercise | Serious | No | No serious indirectness | Yes | ? | Low |
| Feedback Exercise VS Gait Retraining Exercise | Serious | No | No serious indirectness | Yes | ? | Low |
| Feedback Exercise VS Keen Exercise | Serious | No | No serious indirectness | Yes | ? | Low |
| Feedback Exercise VS Blood Flow Restriction Exercise | Serious | No | No serious indirectness | Yes | ? | Low |
| Feedback Exercise VS Knee Arthroscopy + Exercise | Serious | No | No serious indirectness | Yes | ? | Low |
| Feedback Exercise VS Target Exercise | Serious | No | No serious indirectness | Yes | ? | Low |
| Feedback Exercise VS KinesioTaping + Exercise | Serious | No | No serious indirectness | Yes | ? | Low |
| Feedback Exercise VS Manipulation Treatment +Exercise | Serious | No | No serious indirectness | Yes | ? | Low |
| Feedback Exercise VS Education + Exercise | Serious | No | No serious indirectness | Yes | ? | Low |

| | | | | | | |
|---|---|---|---|---|---|---|
| Feedback Exercise VS Motor Control Training | Serious | No | No serious indirectness | Yes | ? | Low |
| Feedback Exercise VS General Exercise | Very serious | No | No | Yes | ? | Modorate |
| Feedback Exercise VS Kinetic Chain Exercise | Serious | No | No serious indirectness | Yes | ? | Low |
| Kinetic Chain Exercise VS Proprioceptive Neuromuscular Facilitation + Exercise | Serious | No | No serious indirectness | Yes | ? | Low |
| Kinetic Chain Exercise VS Whole Body Vibration Exercise | Serious | No | No serious indirectness | Yes | ? | Low |
| Kinetic Chain Exercise VS Knee Exercise + Hip Exercise | Serious | No | No serious indirectness | Yes | ? | Low |
| Kinetic Chain Exercise VS Foot Orthoses + Exercise | Serious | No | No serious indirectness | Yes | ? | Low |
| Kinetic Chain Exercise VS Hip Exercise | Serious | No | No serious indirectness | Yes | ? | Low |
| Kinetic Chain Exercise VS Knee Brace + Exercise | Serious | No | No serious indirectness | Yes | ? | Low |
| Kinetic Chain Exercise VS Supervised Exercise | Serious | No | No serious indirectness | Yes | ? | Low |
| Kinetic Chain Exercise VS Gait Retraining Exercise | Serious | No | No serious indirectness | Yes | ? | Low |
| Kinetic Chain Exercise VS Keen Exercise | Serious | No | No serious indirectness | Yes | ? | Low |

| | | | | | | |
|---|---|---|---|---|---|---|
| Kinetic Chain Exercise VS Blood Flow Restriction Exercise | Serious | No | No serious indirectness | Yes | ? | Low |
| Kinetic Chain Exercise VS Knee Arthroscopy + Exercise | Serious | No | No serious indirectness | Yes | ? | Low |
| Kinetic Chain Exercise VS Target Exercise | Serious | No | No serious indirectness | Yes | ? | Low |
| Kinetic Chain Exercise VS KinesioTaping + Exercise | Serious | No | No serious indirectness | Yes | ? | Low |
| Kinetic Chain Exercise VS Manipulation Treatment +Exercise | Serious | No | No serious indirectness | Yes | ? | Low |
| Kinetic Chain Exercise VS Education + Exercise | Serious | No | No serious indirectness | Yes | ? | Low |
| Kinetic Chain Exercise VS Motor Control Training | Serious | No | No serious indirectness | Yes | ? | Low |
| Kinetic Chain Exercise VS General Exercise | Very serious | No | No | Yes | ? | Modorate |
| General Exercise VS Proprioceptive Neuromuscular Facilitation + Exercise | Very serious | No | No | Yes | ? | Modorate |
| General Exercise VS Whole Body Vibration Exercise | Very serious | No | No | Yes | ? | Modorate |
| General Exercise VS Knee Exercise + Hip Exercise | Serious | No | No serious indirectness | Yes | ? | Low |
| General Exercise VS Foot Orthoses + Exercise | Very serious | No | No | Yes | ? | Modorate |

| | | | | | | |
|---|---|---|---|---|---|---|
| General Exercise VS Hip Exercise | Very serious | No | No | Yes | ? | Modorate |
| General Exercise VS Knee Brace + Exercise | Very serious | No | No | Yes | ? | Modorate |
| General Exercise VS Supervised Exercise | Very serious | No | No | Yes | ? | Modorate |
| General Exercise VS Gait Retraining Exercise | Very serious | No | No | Yes | ? | Modorate |
| General Exercise VS Keen Exercise | Very serious | No | No | Yes | ? | Modorate |
| General Exercise VS Blood Flow Restriction Exercise | Very serious | No | No | Yes | ? | Modorate |
| General Exercise VS Knee Arthroscopy + Exercise | Very serious | No | No | Yes | ? | Modorate |
| General Exercise VS Target Exercise | Very serious | No | No | Yes | ? | Modorate |
| General Exercise VS KinesioTaping + Exercise | Very serious | No | No | Yes | ? | Modorate |
| General Exercise VS Manipulation Treatment +Exercise | Very serious | No | No | Yes | ? | Modorate |
| General Exercise VS Education + Exercise | Very serious | No | No | Yes | ? | Modorate |
| General Exercise VS Motor Control Training | Very serious | No | No | Yes | ? | Modorate |

| | | | | | | |
|---|---|---|---|---|---|---|
| Motor Control Training VS Proprioceptive Neuromuscular Facilitation + Exercise | Serious | No | No serious indirectness | Yes | ? | Low |
| Motor Control Training VS Whole Body Vibration Exercise | Serious | No | No serious indirectness | Yes | ? | Low |
| Motor Control Training VS Knee Exercise + Hip Exercise | Serious | No | No serious indirectness | Yes | ? | Low |
| Motor Control Training VS Foot Orthoses + Exercise | Serious | No | No serious indirectness | Yes | ? | Low |
| Motor Control Training VS Hip Exercise | Serious | No | No serious indirectness | Yes | ? | Low |
| Motor Control Training VS Knee Brace + Exercise | Serious | No | No serious indirectness | Yes | ? | Low |
| Motor Control Training VS Supervised Exercise | Serious | No | No serious indirectness | Yes | ? | Low |
| Motor Control Training VS Gait Retraining Exercise | Serious | No | No serious indirectness | Yes | ? | Low |
| Motor Control Training VS Keen Exercise | Serious | No | No serious indirectness | Yes | ? | Low |
| Motor Control Training VS Blood Flow Restriction Exercise | Serious | No | No serious indirectness | Yes | ? | Low |
| Motor Control Training VS Knee Arthroscopy + Exercise | Serious | No | No serious indirectness | Yes | ? | Low |
| Motor Control Training VS Target Exercise | Serious | No | No serious indirectness | Yes | ? | Low |

| | | | | | | |
|---|---|---|---|---|---|---|
| Motor Control Training VS KinesioTaping + Exercise | Serious | No | No serious indirectness | Yes | ? | Low |
| Motor Control Training VS Manipulation Treatment +Exercise | Serious | No | No serious indirectness | Yes | ? | Low |
| Motor Control Training VS Education + Exercise | Serious | No | No serious indirectness | Yes | ? | Low |
| Education + Exercise VS Proprioceptive Neuromuscular Facilitation + Exercise | Serious | No | No serious indirectness | Yes | ? | Low |
| Education + Exercise VS Whole Body Vibration Exercise | Serious | No | No serious indirectness | Yes | ? | Low |
| Education + Exercise VS Knee Exercise + Hip Exercise | Serious | No | No serious indirectness | Yes | ? | Low |
| Education + Exercise VS Foot Orthoses + Exercise | Serious | No | No serious indirectness | Yes | ? | Low |
| Education + Exercise VS Hip Exercise | Serious | No | No serious indirectness | Yes | ? | Low |
| Education + Exercise VS Knee Brace + Exercise | Serious | No | No serious indirectness | Yes | ? | Low |
| Education + Exercise VS Supervised Exercise | Serious | No | No serious indirectness | Yes | ? | Low |
| Education + Exercise VS Gait Retraining Exercise | Very serious | No | No | Yes | ? | Modorate |
| Education + Exercise VS Keen Exercise | Serious | No | No serious indirectness | Yes | ? | Low |

| | | | | | | |
|---|---|---|---|---|---|---|
| Education + Exercise VS Blood Flow Restriction Exercise | Serious | No | No serious indirectness | Yes | ? | Low |
| Education + Exercise VS Knee Arthroscopy + Exercise | Serious | No | No serious indirectness | Yes | ? | Low |
| Education + Exercise VS Target Exercise | Serious | No | No serious indirectness | Yes | ? | Low |
| Education + Exercise VS KinesioTaping + Exercise | Serious | No | No serious indirectness | Yes | ? | Low |
| Education + Exercise VS Manipulation Treatment +Exercise | Serious | No | No serious indirectness | Yes | ? | Low |
| Manipulation Treatment +Exercise VS Proprioceptive Neuromuscular Facilitation | Serious | No | No serious indirectness | Yes | ? | Low |
| Manipulation Treatment +Exercise VS Whole Body Vibration Exercise | Serious | No | No serious indirectness | Yes | ? | Low |
| Manipulation Treatment +Exercise VS Knee Exercise + Hip Exercise | Serious | No | No serious indirectness | Yes | ? | Low |
| Manipulation Treatment +Exercise VS Foot Orthoses + Exercise | Serious | No | No serious indirectness | Yes | ? | Low |
| Manipulation Treatment +Exercise VS Hip Exercise | Serious | No | No serious indirectness | Yes | ? | Low |
| Manipulation Treatment +Exercise VS Knee Brace + Exercise | Serious | No | No serious indirectness | Yes | ? | Low |
| Manipulation Treatment VS Supervised Exercise | Serious | No | No serious indirectness | Yes | ? | Low |

| | | | | | | |
|---|---|---|---|---|---|---|
| Manipulation Treatment VS Gait Retraining Exercise | Serious | No | No serious indirectness | Yes | ? | Low |
| Manipulation Treatment VS Keen Exercise | Serious | No | No serious indirectness | Yes | ? | Low |
| Manipulation Treatment VS Blood Flow Restriction Exercise | Serious | No | No serious indirectness | Yes | ? | Low |
| Manipulation Treatment VS Knee Arthroscopy + Exercise | Serious | No | No serious indirectness | Yes | ? | Low |
| Manipulation Treatment VS Target Exercise | Serious | No | No serious indirectness | Yes | ? | Low |
| Manipulation Treatment VS KinesioTaping + Exercise | Serious | No | No serious indirectness | Yes | ? | Low |
| KinesioTaping + Exercise VS Proprioceptive Neuromuscular Facilitation | Serious | No | No serious indirectness | Yes | ? | Low |
| KinesioTaping + Exercise VS Whole Body Vibration Exercise | Serious | No | No serious indirectness | Yes | ? | Low |
| KinesioTaping + Exercise VS Knee Exercise + Hip Exercise | Serious | No | No serious indirectness | Yes | ? | Low |
| KinesioTaping + Exercise VS Foot Orthoses + Exercise | Serious | No | No serious indirectness | Yes | ? | Low |
| KinesioTaping + Exercise VS Hip Exercise | Serious | No | No serious indirectness | Yes | ? | Low |
| KinesioTaping + Exercise VS Knee Brace + Exercise | Serious | No | No serious indirectness | Yes | ? | Low |

| | | | | | | |
|---|---|---|---|---|---|---|
| KinesioTaping + Exercise VS Supervised Exercise | Serious | No | No serious indirectness | Yes | ? | Low |
| KinesioTaping + Exercise VS Gait Retraining Exercise | Serious | No | No serious indirectness | Yes | ? | Low |
| KinesioTaping + Exercise VS Keen Exercise | Serious | No | No serious indirectness | Yes | ? | Low |
| KinesioTaping + Exercise VS Blood Flow Restriction Exercise | Serious | No | No serious indirectness | Yes | ? | Low |
| KinesioTaping + Exercise VS Knee Arthroscopy + Exercise | Serious | No | No serious indirectness | Yes | ? | Low |
| KinesioTaping + Exercise VS Target Exercise | Serious | No | No serious indirectness | Yes | ? | Low |
| Target Exercise VS Proprioceptive Neuromuscular Facilitation + Exercise | Serious | No | No serious indirectness | Yes | ? | Low |
| Target Exercise VS Whole Body Vibration Exercise | Serious | No | No serious indirectness | Yes | ? | Low |
| Target Exercise VS Knee Exercise + Hip Exercise | Serious | No | No serious indirectness | Yes | ? | Low |
| Target Exercise VS Foot Orthoses + Exercise | Serious | No | No serious indirectness | Yes | ? | Low |
| Target Exercise VS Hip Exercise | Serious | No | No serious indirectness | Yes | ? | Low |
| Target Exercise VS Knee Brace + Exercise | Serious | No | No serious indirectness | Yes | ? | Low |

| | | | | | | |
|---|---|---|---|---|---|---|
| Target Exercise VS Supervised Exercise | Serious | No | No serious indirectness | Yes | ? | Low |
| Target Exercise VS Gait Retraining Exercise | Serious | No | No serious indirectness | Yes | ? | Low |
| Target Exercise VS Keen Exercise | Serious | No | No serious indirectness | Yes | ? | Low |
| Target Exercise VS Blood Flow Restriction Exercise | Serious | No | No serious indirectness | Yes | ? | Low |
| Target Exercise VS Knee Arthroscopy + Exercise | Serious | No | No serious indirectness | Yes | ? | Low |
| Knee Arthroscopy + Exercise VS Proprioceptive Neuromuscular Facilitation | Serious | No | No serious indirectness | Yes | ? | Low |
| Knee Arthroscopy + Exercise VS Whole Body Vibration Exercise | Serious | No | No serious indirectness | Yes | ? | Low |
| Knee Arthroscopy + Exercise VS Knee Exercise + Hip Exercise | Serious | No | No serious indirectness | Yes | ? | Low |
| Knee Arthroscopy + Exercise VS Foot Orthoses + Exercise | Serious | No | No serious indirectness | Yes | ? | Low |
| Knee Arthroscopy + Exercise VS Hip Exercise | Serious | No | No serious indirectness | Yes | ? | Low |
| Knee Arthroscopy + Exercise VS Knee Brace + Exercise | Serious | No | No serious indirectness | Yes | ? | Low |
| Knee Arthroscopy + Exercise VS Supervised Exercise | Serious | No | No serious indirectness | Yes | ? | Low |

| | | | | | | |
|---|---|---|---|---|---|---|
| Knee Arthroscopy + Exercise VS Gait Retraining Exercise | Serious | No | No serious indirectness | Yes | ? | Low |
| Knee Arthroscopy + Exercise VS Keen Exercise | Serious | No | No serious indirectness | Yes | ? | Low |
| Knee Arthroscopy + Exercise VS Blood Flow Restriction Exercise | Serious | No | No serious indirectness | Yes | ? | Low |
| Blood Flow Restriction Exercise VS Proprioceptive Neuromuscular Facilitation | Serious | No | No serious indirectness | Yes | ? | Low |
| Blood Flow Restriction Exercise VS Whole Body Vibration Exercise | Serious | No | No serious indirectness | Yes | ? | Low |
| Blood Flow Restriction Exercise VS Knee Exercise + Hip Exercise | Serious | No | No serious indirectness | Yes | ? | Low |
| Blood Flow Restriction Exercise VS Foot Orthoses + Exercise | Serious | No | No serious indirectness | Yes | ? | Low |
| Blood Flow Restriction Exercise VS Hip Exercise | Serious | No | No serious indirectness | Yes | ? | Low |
| Blood Flow Restriction Exercise VS Knee Brace + Exercise | Serious | No | No serious indirectness | Yes | ? | Low |
| Blood Flow Restriction Exercise VS Supervised Exercise | Serious | No | No serious indirectness | Yes | ? | Low |
| Blood Flow Restriction Exercise VS Gait Retraining Exercise | Serious | No | No serious indirectness | Yes | ? | Low |
| Blood Flow Restriction Exercise VS Keen Exercise | Serious | No | No serious indirectness | Yes | ? | Low |

| | | | | | | |
|---|---|---|---|---|---|---|
| Keen Exercise VS Proprioceptive Neuromuscular Facilitation + Exercise | Serious | No | No serious indirectness | Yes | ? | Low |
| Keen Exercise VS Whole Body Vibration Exercise | Serious | No | No serious indirectness | Yes | ? | Low |
| Keen Exercise VS Knee Exercise + Hip Exercise | Very serious | No | No | Yes | ? | Modorate |
| Keen Exercise VS Foot Orthoses + Exercise | Serious | No | No serious indirectness | Yes | ? | Low |
| Keen Exercise VS Hip Exercise | Very serious | No | No | Yes | ? | Modorate |
| Keen Exercise VS Knee Brace + Exercise | Serious | No | No serious indirectness | Yes | ? | Low |
| Keen Exercise VS Supervised Exercise | Serious | No | No serious indirectness | Yes | ? | Low |
| Keen Exercise VS Gait Retraining Exercise | Serious | No | No serious indirectness | Yes | ? | Low |
| Gait Retraining Exercise VS Proprioceptive Neuromuscular Facilitation + Exercise | Serious | No | No serious indirectness | Yes | ? | Low |
| Gait Retraining Exercise VS Whole Body Vibration Exercise | Serious | No | No serious indirectness | Yes | ? | Low |
| Gait Retraining Exercise VS Knee Exercise + Hip Exercise | Serious | No | No serious indirectness | Yes | ? | Low |
| Gait Retraining Exercise VS Foot Orthoses + Exercise | Serious | No | No serious indirectness | Yes | ? | Low |

| | | | | | | |
|---|---|---|---|---|---|---|
| Gait Retraining Exercise VS Hip Exercise | Serious | No | No serious indirectness | Yes | ? | Low |
| Gait Retraining Exercise VS Knee Brace + Exercise | Serious | No | No serious indirectness | Yes | ? | Low |
| Gait Retraining Exercise VS Supervised Exercise | Serious | No | No serious indirectness | Yes | ? | Low |
| Supervised Exercise VS Proprioceptive Neuromuscular Facilitation + Exercise | Serious | No | No serious indirectness | Yes | ? | Low |
| Supervised Exercise VS Whole Body Vibration Exercise | Serious | No | No serious indirectness | Yes | ? | Low |
| Supervised Exercise VS Knee Exercise + Hip Exercise | Serious | No | No serious indirectness | Yes | ? | Low |
| Supervised Exercise VS Foot Orthoses + Exercise | Serious | No | No serious indirectness | Yes | ? | Low |
| Supervised Exercise VS Hip Exercise | Serious | No | No serious indirectness | Yes | ? | Low |
| Supervised Exercise VS Knee Brace + Exercise | Serious | No | No serious indirectness | Yes | ? | Low |
| Knee Brace + Exercise VS Proprioceptive Neuromuscular Facilitation + Exercise | Serious | No | No serious indirectness | Yes | ? | Low |
| Knee Brace + Exercise VS Whole Body Vibration Exercise | Serious | No | No serious indirectness | Yes | ? | Low |
| Knee Brace + Exercise VS Knee Exercise + Hip Exercise | Serious | No | No serious indirectness | Yes | ? | Low |

| | | | | | | |
|---|---|---|---|---|---|---|
| Knee Brace + Exercise VS Foot Orthoses + Exercise | Serious | No | No serious indirectness | Yes | ? | Low |
| Knee Brace + Exercise VS Hip Exercise | Serious | No | No serious indirectness | Yes | ? | Low |
| Hip Exercise VS Proprioceptive Neuromuscular Facilitation + Exercise | Serious | No | No serious indirectness | Yes | ? | Low |
| Hip Exercise VS Whole Body Vibration Exercise | Serious | No | No serious indirectness | Yes | ? | Low |
| Hip Exercise VS Knee Exercise + Hip Exercise | Serious | No | No serious indirectness | Yes | ? | Low |
| Hip Exercis VS Foot Orthoses + Exercise | Serious | No | No serious indirectness | Yes | ? | Low |
| Foot Orthoses + Exercise VS Proprioceptive Neuromuscular Facilitation + Exercise | Serious | No | No serious indirectness | Yes | ? | Low |
| Foot Orthoses + Exercise VS Whole Body Vibration Exercise | Serious | No | No serious indirectness | Yes | ? | Low |
| Foot Orthoses + Exercise VS Knee Exercise + Hip Exercise | Serious | No | No serious indirectness | Yes | ? | Low |
| Knee Exercise + Hip Exercise VS Proprioceptive Neuromuscular Facilitation | Serious | No | No serious indirectness | Yes | ? | Low |
| Knee Exercise + Hip Exercise VS Whole Body Vibration Exercise | Serious | No | No serious indirectness | Yes | ? | Low |
| Whole Body Vibration Exercise VS Proprioceptive Neuromuscular Facilitation | Serious | No | No serious indirectness | Yes | ? | Low |

| Knee function measure with AKPS | | | | | | |
|---|---|---|---|---|---|---|
| Nontreatment VS Knee Exercise + Hip Exercise | Very serious | No | No | Yes | ? | Modorate |
| Nontreatment VS Knee Brace + Exercise | Serious | No | No serious indirectness | Yes | ? | Low |
| Nontreatment VS Target Exercise | Very serious | No | No | Yes | ? | Modorate |
| Nontreatment VSFeedback Exercise | Serious | No | No serious indirectness | Yes | ? | Low |
| Nontreatment VS Supervised Exercise | Serious | No | No serious indirectness | Yes | ? | Low |
| Nontreatment VS Blood Flow Restriction Exercise | Serious | No | No serious indirectness | Yes | ? | Low |
| Nontreatment VS Proprioceptive Neuromuscular Facilitation + Exercise | Serious | No | No serious indirectness | Yes | ? | Low |
| Nontreatment VS KinesioTaping + Exercise | Serious | No | No serious indirectness | Yes | ? | Low |
| Nontreatment VS Education | Serious | No | No serious indirectness | Yes | ? | Low |
| Nontreatment VS Whole Body Vibration Exercise | Serious | No | No serious indirectness | Yes | ? | Low |
| Nontreatment VS General Exercise | Very serious | No | No | Yes | ? | Modorate |

| | | | | | | |
|---|---|---|---|---|---|---|
| Nontreatment VS Knee Exercise | Very serious | No | No | Yes | ? | Modorate |
| Nontreatment VS Hip Exercise | Serious | No | No serious indirectness | Yes | ? | Low |
| Hip Exercise VS Knee Exercise + Hip Exercise | Serious | No | No serious indirectness | Yes | ? | Low |
| Hip Exercise VS Knee Brace + Exercise | Serious | No | No serious indirectness | Yes | ? | Low |
| Hip Exercise VS Target Exercise | Serious | No | No serious indirectness | Yes | ? | Low |
| Hip Exercise VS Feedback Exercise | Serious | No | No serious indirectness | Yes | ? | Low |
| Hip Exercise VS Supervised Exercise | Serious | No | No serious indirectness | Yes | ? | Low |
| Hip Exercise VS Blood Flow Restriction Exercise | Serious | No | No serious indirectness | Yes | ? | Low |
| Hip Exercise VS Proprioceptive Neuromuscular Facilitation + Exercise | Serious | No | No serious indirectness | Yes | ? | Low |
| Hip Exercise VS KinesioTaping + Exercise | Serious | No | No serious indirectness | Yes | ? | Low |
| Hip Exercise VS Education | Serious | No | No serious indirectness | Yes | ? | Low |
| Hip Exercise VS Whole Body Vibration Exercise | Serious | No | No serious indirectness | Yes | ? | Low |

| | | | | | | |
|---|---|---|---|---|---|---|
| Hip Exercise VS General Exercise | Serious | No | No serious indirectness | Yes | ? | Low |
| Hip Exercise VS Knee Exercise | Very serious | No | No | Yes | ? | Modorate |
| Knee Exercise VS Knee Exercise + Hip Exercise | Serious | No | No serious indirectness | Yes | ? | Low |
| Knee Exercise VS Knee Brace + Exercise | Serious | No | No serious indirectness | Yes | ? | Low |
| Knee Exercise VS Target Exercise | Serious | No | No serious indirectness | Yes | ? | Low |
| Knee Exercise VS Feedback Exercise | Serious | No | No serious indirectness | Yes | ? | Low |
| Knee Exercise VS Supervised Exercise | Serious | No | No serious indirectness | Yes | ? | Low |
| Knee Exercise VS Blood Flow Restriction Exercise | Serious | No | No serious indirectness | Yes | ? | Low |
| Knee Exercise VS Proprioceptive Neuromuscular Facilitation + Exercise | Serious | No | No serious indirectness | Yes | ? | Low |
| Knee Exercise VS KinesioTaping + Exercise | Serious | No | No serious indirectness | Yes | ? | Low |
| Knee Exercise VS Education | Serious | No | No serious indirectness | Yes | ? | Low |
| Knee Exercise VS Whole Body Vibration Exercise | Serious | No | No serious indirectness | Yes | ? | Low |

| | | | | | | |
|---|---|---|---|---|---|---|
| Knee Exercise VS General Exercise | Serious | No | No serious indirectness | Yes | ? | Low |
| General Exercise VS Knee Exercise + Hip Exercise | Serious | No | No serious indirectness | Yes | ? | Low |
| General Exercise VS Knee Brace + Exercise | Very serious | No | No | Yes | ? | Modorate |
| General Exercise VS Target Exercise | Very serious | No | No | Yes | ? | Modorate |
| General Exercise VS Feedback Exercise | Very serious | No | No | Yes | ? | Modorate |
| General Exercise VS Supervised Exercise | Very serious | No | No | Yes | ? | Modorate |
| General Exercise VS Blood Flow Restriction Exercise | Very serious | No | No | Yes | ? | Modorate |
| General Exercise VS Proprioceptive Neuromuscular Facilitation + Exercise | Very serious | No | No | Yes | ? | Modorate |
| General Exercise VS KinesioTaping + Exercise | Very serious | No | No | Yes | ? | Modorate |
| General Exercise VS Education | Serious | No | No serious indirectness | Yes | ? | Low |
| General Exercise VS Whole Body Vibration Exercise | Very serious | No | No | Yes | ? | Modorate |
| Whole Body Vibration Exercise VS Knee Exercise + Hip Exercise | Serious | No | No serious indirectness | Yes | ? | Low |

| | | | | | | |
|---|---|---|---|---|---|---|
| Whole Body Vibration Exercise VS Knee Brace + Exercise | Serious | No | No serious indirectness | Yes | ? | Low |
| Whole Body Vibration Exercise VS Target Exercise | Serious | No | No serious indirectness | Yes | ? | Low |
| Whole Body Vibration Exercise VS Feedback Exercise | Serious | No | No serious indirectness | Yes | ? | Low |
| Whole Body Vibration Exercise VS Supervised Exercise | Serious | No | No serious indirectness | Yes | ? | Low |
| Whole Body Vibration Exercise VS Blood Flow Restriction Exercise | Serious | No | No serious indirectness | Yes | ? | Low |
| Whole Body Vibration Exercise VS Proprioceptive Neuromuscular Facilitation | Serious | No | No serious indirectness | Yes | ? | Low |
| Whole Body Vibration Exercise VS KinesioTaping + Exercise | Serious | No | No serious indirectness | Yes | ? | Low |
| Whole Body Vibration Exercise VS Education | Serious | No | No serious indirectness | Yes | ? | Low |
| Education VS Knee Exercise + Hip Exercise | Serious | No | No serious indirectness | Yes | ? | Low |
| Education VS Knee Brace + Exercise | Very serious | No | No | Yes | ? | Modorate |
| Education VS Target Exercise | Serious | No | No serious indirectness | Yes | ? | Low |
| Education VS Feedback Exercise | Serious | No | No serious indirectness | Yes | ? | Low |

| | | | | | | |
|---|---|---|---|---|---|---|
| Education VS Supervised Exercise | Serious | No | No serious indirectness | Yes | ? | Low |
| Education VS Blood Flow Restriction Exercise | Serious | No | No serious indirectness | Yes | ? | Low |
| Education VS Proprioceptive Neuromuscular Facilitation + Exercise | Serious | No | No serious indirectness | Yes | ? | Low |
| Education VS KinesioTaping + Exercise | Serious | No | No serious indirectness | Yes | ? | Low |
| KinesioTaping + Exercise VS Knee Exercise + Hip Exercise | Serious | No | No serious indirectness | Yes | ? | Low |
| KinesioTaping + Exercise VS Knee Brace + Exercise | Serious | No | No serious indirectness | Yes | ? | Low |
| KinesioTaping + Exercise VS Target Exercise | Serious | No | No serious indirectness | Yes | ? | Low |
| KinesioTaping + Exercise VS Feedback Exercise | Serious | No | No serious indirectness | Yes | ? | Low |
| KinesioTaping + Exercise VS Supervised Exercise | Serious | No | No serious indirectness | Yes | ? | Low |
| KinesioTaping + Exercise VS Blood Flow Restriction Exercise | Serious | No | No serious indirectness | Yes | ? | Low |
| KinesioTaping + Exercise VS Proprioceptive Neuromuscular Facilitation | Serious | No | No serious indirectness | Yes | ? | Low |
| Proprioceptive Neuromuscular Facilitation + Exercise VS Knee Exercise + Hip | Serious | No | No serious indirectness | Yes | ? | Low |

| | | | | | | |
|---|---|---|---|---|---|---|
| Proprioceptive Neuromuscular Facilitation + Exercise VS Knee Brace + Exercise | Serious | No | No serious indirectness | Yes | ? | Low |
| Proprioceptive Neuromuscular Facilitation + Exercise VS Target Exercise | Serious | No | No serious indirectness | Yes | ? | Low |
| Proprioceptive Neuromuscular Facilitation + Exercise VS Feedback Exercise | Serious | No | No serious indirectness | Yes | ? | Low |
| Proprioceptive Neuromuscular Facilitation + Exercise VS Supervised Exercise | Serious | No | No serious indirectness | Yes | ? | Low |
| Proprioceptive Neuromuscular Facilitation + Exercise VS Blood Flow Restriction | Serious | No | No serious indirectness | Yes | ? | Low |
| Blood Flow Restriction Exercise VS Knee Exercise + Hip Exercise | Serious | No | No serious indirectness | Yes | ? | Low |
| Blood Flow Restriction Exercise VS Knee Brace + Exercise | Serious | No | No serious indirectness | Yes | ? | Low |
| Blood Flow Restriction Exercise VS Target Exercise | Serious | No | No serious indirectness | Yes | ? | Low |
| Blood Flow Restriction Exercise VS Feedback Exercise | Serious | No | No serious indirectness | Yes | ? | Low |
| Blood Flow Restriction Exercise VS Supervised Exercise | Serious | No | No serious indirectness | Yes | ? | Low |
| Blood Flow Restriction Exercise VS Knee Exercise + Hip Exercise | Serious | No | No serious indirectness | Yes | ? | Low |
| Blood Flow Restriction Exercise VS Knee Brace + Exercise | Serious | No | No serious indirectness | Yes | ? | Low |

| | | | | | | |
|---|---|---|---|---|---|---|
| Blood Flow Restriction Exercise VS Target Exercise | Serious | No | No serious indirectness | Yes | ? | Low |
| Blood Flow Restriction Exercise VS Feedback Exercise | Serious | No | No serious indirectness | Yes | ? | Low |
| Blood Flow Restriction Exercise VS Knee Exercise + Hip Exercise | Serious | No | No serious indirectness | Yes | ? | Low |
| Feedback Exercise VS Knee Brace + Exercise | Serious | No | No serious indirectness | Yes | ? | Low |
| Feedback Exercise VS Target Exercise | Serious | No | No serious indirectness | Yes | ? | Low |
| Target Exercise VS Knee Exercise + Hip Exercise | Serious | No | No serious indirectness | Yes | ? | Low |
| Target Exercise VS Knee Brace + Exercise | Serious | No | No serious indirectness | Yes | ? | Low |
| Knee Brace + Exercise VS Knee Exercise + Hip Exercise | Serious | No | No serious indirectness | Yes | ? | Low |

Abbreviations: GRADE = Grading of Recommendations Assessment, Development, and Evaluationb a
Populations, treatments and outcomes measures followed those used in clinical practice, hence there was no indication of indirectness in the evidence.

### Web appendix8 Data analyses, treatment level results

Figure1 Ranking of treatment strategies

Figure1A Primary outcome, VAS

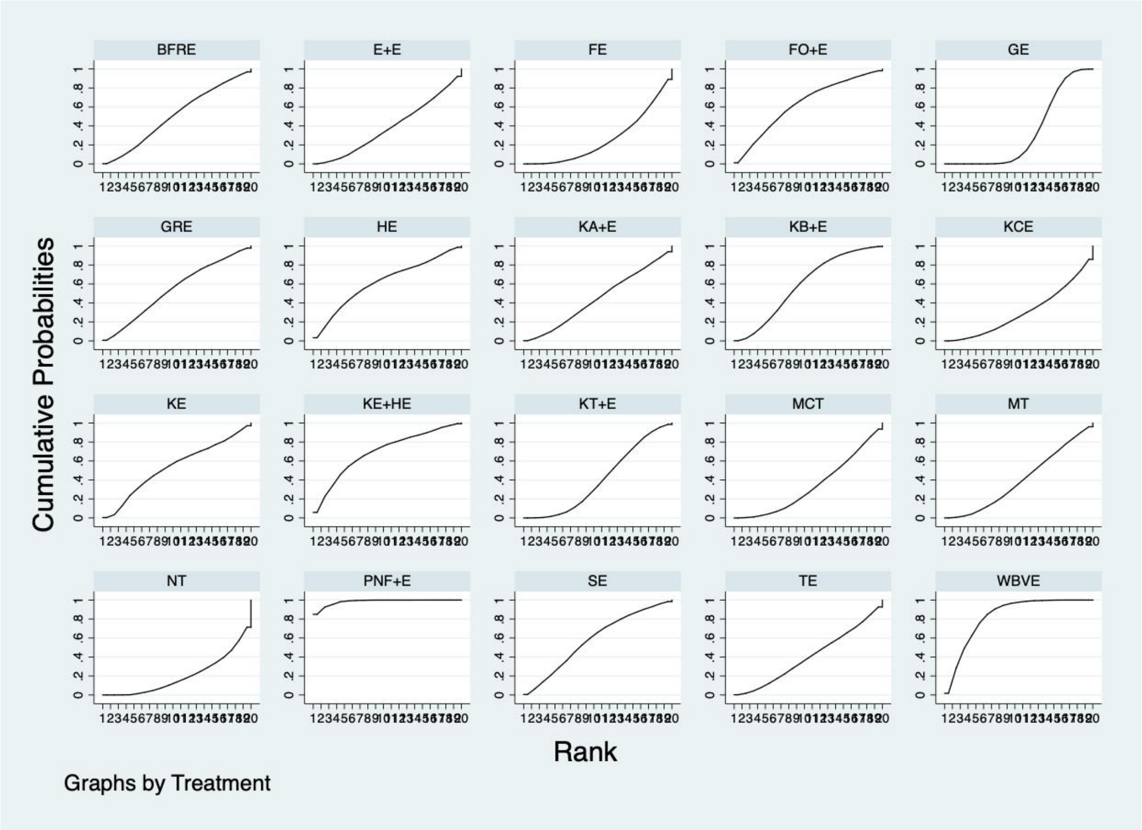

Ranking of treatment strategies based on probability of their protective effects on outcome of VAS according to the cumulative ranking area (SUCRA). Larger probability, stronger protective effects

Figure1B Secondary outcomes, AKPS

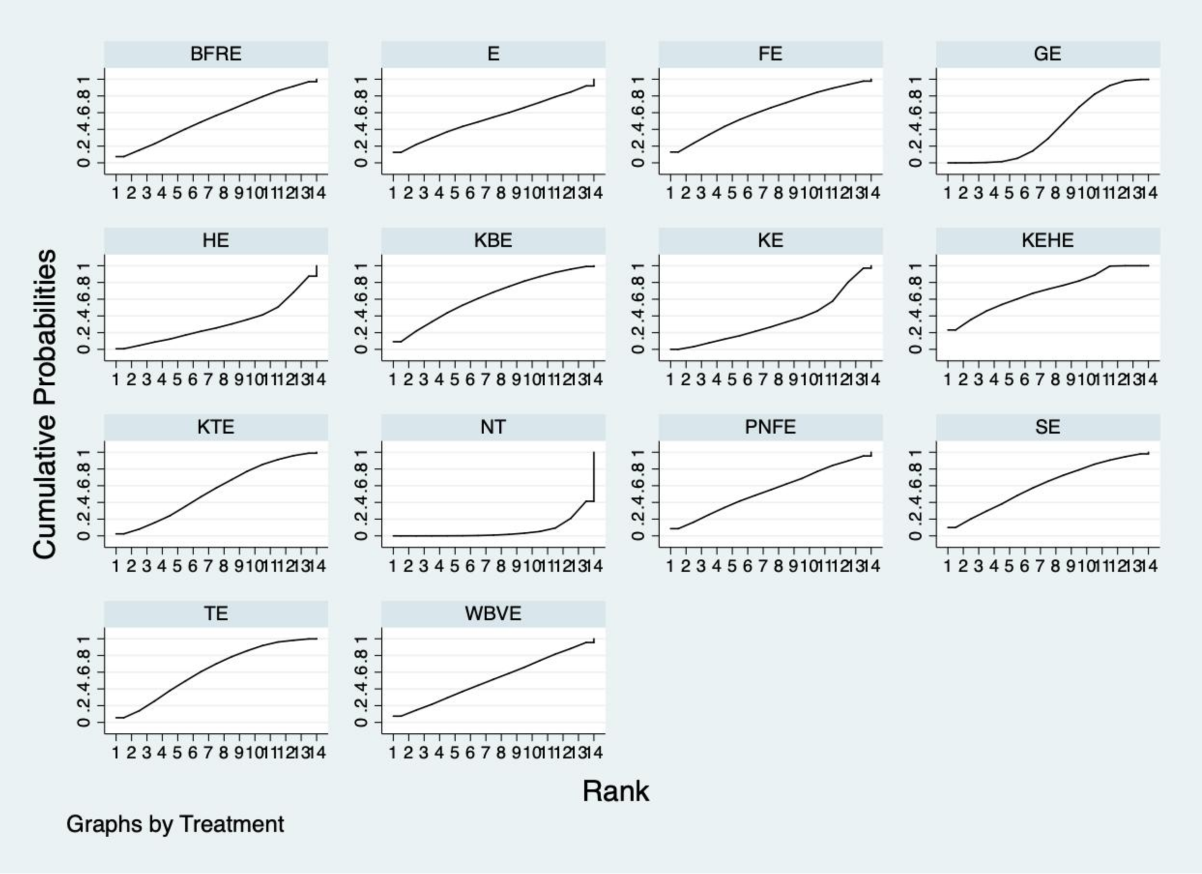

Ranking of treatment strategies based on probability of their protective effects on outcome of VAS according to the cumulative ranking area (SUCRA). Larger probability, stronger protective effects.

GE=General Exercise; GRE=Gait Retraining Exercise; E+E=Education + Exercise; FO+E=Foot Orthoses + Exercise; FE=Feedback Exercise; MT+E=Manipulation Treatment +Exercise; KE=Keen Exercise; KB+E=Knee Brace + Exercise; KT+E=KinesioTaping + Exercise; NT=Nontreatment; KA+E=Knee Arthroscopy + Exercise; E=Education; BFRE=Blood Flow Restriction Exercise; WBVE=Whole Body Vibration Exercise; PNF+E=Proprioceptive Neuromuscular Facilitation + Exercise; MCT=Motor Control Training; TE=Target Exercise; SE=Supervised Exercise; KCE= Kinetic Chain Exercise; HE=Hip Exercise; KE+HE=Knee Exercise + Hip Exercise

#### Web appendix9. A recommended exercise prescription

<table>
<tr><td></td><td colspan="2">Recommended exercise therapy</td></tr>
<tr><td>Risk factor</td><td colspan="2">Treatment</td></tr>
<tr><td>Muscle weakness</td><td colspan="2">❖Hip-Focused Exercises<br><br>The hip-focused exercises were based on previous studies and consisted of side-lying hip abduction, hip external rotation (clam shell), and prone hip extension. These exercises were intended to maximally isolate the hip abductors, extensors, and external rotators.<br><br>❖Knee-Focused Exercises<br><br>The knee-focused exercise regimens were based on previous studies and were intended to maximally isolate the quadriceps muscles. The exercises consisted of straight-leg raises in the supine position, supine terminal knee extensions (from 10 of flexion to full extension), and a mini-squat (45 of flexion) with the back supported against the wall (to reduce stabilizing requirements from the hip muscles).</td></tr>
<tr><td rowspan="3">PFJ alignment</td><td rowspan="3">Individual evaluation</td><td>❖Foot orthoses</td></tr>
<tr><td>❖Kinesiotaping</td></tr>
<tr><td>❖Knee brace</td></tr>
<tr><td>Lack of self-management</td><td colspan="2">❖Education<br><br>It covered: pain management; how to modify physical activity using pacing and load management strategies; information on optimal knee alignment during daily tasks; and responses to questions from the adolescent or the parents.</td></tr>
<tr><td>Tips</td><td colspan="2">❖Guiding principles<br><br>●Dosage is chosen in which the last repetitions are challenging but</td></tr>
</table>

| | |
|---|---|
| | the quality of movement is maintained.<br>●Dosage is individually adjusted once per week by the physiotherapist.<br>❖Progression (all exercises):<br>●The number of repetitions is increased from 3 sets of 10 repetitions to a maximum of 3 sets of 20 repetitions.<br>●Thereafter resistance is increased using a weight cuff or resistance tubing (see individual exercise).<br>o Weight cuffs are available in 0.5 kg increments.<br>o Resistance tubing is selected from 3 possible variants. In order of increasing resistance: red (medium), green (heavy), black (special heavy)<br>❖Other details:<br>●Repetitions performed dynamically over 2-3 seconds<br>●2-second pause between repetitions.<br>●30-second pause between sets.<br>●Minimum one rest day between sessions |
| The need for an individual exercise therapy approach to PFP treatment. Clinicians can devise an individual prescription. Personalized exercise prescriptions should be further studied by clinicians or therapists in the future. | |
